# Identifying efficient linkage strategies for HIV self-testing (IDEaL): a study protocol for an individually randomized control trial

**DOI:** 10.1101/2022.12.23.22283834

**Authors:** Kathryn Dovel, Kelvin Balakasi, Julie Hubbard, Khumbo Phiri, Brooke E. Nichols, Thomas J. Coates, Michal Kulich, Elijah Chikuse, Sam Phiri, Lawrence Long, Risa Hoffman, Augustine Choko

## Abstract

**Introduction:** Men in sub-Saharan Africa are less likely than women to initiate antiretroviral therapy (ART) and are more likely to have longer cycles of disengagement from ART programs. Treatment interventions that meet the unique needs of men are needed, but they must be scalable. We will conduct a study to test the impact of various interventions on six-month retention in ART programs among men living with HIV who are not currently engaged in care.

**Methods and Analysis:** We will conduct a programmatic, individually randomized, non-blinded, non-inferiority controlled trial. “Non-engaged” men will be randomized 1:1:1 to either a Stepped, Low-Intensity, or High-Intensity arm. In the Stepped arm, intervention activities build in intensity over time for those who do not reengage in care with the following steps: 1) one-time male-specific counseling + facility navigation ⟶ 2) ongoing male mentorship + facility navigation ⟶ outside-facility ART initiation + male-specific counseling + facility navigation for follow-up ART visits. The Low-Intensity Intervention includes one-time male-specific counseling + facility navigation only and the High-Intensity Intervention offers immediate outside-facility ART initiation + male-specific counseling + facility navigation for follow-up ART visits. Our primary outcome is 6-month retention in care. Secondary outcomes include cost-effectiveness and rates of adverse events.

**Ethics and Dissemination:** The Institutional Review Board of the University of California, Los Angeles and the National Health Sciences Research Council in Malawi have approved the trial protocol. Findings will be disseminated rapidly in national and international forums, as well as in peer-reviewed journals and are expected to provide urgently needed information to other countries and donors.

**ARTICLE SUMMARY:** *Strengths and limitations:*

- IDEAL provides male-specific differentiated models of care aimed to improve men’s ART outcomes. We specifically focus on building trusting relationships with health care workers and developing client-led, individualized strategies to overcome barriers to care.
- IDEAL will test the impact of a stepped intervention for men. This approach promises to improve the efficiency and reach of HIV programs for men as the highest-resource interventions will only be received by the minority of men who are most in need.
- IDEAL develops and tests male-specific counseling curriculum that, if effective, could easily be taken to scale. Findings from the study will identify critical components for male-specific counseling, especially among men who struggle to be retained in HIV care.

## INTRODUCTION

Men in sub-Saharan Africa (SSA) are underrepresented in HIV programs.^1^ Men are less likely than women to know their HIV status and to initiate antiretroviral therapy (ART), and more likely to face treatment interruptions once in care.^2^ Men are particularly prone to cycling through ART programs, with more frequent stop-start instances and longer periods outside of care as compared to women.^5-8^ Engagement in ART programs is not static – many men and women cycle through care, starting and stopping HIV care multiple times throughout their lifetime.^3,4^ Men who disengage from HIV programs (either after testing HIV-positive or after enrolling in HIV care services) are frequently described as a difficult and ‘hard-to-reach’ population.^9,10^ However, growing evidence suggests that men desire HIV services^11,12^ but encounter multiple health systems barriers to care that make it impossible to stay in care long-term.^13^ There is an urgent need to develop client-centered strategies tailored to men that facilitate men’s engagement and re-engagement in HIV treatment programs.

Some men may require male-specific interventions to facilitate engagement in HIV care. Men have less exposure to HIV services than women^13,14^ and work demands may conflict with ART clinic schedules.^15,16^ Difficult interactions with health care workers (HCWs) can also prevent men from engaging or re-engaging in care.^17,18^ Furthermore, most ART counseling curricula do not target men and often lack the client-centered counseling needed to develop internal motivation to engage and stay engaged in care.

Differentiated service delivery models (DSD) are now being developed to improve men’s ART engagement throughout SSA.^19-21^ As DSDs for men are developed, it is critical that strategies be feasible and cost-effective to allow scale-up. A “one size fits all” model is not as effective as more nuanced approaches.^22-24^ Stepped interventions increase in intensity over time and are purposively designed to address prevailing barriers in the target population in order to positively affect the desired outcome.^25,26^ An incremental, stepped approach may be the most appropriate and scalable way to improve men’s care in low-resource settings. Men are not homogeneous: some men may require minimal support to engage in care, while others may require extensive support. Stepped interventions allow programs to target the highest-resource interventions to the minority of men who need them most.

The *Identifying efficient linkage strategies for HIV self-testing (IDEaL) Trial* is an individually randomized control trial aimed to test the impact of various interventions on ART (re-) engagement and six-month retention among men living with HIV who are not currently engaged in HIV care in Malawi. We will compare a Stepped intervention against Low-Intensity and High-Intensity interventions to assess the impact of the Stepped intervention on men’s use of ART services over time (see Supporting Information S1).

## METHODS AND ANALYSIS

### Objectives

Our primary objective is to test the effect of a male-specific, Stepped intervention on men’s 6-month retention in ART care compared to male-specific Low-Intensity and High-Intensity interventions (retention is defined as <28-days late for their ART appointment). Secondary objectives are to understand the impact of a Stepped intervention on: (1) ART initiation; (2) the presence of adverse events (i.e., unwanted disclosure, end of relationship, or intimate partner violence (IPV)); (3) intervention acceptability; and (4) cost-effectiveness.

### Trial Design

IDEaL is a programmatic, individually randomized, non-blinded, non-inferiority controlled trial design. We will recruit men from 15 high-burden health facilities in Malawi using medical chart reviews to identify men who are living with HIV but not engaged in HIV care.

### Randomization

Individual men will be block randomized by a biostatistician using a 1:1:1 ratio to either the Stepped, Low-Intensity, or High-Intensity study arm using a computer-generated program. Participants will be randomized in blocks of 3 and 6, depending on the number of men available for recruitment at each facility. After enrolling in the trial and completing a baseline survey, men will be assigned to a study ID based on the randomization list. Study ID’s will be linked with the pre-assigned blocked randomization and pre-loaded into the tablet device, but will be unknown to the study staff until survey and randomization modules are completed and saved, ensuring randomization cannot be manipulated by the study staff. Once finalized, the randomization results will appear on the tablet device as a picture, and will be shown to the participant to maximize transparency and study buy-in.

### Interventions

The effectiveness of the Stepped Intervention will be compared to a Low-Intensity Intervention (one time male-specific counseling + facility navigation) and to a High-Intensity Intervention (outside-facility ART initiation + male-specific counseling + facility navigation for follow-up ART visits). Across all arms, men who do not (re-)engage in ART will continue receiving follow-up visits for up to three months, depending on preferences of the client. The number of intervention visits delivered for each participant will be documented.

#### Arm 1: Stepped Arm

The Stepped arm will build in intensity over time for those who have not (re-)engaged in care 14-days after enrollment, or who do not return for their first ART follow-up appointment after (re-)engagement (see Fig 1). Individuals will move to the next “step” every 2-weeks, moving from the lightest to the most intensive interventions over the course of X weeks until (re-)engagement has been achieved (as defined by starting or restarting ART). The Stepped arm includes the following steps:

**Figure 1:**
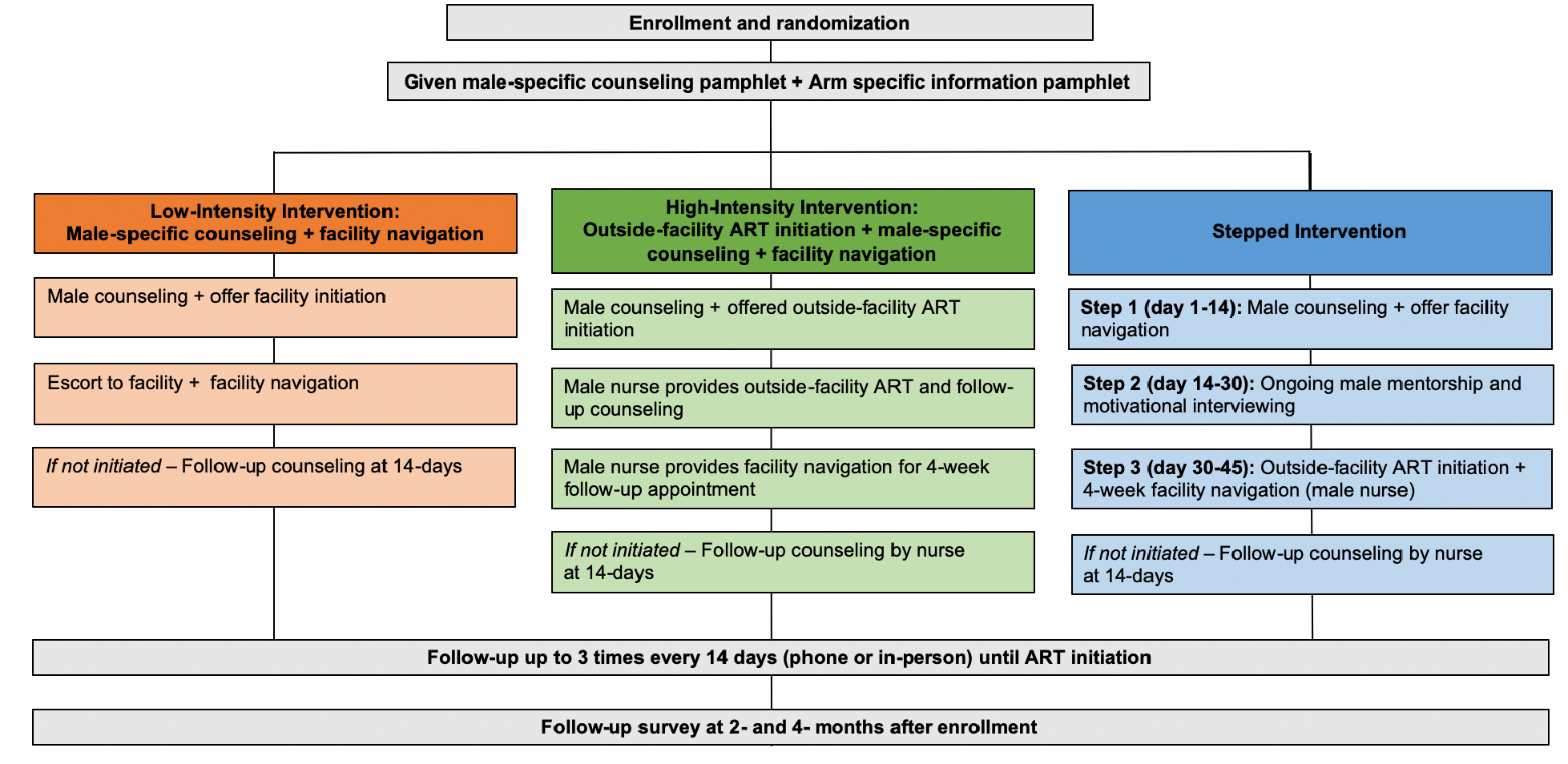
Trial Design.

##### Step 1: Male-Specific Counseling + Facility Navigation

Participants will be traced in the community and receive a one-time, one-on-one, intensive counseling session in the community by a lower-level cadre male HIV counselor (called Patient Supporter in Malawi) using a male-specific counseling curriculum developed specifically for this trial.

Ministry of Health counseling materials will be adapted to develop the male-specific counseling curriculum, based on formative in-depth interviews, focus group discussions, and a systematic literature review. Adaptations will include exploring topics of most concern to men in Malawi (i.e., earning money while HIV-positive, side effects and concerns regarding lifelong medication, ART as a tool to provide and care for family, etc.). The materials will also include language and pictures that resonate with men (i.e., emphasizing how HIV and HIV services interact with men’s strength, responsibility, planning for the future), and male-specific case studies of challenges men face and how they overcome them. The adapted male-specific counseling curriculum will be developed into a standardized counseling flip chart to promote consistent and accurate implementation of the intervention.

Men who wish to (re-)engage in care will be provided with facility navigation, including being escorted to the facility (if desired) and being oriented to the ART clinic within the health facility by the same counselor who delivers the intervention. Men will access all ART clinical services at a health facility of their choice (but counseling described above will be provided in the community).

##### Step 2: Ongoing Motivational Interviewing + Facility Navigation

Men who do not (re-)engage in care (either have not engaged ART within 14 days of enrollment or do (re-)engage ART but are ≥7-days late for a follow-up appointment) will move to the next ‘step’ of the intervention, which adds ongoing motivational interviewing to their package of activities. Motivational interviewing is a validated form of counseling aimed to help individuals identify barriers to a desired outcome and develop personalized solutions.^27^ Mentors will work with participants to: (1) build self-efficacy, (2) identify internal motivations for the desired behavior, and (3) establish strategies and short- and long-term goals needed to reach ART initiation and retention. A male mentor specifically trained in motivational interviewing will provide ongoing, one-on-one in-depth counseling, motivational interviewing, and general “check-ins” approximately twice within a two-week period. The mentor will not necessarily be HIV-positive (unlike other mentorship models) as the Malawi Ministry of Health has moved away from HIV-positive peer mentor cadres. However, they will be experienced in HIV counseling and trained on male-specific needs. Motivational interviewing will take place in a location preferred by the participant, likely in the community. Participants who choose to (re)engage in care will be given facility navigation.

##### Step 3: Outside-facility ART initiation + male-specific counseling + facility navigation

Men who are not engaged after Step 2 (either have not engaged in ART care within 14 days of receiving the first motivational interviewing or have engaged but are ≥7-days late for a follow-up ART appointment) will be offered outside-facility ART initiation by a male nurse affiliated with the nearest public health facility. The nurse will meet the participant at a convenient and private location of his choice. Outside-facility ART will be male-friendly, including components like early hours and convenient locations.

The nurse will provide client-centered counseling by reviewing key points from male-specific counseling that are most relevant to the individual, and conduct WHO staging. Individuals classified as WHO Stage 3 or 4 will not be eligible for outside-facility ART and will be escorted to the nearest public health facility for additional services. Participants classified as WHO Stage 1 or 2 will be given ART for same day initiation. The nurse will schedule a 4-week follow-up ART refill appointment at the health facility of the man’s choice. At the time of the follow-up appointment, the nurse will provide facility navigation to facilitate a positive experience engaging in or returning to care and serve as the ART provider for this appointment to facilitate service continuity and familiarity.

#### Arm 2: Low-Intensity Arm: Male-specific counseling + facility navigation

Participants randomized to the Low-Intensity Arm will be traced in the community and receive a one-time, one-on-one, intensive counseling session in the community by a lower-level cadre male HIV counselor. The counseling session will use the same male-specific counseling curriculum described in Step 1 of the Stepped Arm. For men who do not (re-)engage in care, follow-up counseling will be offered approximately every two weeks until individuals engage in care or inform the counselor that they do not wish to be contacted. Those who choose to engage in ART following male-specific counseling will be given facility navigation for their first ART appointment.

#### Arm 3: High-Intensity Arm: Outside-facility ART initiation + male-specific counseling + facility navigation

The High-Intensity Arm will offer the male-specific counseling described above, outside-facility ART initiation, and facility navigation for their 4-week follow-up appointment. After receiving male-specific counseling, participants interested in outside-facility ART initiation will be referred to a male study nurse who will meet men one-on-one at convenient times and locations. Those who decline outside facility ART initiation will be referred immediately to the nearest health facility and receive facility navigation on a day and time that is convenient for them. For outside-facility ART, the nurse will provide client-centered counseling, reviewing key points from male-specific counseling that are most relevant and/or of greatest concern to the individual, conduct WHO staging, and for those in WHO Stage 1 or 2, provide same day ART initiation (as described in the Stepped Arm). The nurse will provide facility navigation for the 4-week ART refill appointment. Men who do not (re-)engage in care will be offered biweekly follow-up counseling at times and intervals determined by participants’ preferences until they engage in care or inform the nurse that they no longer wish to be contacted.

### Trial setting

The study will take place in central and southern Malawi. Malawi has an HIV prevalence of 9.6%^28^ and, of the estimated 330,000 men living with HIV in the country, 54,500 are not in care.^29^ Men in Malawi live in primarily rural settings, are self-employed, subsistence farmers, the minority have regular access to a private phone, and most are highly mobile.^30,31^

### Population

We will recruit men from 15 high-burden health facilities in Malawi, using medical chart reviews to identify men living with HIV who are not engaged in HIV care. Study facilities will vary by facility type (hospital/health center), management (public/mission), location (rural/urban), and region (central/southern Malawi).

Eligibility criteria for men include: (1) ≥15years of age; (2) live in facility catchment area; and (3) tested HIV-positive and either (a) self-report having not yet initiated ART within 7-days of testing HIV-positive, (b) initiated ART but are at risk of immediate default (i.e., ≥7-days late for their 30-day ART refill appointment), or (c) initiated ART and attended their first refill appointment but later defaulted (i.e. ≥28-days late to care). For those who never initiated ART and do not have proof of a confirmatory HIV test, study staff will offer an HIV self-test kit prior to enrollment, to confirm a positive HIV status. Those who choose to initiate ART will receive the standard Determine and Unigold confirmatory tests prior to ART initiation, following routine care.

### Study outcomes

The primary outcome is the proportion of men who are retained in ART care 6-months after (re-) engagement. Secondary outcomes include: (1) ART initiation; (2) adverse events experienced by men or their female partners (i.e., unwanted disclosure, end of relationship, or intimate partner violence(IPV); (3) intervention acceptability; and (4) cost-effectiveness. ART retention outcomes will be measured through medical chart reviews, while secondary outcomes will be measured through self-reports. Process outcomes include: (1) the proportion of men who were successfully traced; (2) the proportion of eligible men who consented to participate; (3) men’s experience with the intervention; and (4) the quality of the intervention.

#### Sample size considerations

We powered the study to detect differences in 6-month retention between Stepped and Low-Intensity Arms, and the Stepped and High-Intensity Arms. Based on previous trials, we assumed that 40% of men in the Low-Intensity Arm, 60% in the Stepped Arm, and 80% in the High-Intensity Arm will engage in ART and be retained at 6-months. Any man lost to follow-up will be treated as a failure for the outcome evaluation. With 181 men per arm and 20% loss to follow-up from the study in all arms, the power for detecting the specified difference between Stepped and Low Intensity arms will be 0.8 and the power to detect the specified difference between Stepped and High-Intensity arms will be >0.99, taking into account that two comparisons are made. The calculation is based on asymptotic normality of log odds ratio.^32^ We need to enroll and randomize 181 men per arm (a total of 543 men living with HIV).

### Data Collection

Study recruitment, enrollment, and data collection will be conducted by study staff, who are distinct from local HCWs implementing the interventions.

#### Recruitment

Men will be identified through both medical register reviews and in-person recruitment at participating health facilities. Various medical charts will be reviewed to identify different types of eligible men: HIV testing and counseling (HTC) to identify men who tested HIV positive but never initiated ART; client follow-up registers to identify those who initiated but never returned for their first ART appointment, or those who defaulted from care; and index counseling and testing (ICT) registers to identify male partners of female ART clients (Figure 2). In-person recruitment will involve screening men at outpatient departments (OPD) because our previous research has found that men in Malawi frequent OPD settings for health needs,^12^ and our formative work suggests that men who disengage from ART services still frequent the OPD for care. In-person recruitment will be used for all client types.

**Fig 2.**
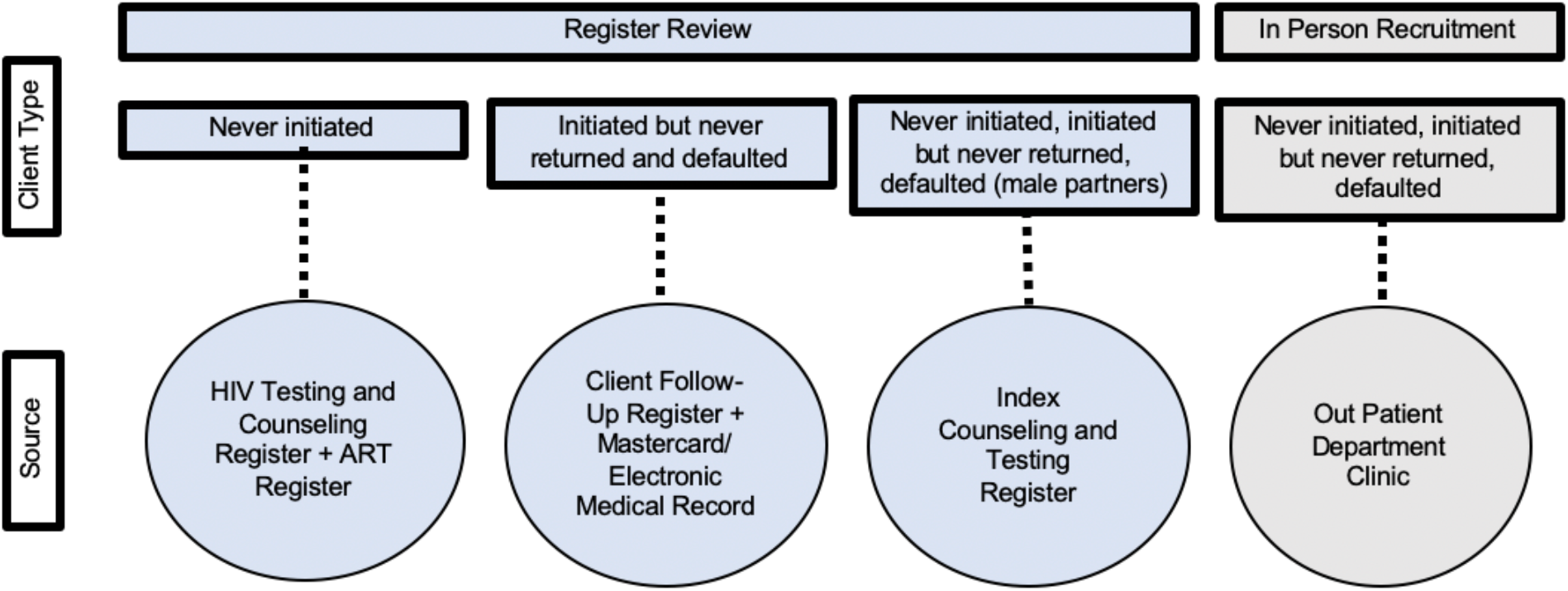
Recruitment sources and ART disengagement criteria by recruitment type.

#### Tracing and Eligibility Screening

Study staff will trace potential participants identified through medical chart reviews via phone (if available) or home visits based on tracing data provided in medical documentation. All potential participants will be traced up to three times before being considered lost to follow-up. All screening and enrollment processes will take place in-person.

#### Consent, Enrollment, and Baseline Survey

Men who are eligible for the study will complete written informed consent and complete a baseline survey immediately following enrollment. The baseline survey will collect data on key demographic variables (marital status, number of children, employment, self-rated health) and previous engagement with HIV and non-HIV health services. All surveys will be conducted in the local language (Chichewa) by trained study staff using electronic tablets. Surveys will be programmed using SurveyCTO software (http://www.surveycto.com).

#### Follow-Up Data

Study staff will administer follow-up surveys at 2- and 4-months after enrollment. Follow-up surveys will measure exposure to (and acceptability of) the interventions, changes in key demographics since enrollment (i.e., marital status, number of children, employment, self-rated health), any adverse events since enrollment (i.e., unwanted status disclosure, termination of relationship due to the intervention), and use of ART services. The location and specific time of the follow-up survey will be based on participant preference.

Medical chart reviews will be conducted to assess men’s engagement with ART services 6-months after study enrollment. Individuals without a medical chart outcome will be followed-up in person and their health passport, a pocket medical record where providers record data during health visits, will be reviewed to collect the ART outcome. Men who cannot be reached or are lost to follow-up in any arm will be counted as failures for that specific ART outcome of interest: (re-)engagement or 6-month retention).

### Patient and Public Involvement

Extensive formative work informed the development of the study protocol including in-depth interviews, focus group discussions, and a systematic literature review. The study protocol and tools were presented to Ministry of Health, national stakeholders and implementing partners (see Supporting Information S2).

### Cost data

The average cost per successful outcome (6-month retention) will be calculated and compared across arms incrementally. We will use micro-costing methods by creating an inventory of the resources used to achieve the observed study outcomes including: (1) standard counseling interactions (staff cadre, training received, duration of interaction and distance from facility travelled where applicable); (2) motivational interviewing interactions (staff cadre, training received, duration of interaction and distance from facility travelled where applicable); (3) provider interactions (staff cadre, training received, duration of interaction and distance from facility travelled where applicable); and (4) cost of reminder messages sent, when messages are delivered telephonically instead of in person. For each study patient, the quantity (number of units) of resources used will be determined. Costs will be measured from the health care provider perspective. Unit costs of resources, which are not human subject data, will be obtained from external suppliers and the health facilities’ finance and procurement records and multiplied by the resource usage data to provide an average cost per study patient in each study arm. A cost-effectiveness analysis will be conducted by dividing the incremental cost between two arms by the incremental effectiveness (number of people retained at 6-months) in the respective arms.

### Analysis plan

Data analysis will be conducted in R: A Language and Environment for Statistical Computing (R Foundation for Statistical Computing). We will use the Consolidated Standards of Reporting Trials (CONSORT) standards for reporting trial outcomes.^33^ Using an intention-to-treat analysis, all randomized men will be included in the analysis of the primary outcome; men with missing outcome assessment due to loss to follow-up will be treated as outcome failures. We will calculate descriptive statistics, including mean, standard deviation, range, and frequency distributions for the demographic characteristics and study outcomes by study arm. The primary outcome and all other binary outcomes will be analyzed by logistic regression models with key sociodemographic variables included as covariates (i.e., age and marital status). The intervention effects will be tested by pairwise Wald tests with Bonferroni adjustment. Confidence intervals for intervention effects will be calculated by profile likelihood methods, also with Bonferroni adjustment. To address the secondary objectives, more elaborate logistic regression models will be built for each of the binary outcomes with available individual-, community-, and facility-level factors included as covariates in addition to the intervention status.

#### Nested studies

A series of nested, mixed methods studies will be conducted to identify factors associated with ART engagement within each intervention arm, and to explore the implementation and acceptability of interventions.

##### Qualitative data collection

We will conduct in-depth interviews with a random subset of 120 male participants throughout the study period in order to assess characteristics of men who fail to engage in care, contextualize decisions around ART initiation and retention, and identify additional strategies that may be needed for men to successfully engage and be retained in ART programs.

Interviews will be conducted by a trained male interviewer. Interviews will be digitally recorded, transcribed, and translated into English for analysis. Investigators will pilot a codebook by independently reading and coding a randomly-selected subset of transcripts. Through an iterative consultative process, each investigator will revise their respective codebook until there is high interrater reliability among the group. All transcripts will be coded in Atlas.ti v8.3 using constant comparison, and coding disagreements will be resolved by consensus.

##### Implementation Log Sheet

During the course of the intervention, HCWs will keep daily logs as one of the study monitoring and evaluation tools to assess the implementation of the intervention for each participant. Primary events to be recorded in the daily logs are: (1) unable to reach participant (and reason); (2) contacted participant; (3) intervention provided (and notes about the challenges and successes of the interaction); and (4) other comments relevant to intervention implementation. Each event will be recorded with a corresponding date. Logs will be digitized in English. Findings may influence how similar interventions are implemented in the future.

### Ethics and Dissemination

The IDEaL trial is registered with ClinicalTrials.gov as NCT05137210. The protocol was approved by the Institutional Review Board of the University of California, Los Angeles and the National Health Sciences Research Council in Malawi. Study findings will be disseminated through peer-reviewed journal articles, national and international conference presentations, and meetings with Malawi Ministry of Health, facility, and community stakeholders.

## DISCUSSION

Studies have reported poorer outcomes for HIV testing, treatment initiation, and treatment adherence in men compared to women^2^ for over a decade.^34,35^ Men are often portrayed as difficult, hard-to-reach, and actively avoiding health facilities. In IDEaL, we aim to investigate whether men really are hard-to-reach or, will men engage in care when services are offered in ways that are accessible to them and resonate with their needs, as growing evidence suggests.^12^ We propose to test a Stepped intervention that increases in intensity over time against Low- and High-intensity interventions – all tailored to men – to identify the most cost effective strategy to (re-)engage men in HIV treatment services in Malawi.

IDEaL is different from other ART engagement and re-engagement interventions in several important ways. First, we will enroll men living with HIV across the treatment cascade, including those who have <u>never initiated</u> ART, those who are at risk of <u>immediate default</u> after initiation, and those who have been in care but subsequently <u>default</u>. Formative research suggests that barriers to ART initiation and re-initiation may be similar,^36^ however most interventions focus specifically on either first-time initiation or re-engagement, but not both. Our study will assess if one overarching program can improve men’s engagement across the treatment cascade, regardless of whether they are starting ART for the first time or returning to care after a period of disengagement. One overarching intervention may be more scalable than multiple, separate interventions across the cascade. Second, we tailor interventions to men’s unique needs and motivations, based on extensive formative work. While innovative interventions for men are underway,^19-21^ few have rigorously tested the impact of male-tailored interventions on ART engagement.^37^

Our study will measure adverse events including unwanted disclosure, end of a relationship, and/or intimate partner violence (IPV). We will also measure female partner perceptions of the feasibility and acceptability of a male-only intervention, and any unintended or adverse events such as IPV. Understanding these events will be critical to evaluating the feasibility of scale-up should one or more of the approaches prove effective.

Finally, we will test a Stepped intervention that builds in intensity over time until men (re-) engage in care. This approach allows men who are ready to (re-)engage to do so at minimal cost to the health system, while those who need additional support can receive more resource-intensive interventions to support their ART engagement.^25^ Stepped interventions have been effective in other settings and can address multiple barriers faced by the target population with minimal cost.^25,26^ Findings from IDEaL will provide crucial knowledge to how best men can be reached and can inform intervention scale-up.

## Data Availability

All data used to draft this protocol manuscript are publicly available and can be found in the reference section.

## Contributorship statement

KD and AC conceptualized the study. KD is responsible for funding acquisition. KD, KB, JH, KP, BN, RH and AC developed study protocol and materials. JH, KB, KP, and EC will implement the study. KD, MK, KB, TC, BN, LL, TC, and AC developed the analysis plan and KD, MK, KB, and AC will analyze the data. KD and EC wrote the first draft and KB, JH, KP, BN, LL, RH, SP, EC, RH, TC, and AC edited following drafts. All authors have read and approved the final manuscript.

## Acknowledgements

We wish to thank the Malawi Ministry of Health for their support of this trial. We would also like to acknowledge Joep van Oosterhout, Misheck Mphande, Isabella Robson, Thoko Banda, Peter Mwamlima, and Eric Lungu for their contributions to protocol development and for their support of study implementation, and to Hannah Whitehead for assistance with manuscript development and submission.

