## Supporting Information 2 for "Identifying efficient linkage strategies for HIV self-testing (IDEaL): a study protocol for an individually randomized control trial"

Kathyrn Dovel, Principle Investigator	Partners in Hope	PO Box 302

3-6-2019

**ABSTRACT**

**Background:** HIV self-testing (HIVST) has been found to be a highly acceptable approach for men to learn of their HIV status and has resulted in increased testing uptake (Dovel 2019, cite Augustines stuff). However, rates of antiretroviral therapy (ART) initiation among those tested with HIVST are difficult to capture and some studies have suggested that linkage rates are low (Ortblad 2017, MacPherson 2014), particularly amongst men. We propose a clinical trial to test varying approaches to ART initiation among men who test HIV-positive through HIVST. We will test three interventions:

*Lightest Touch Intervention (Arm 1)*: simple reminders to visit the health facility (given every two weeks);

*Staged Intervention (Arm 2):* a staged intervention that consecutively increases in intensity every month that a participant does not initiate ART (intervals include reminders, motivational interviewing, and home-based ART initiation);

*Intensive Intervention (Arm 3):* home-based ART initiation followed by linkage to the health facility of their choice the following month.

**Objective:** Our primary objective is to identify a cost-effective package for ART initiation among men identified as HIV-positive through HIVST in Malawi. Our specific objectives are:

Objective 1. Evaluate the effectiveness of the Staged ART Intervention vs Lightest Touch Intervention (primary analysis) and the effectiveness of the Staged ART Intervention vs Intensive Intervention (secondary analysis) on ART initiation within 4-months after enrolment in the trial.

Objective 2. Identify individual-, community-, and facility-level factors associated with ART initiation within each intervention arm (Lightest Touch; Staged; and Intensive Interventions).

Objective 3. Determine the cost and scalability of each intervention (Lightest Touch; Staged; and Intensive Interventions).

**Methods:** We will preform an individually randomized control trial with 543 HIV-positive men identified through HIVST and their female partners. Men will be individually randomized 1:1:1 to one of the three intervention arms described above. The study will be preformed at 10 health facilities suppored by Partners in Hope (PIH). Data collection will include baseline and follow-up surveys and interviews with men and women; medical charter reviews at four-months after study enrollment; qualitative interviews; and a cost analysis of costs associated with each arm. Participants will be enrolled in the study for a total of 4 months with approximately 2 or 3 study visits throughout that period.

**Anticipated results**: We anticipate learning about the most effective stragty to engage men in ART. We also anticipate learning about the type and degree of followup necessary to support men’s engagement in ART services. Finally, we anticipate learning about the cost-effectiveness of intervention, with the goal of improving cost-effectiveness for the Ministry of Health. Results from this study could be used to define best practices and to further scale ART-focused programs for men in Malawi.

### 1. INTRODUCTION

#### 1.1. Background

HIV self-testing (HIVST) is an effective strategy to improve HIV testing coverage, especially among hard-to- reach populations such as men and youth. Index testing, whereby a HIV positive client gives an HIVST kit to their sexual partner to use at home, is considered beneficial for its ability to maintain a testers privacy. The method is now recommended by the World Health Organization (WHO) and is being adopted as policy throughout sub-Saharan Africa (SSA). However, uptake of antiretroviral therapy (ART) and adherence after utilizing HIVST remains sub-optimal among certain population, specifically men. Innovative ART initiation and early retention strategies are urgently needed for Index HIVST to be successful.

#### 1.2. Problem statement

We must better understand how to engage men in HIV care. Specifically, there is limited literature on feasible differentiated models to support men to start and stay on treatment that can be taken to scale. Further, there is increased recognition that individuals are at greatest risk of loss-to-follow-up during transition periods across the cascade (i.e. when starting ART). ART initiation and early retention must be improved if HIVST is to become a viable option for high-risk groups in SSA. To address this gap, we propose to conduct a study to test and evaluate varying strategies for ART initiation and retention amongst men.

#### 1.3. Justification

This study will combine HIVST with a second-level intervention focused on ART initiation to address the urgent gap in ART initiation and early retention among HIVST users. Additionlly, Objective 3 will allow us to develop the lowest cost intervention package while reaching the highest number of male partners.

### 2. OBJECTIVES

#### 2.1. Primary objective

***Objective 1.*** Test the impact of a staged ART intervention vs simple reminders and the effectiveness of a staged ART intervention vs home-based ART on ART initiation within 3-months of an HIV-positive diagnosis

#### 2.2. Secondary objectives

***Objective 2.*** Identify individual-, community-, and facility-level factors associated with ART initiation within each intervention.

***Objective 3.*** Determine the cost and scalability of each intervention.

### 3.  LITERATURE REVIEW

***Background***

Men in sub-Saharan Africa are less likely than women to use HIV services.^1^ Men’s absence from care is concerning not only for their own health, but also for the health of girls and young women who continue to be infected at unacceptably high rates.^2^ HIV prevention and treatment programs have not traditionally been directed at men. Men are notably absent from international guidelines, national policies, and local HIV interventions. Research shows that women are 322% more likely to be mentioned in international HIV guidelines than men.^3^ In the context of Malawi, national guidelines expect women of reproductive age to attend a health facility 5-17 times per year (equivalent to 19-63 hours)^4^, and 180-472 times in their reproductive lifespan (15-44 years). There are no such expectations for men (see Table 1). The justification for the global attention of HIV program thus far on women and girls is without dispute. Gender inequality is a key driver which impacts women's health and access to HIV services and creates specific vulnerabilities for women to HIV infection.^5^ However, framing HIV as a woman’s concern means we have failed to understand how gender affects and drives the burden of ill health for men, and inadvertently perpetuates the epidemic for young women and girls. Targeted strategies specific to men are urgently needed if we are to engage them in care.

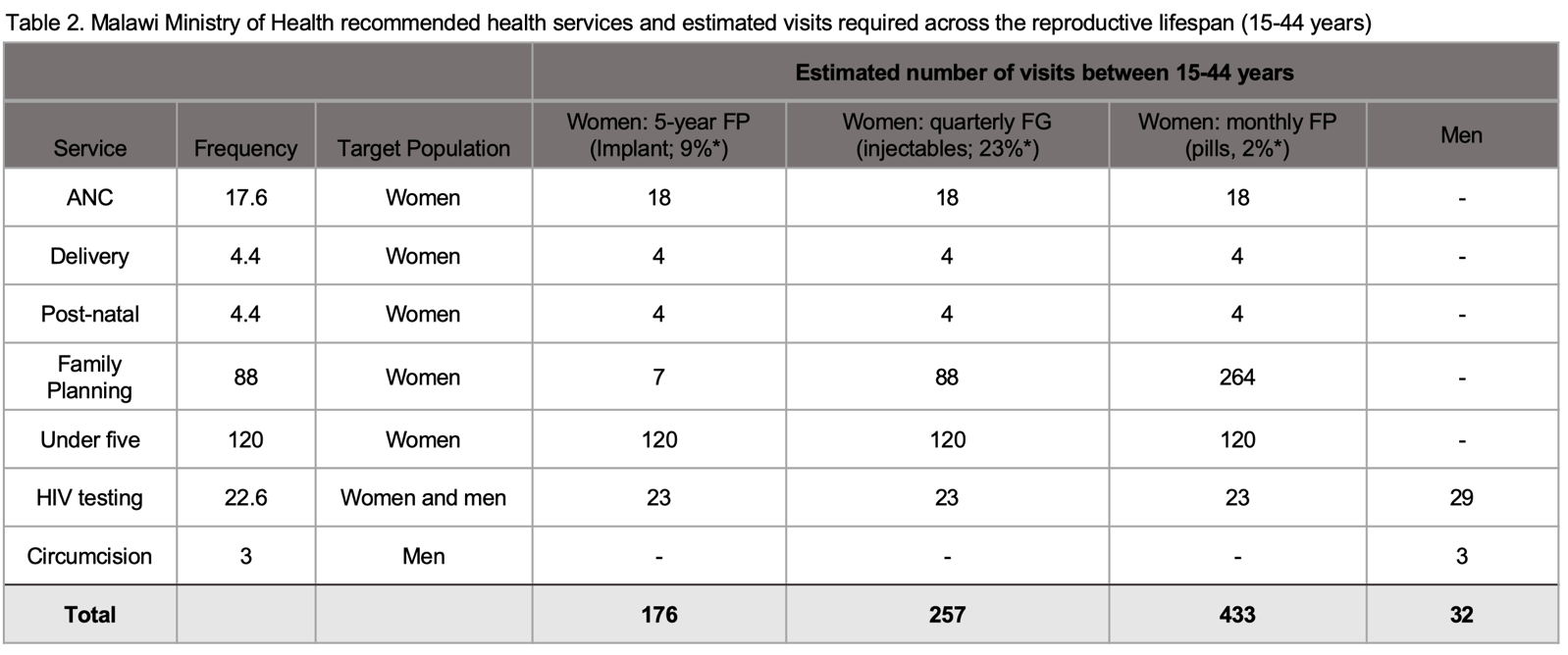
***Table 1:*** *Malawi ministry of health recommended health services and estimated visits required across the reproductive life span (15-44 years)*

Male partners of women who are already identified as HIV-positive (index partners) are still a major concern for epidemic control due to high rates of multiple and concurrent partnerships among men^6^ and the fact that index male partners have two times the risk of being HIV-positive as compared to the general male population.^7^ Data from a recent HIVST study show that there is a high need for index testing among men in Malawi: across 3 high-burden district hospitals in Malawi, men represented 73% of all index partners in need of testing. Among those who did test, male partners were 4 times more likely to test HIV-positive than female partners (23% versus 3%), representing urgent unmet need among men.^8^

A recent Index HIV Trial in Malawi found that HIV testing among men increased dramatically when HIV-positive clients give HIVST kits to their sexual partners to use at home.^9^ The study found that 66% of male partners in the HIVST arm tested for HIV compared to only 22% of men in the standard partner referral slip arm. Within the HIVST arm, men who tested for HIV had an HIV-positivity rate of 23%, with no adverse events reported (see Fig 1).^2^ Index HIVST is highly acceptable and allows men to test at times and locations convenient for them, with complete privacy in their own homes.^10,11^

However, innovative ART initiation and early retention strategies are urgently needed for Index HIVST to be successful. The aforementioned study showed that ART initiation was unacceptably low, with only 22% of HIV-positive men in the HIVST arm initiating ART at 6-months versus 75% of men in standard partner referral slip arm).^3^ (See Figure 1). Poor rates of ART initiation are commonly reported across most HIVST studies, with ART initiation rates ~20-45%^7,12–14^, although ART initiation is notoriously difficult to measure within HIVST strategies. A cost analysis for national scale-up of Index HIVST in Malawi showed that 76% of men tested must initiate ART for Index HIVST to be cost-neutral at the national level as compared to using partner referral slips.

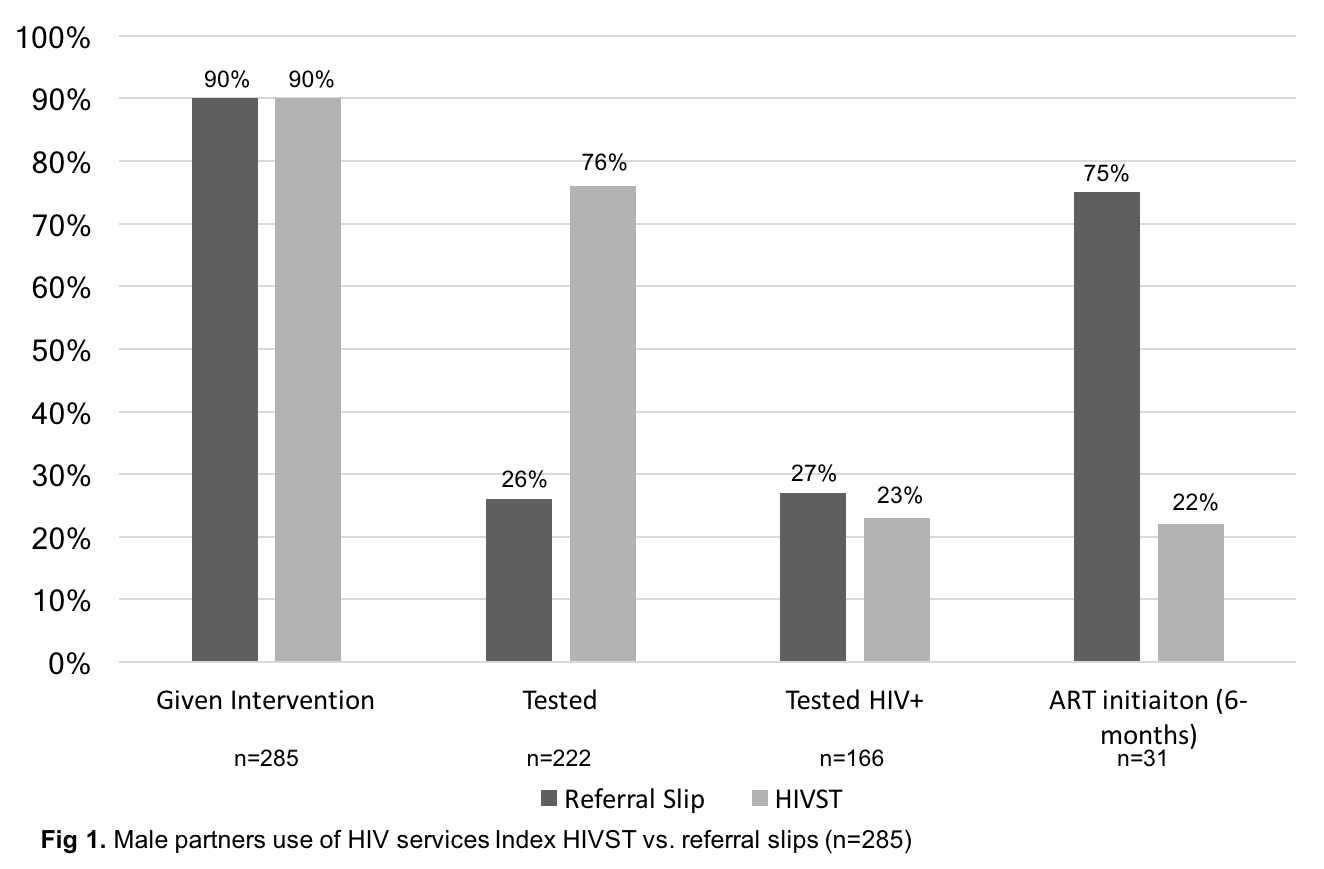
***Figure 1:*** *Male partner use of HIV services Index HIVST vs. referral slips (n=285) from HIVST trial (PI: Dovel)*

Two overarching barriers keep HIV-positive men from accessing ART services: (1) lack of male-friendly services;^15–17^ and (2) harmful gender norms.^18–20^ Male friendly services are private and convenient (requiring minimal time), and offered by health workers who understand the unique needs of men.^21^ In addition, men are often unfamiliar with the health system, and are unsure how to navigate facility-based services. Gender norms that prioritize men as strong and self-reliant perpetuate fear of unwanted disclosure and stigma, and discourage men’s engagement in ART.^18,21^ Our research in Malawi found similar barriers to ART initiation for men who tested HIV-positive: men avoided ART services due to (1) fear of unwanted disclosure and stigma due to lack of privacy; (2) time/cost required to access care; (3) poor knowledge about the benefits of early ART initiation; and (4) beliefs that require men be strong, in control, and focused on short-term benefits such as daily financial earnings and respect from their male friends. Index HIVST must be combined with innovative ART interventions that address these barriers.

***Evidence based for interventions that increase ART initiation***

We have conducted a thorough search of the literature and have identified several intervention strategies that may increase ART initiation among men who use HIVST: (1) reminders + peer navigation; (2) motivational interviewing; and (3) home-based ART.

*Reminders + Peer Navigation* is shown to help clients overcome fears about facility-based services and provide peer modeling how to live successfully with HIV.^22^ While the strategy has been primarily tested within traditional HIV testing strategies, we hypothesize that the same mechanisms will work for men who test through Index HIVST. Reminders are usually done over the phone via phone calls or SMS and can vary in frequency based on the health care workers disgression. Peer Navigation is assisted guidance to the health clinic as well as overviews of where to go/what to do when at the facility once there to ensure men feel more comfortable in the clinic environment.

*Motivational Interviewing* is becoming widely recognized as a key strategy to help clients navigate barriers to the desired outcome by building client’s self-efficacy, identifying internal motivation for the desired behavior, and establishing strategies and short- and long-term goals needed to reach a desired outcome.^8,23^ Motivational interviewing is seen as particularly effective when clients need to make difficult decisions and overcome multi-level barriers to behavior change.^24,25^ The strategy has been used to improve ART adherence^8,25^ and reduce sexual risk behavior.^15^

In contrast, traditional counselling efforts are largely informational and directive, whereby health care workers deliver a pre-determined counseling package that is not responsive to a client’s individual situation.^16,17^ Such methods have been proven largely ineffective,^18^ particularly with hard-to-reach populations such as men.^19^ Motivational interviewing differs from traditional strategies by adopting a client-centered approach is based on collaboration, evocation and respect for autonomy. We hypothesize that these counseling techniques will encourage HIV status acceptance and disclosure, promote health seeking behavior, provide coping strategies men need to overcome barriers related to facility-based care, and ultimately, facilitate ART initiation.

Furthermore, motivational interviewing and client-centered care should resonate with and address the needs of men. Partners in Hope Malawi conducted 25 interviews with men and 6 focus group discussions with health care workers and female partners (n=42) to assess what health services men desired. Overwhelmingly, men reported wanting increased counseling on sexual health (including HIV) and marital concerns. Exit surveys with male ART clients (n=180) show that only 38% of men were aware of Treatment as Prevention and 65% aware of the benefits of early ART initiation, highlighting major knowledge gaps that may influence engagement in care. Motivational interviewing and client-centered counseling will be able to address both gaps in ART treatment and sexual health knowledge.

*Home-based ART initiation* has improved ART initiation across the region. A systematic review found that home-based ART is associated with ART retention, decreased mortality,^26^ and in some cases, reduced stigma and increased privacy.^27,28^ We conducted one of the only studies to examine home-based ART initiation within a community HIVST distribution strategy (Co-I: Choko). We found that home-based ART initiation alongside home-based HIVST significantly increased ART initiation as compared to standard facility-based initiation (RR 2.94; p-value<0.001).^13^

Home-based ART may be particularly attractive to hard-to-reach men because it reduces client time required to access services and provides an easy, opt-out entry point for men who otherwise may have never engaged with the health care system, or know how to navigate complicated, busy health facilities. Home-based ART has been associated with a three-fold reduction in financial costs to clients.^29^ Further, home-based ART facilitates client-centered, one-on-one care that is often not feasible in busy clinic settings.

The Malawi Ministry of Health is in the process of rolling out community-based ART distribution strategies, and may consider home-based ART initiation for hard-to-reach populations. However, home-based ART is considered a resource intensive strategy, and therefore should (1) only be offered to the hardest-to-reach populations, and (2) requires that clients who initiate ART at home-based eventually link into facility-based care. Additionally, findings from a recent Index HIVST Trial show that home-based services for men is acceptable to female ART clients in the Malawian context, with minimal risk of adverse events. Over 90% of female ART clients had disclosed their HIV-status to their partner and were willing to have their male partners traced in their homes for additional services.

Finally, increased privacy and decreased wait-times are essential if men are to engage in HIV services. As part of a study on new Universal Treatment policies in Malawi, 15 in-depth interviews and 208 surveys were conducted with newly diagnosed HIV-positive men. Fear of unwanted disclosure due to limited privacy and a lack of trust in the health facility were the primary barriers to men’s ART initiation. Home-based ART initiation can help address these barriers for ART initiation, and motivational interviewing can help provide men the skills needed to navigate these barriers within the health system in order to promote ART retention. Further, the vast majority (>95%) of men who used HIVST in the Index HIVST trial disclosed their HIV status to their female partner^8^, meaning that home visits (i.e., reminders, peer navigation, motivational interviewing, or home-based ART) will not increase risk of unwanted disclosure to one’s sexual partner. Table 2 below outlines how the proposed interventions will address barriers to ART initiaiton identified in the literature.

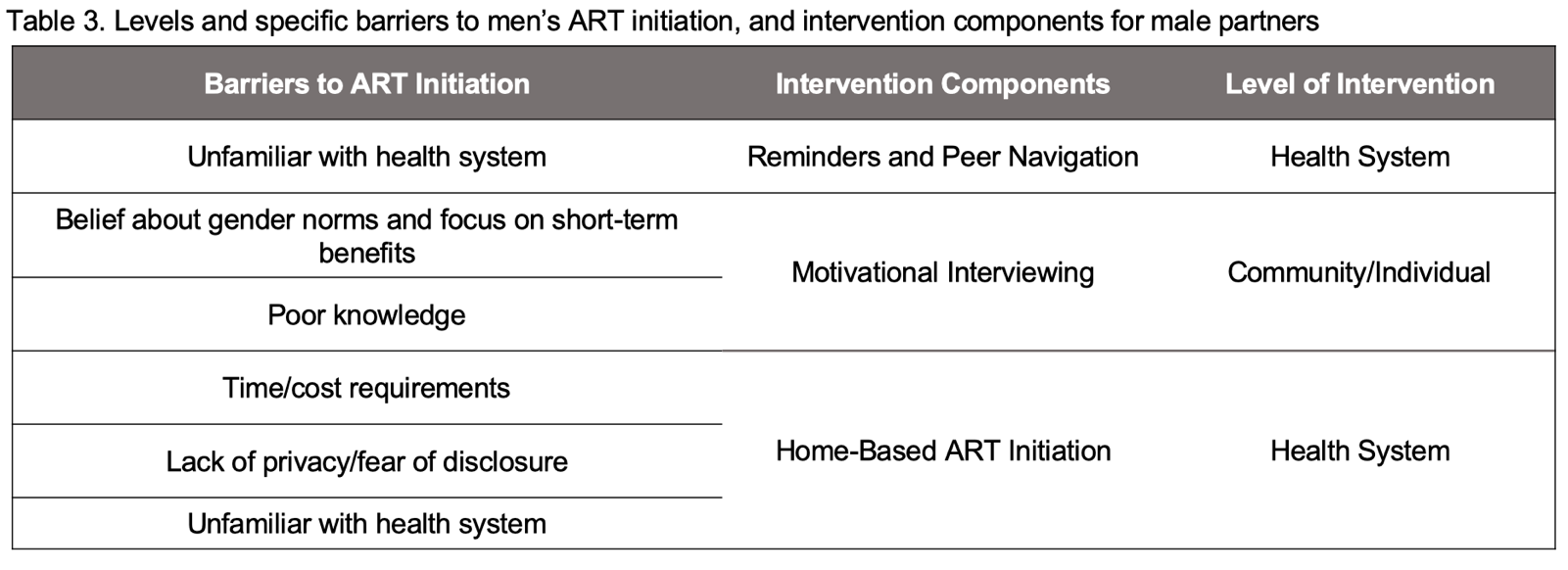

***Table 2.*** *Levels and specific barriers to men’s ART initiation, and intervention components*

### 4. METHODOLOGY

This study will be an individually randomized trial comparing three dfferent strategies to improve ART initiation and early retention among men who test HIV-positive with HIVST. Study staff will utilize the Minsitry of Health Index Testing Register and trace HIV positive women and their male partners to be screened and enrolled if they meet the inclusion criteria. Enrolled men will be randomized to one of three arms and will receive follow up and varying degrees of support based on the arm assigned. Outcomes will be assessed after 90 days after enrollment. Survey data, qualitative data, medical chart data (Objectives 1 and 2) and costing data (Objective 3) will be collected over years 1-3. A study flow chart is illustrated below (Figure 2).

**Figure 2:** *Study flow chart*

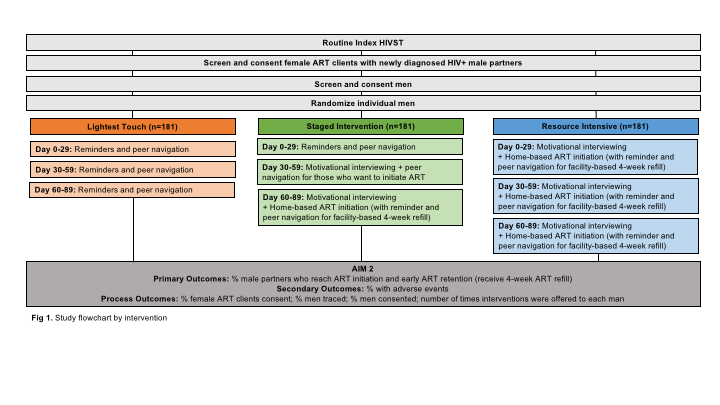

#### 4.1. Intervention description

Each arm will offer an intervention immediately after study enrollment (that same day; day 0). Follow-up interventions will be offered every 14 days after that until 76 days or ART initiation is reached, whichever comes first.

***Description of study arms:***

**Arm 1: “Lightest Touch” Intervention,** whereby facility staff provide reminders and peer navigation (SMS and home visits) for men to encourage enrolment in facility-based ART programs. One reminder on the day of enrollment and every 14 days thereafter, until 76 days or ART initiation, whichever comes first. If initiation is not reached at 90 days, the patient will be classified as not initiated for study purposes. See Arm 1 diagram below:
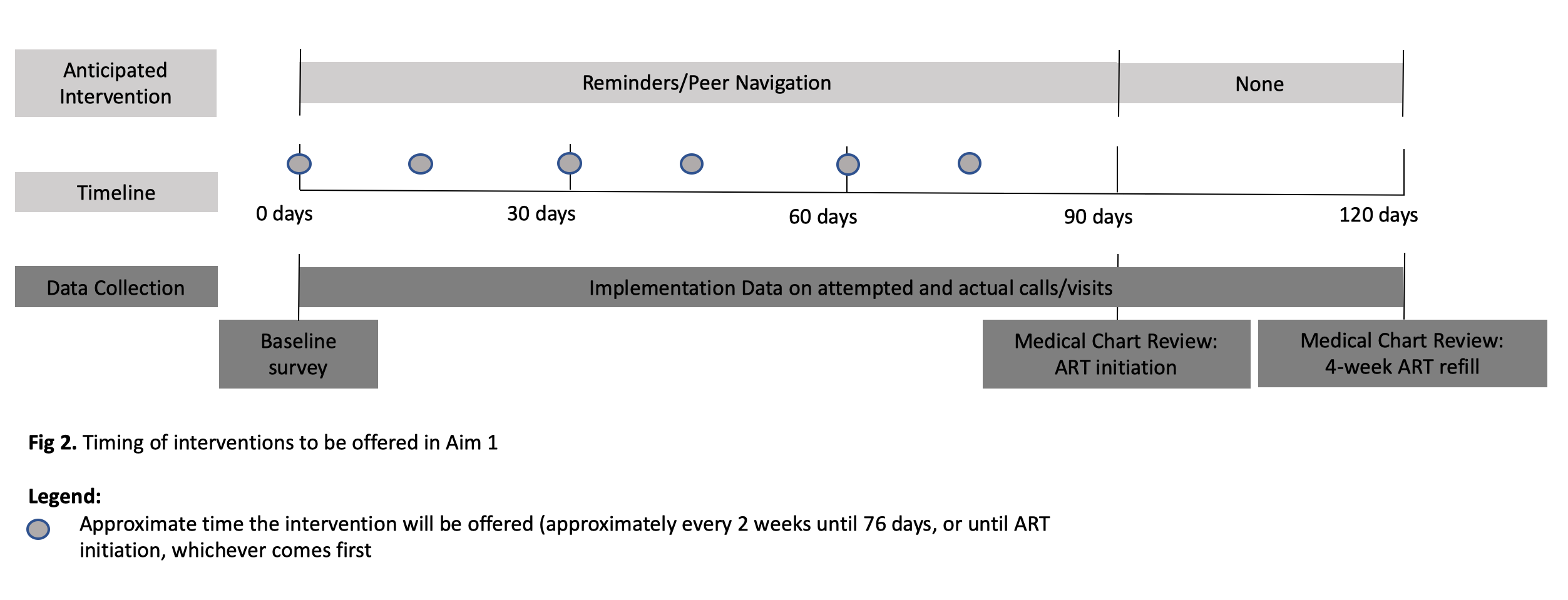

**Arm 2: “Staged” Intervention**, whereby the intervention will build in intensity each month for those who have not initiated ART in the previous month, or for a maximum of 3 months, whichever comes first. The following intervention components that will be added each month (incrementally) until the first ART distribution is completed:

- Day 0-29: Reminders and, for those who agree to initiate, peer navigation;
- Day 30-59: Motivational interviewing and, for those who agree to initiate, peer navigation;
- Day 60-79: Motivational interviewing + home-based ART initiation and, for those who initiate, reminders and peer navigation for the facility-based 4-week ART refill appointment).

See Arm 2 diagram delow:

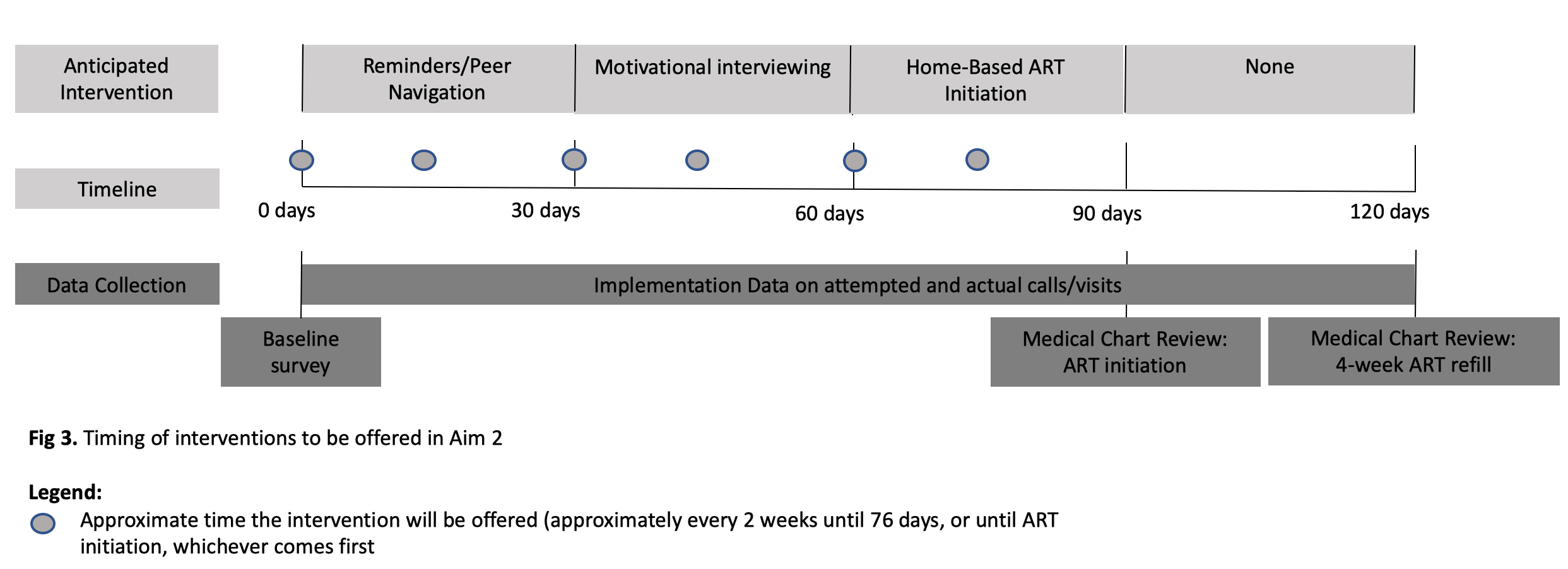

**Arm 3: “Intensive” Intervention**, whereby the most resource intensive intervention is offered immediately to all HIV-positive male partners. See Fig 4 for timeline. Components include:

- Motivational interviewing
- Home-based ART initiation
  - 4-weeks after ART initiation: Reminders and peer navigation to facility for 4-week ART refill appointment

Home-based ART initiation will be scheduled at times convenient for men, including evening and weekend hours. Men who prefer to initiate in another private location in the community (besides their home) will be able to do so. The first home-visit will be conducted by a trained nurse and will include confirmatory HIV testing using Ministry of Health standard algorithm (Determine + Unigold), pre-ART counseling and motivational interviewing, a basic health evaluation, and ART initiation with a 30-day supply of first-line ART in Malawi – dolutegravir, tenofovir, and lamivudine as a single tablet. Clients will also be given a 30-day supply of cotrimoxazole, which is standard of care for all HIV-positive individuals.

Prior to ART initiation, a basic health evaluation will be performed by the nurse, including screening for tuberculosis with routine questions.^66^ Any individual identified by the study nurse with concerns for an active opportunistic infection or other health problem(s) that could complicate home-based ART will be immediately referred and escorted to the facility.

At the same visit, motivational interviewing will be performed in preparation for men to engage in facility-based ART services. This includes counseling on the benefits of early ART, strategies for disclosure and positive living, strategies to overcome facility-based barriers to ART services, and addressing harmful gender norms that may discourage men from using care. Counseling will be adaptive to the needs and concerns of male clients. At 4-weeks after ART initiation, an expert client will escort the man to a nearby facility of his choice to join the facility-based ART cohort. Peer navigation will be provided to ensure men become familiar with the facility-based program. Men who wish to attend a facility that is not nearby will be linked with a male counselor from the selected facility. After completing all facility-based ART services for that day, the male partner will receive additional client-centered counseling with the same counselor to discuss the experience, benefits and challenges associated with facility-based ART, and strategies to overcome barriers. See Arm 3 diagram below:

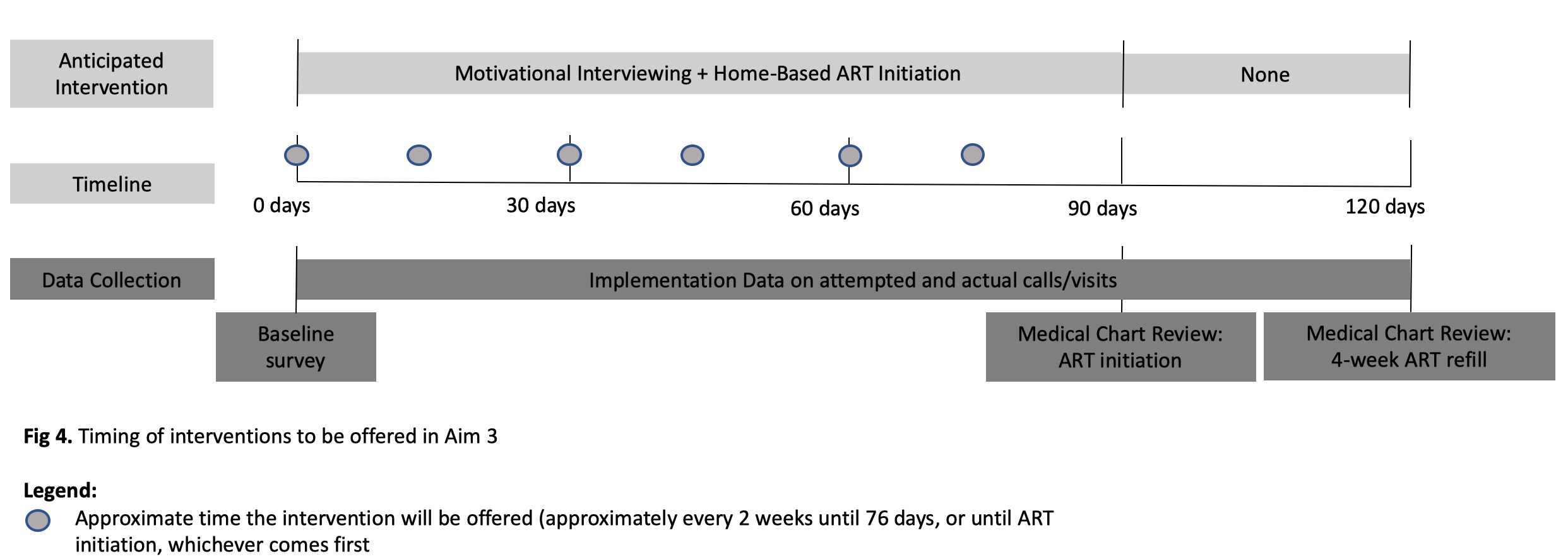

#### 4.2. Place of study

All study activities will take place in 10 Partners in Hope supported facilities within the Lilongwe and Chikwawa districts, representing 19,198 adult female ART clients across all sites. These districts were chosen because they are priority districts for the Presidents Emergency Plan for AIDS Relief (PEPFAR) and have the highest HIV prevalence and unmet need of all Partners in Hope supported districts. See Table 3 for the selected 10 health facilities.

***Table 3:*** *Selected study sites*

| **District** | **Facility name** | **ART Cohort Size** |
| --- | --- | --- |
| Chickwawa | St. Monftord Mission Hospital | 3533 |
| Chickwawa | Kalemba Community Hospital | 2087 |
| Chikwawa | Chickwawa District Hospital | 4898 |
| Kasungu | Kasungu District Hospital | 5942 |
| Lilongwe | Nkhoma Community Hospital | 2166 |
| Lilongwe | Mponela Rural Hospital | 1099 |
| Lilongwe | Daeyang Luke Hospital | 1594 |
| Nkhotakota | Nkhotakota District Hospital | 5361 |
| Nsanje | Nsanje District Hospital | 3904 |
| Nsanje | Ngabu Rural Hospital | 1237 |

Specific methodology for each Objective is described below:

#### 4.3. OBJECTIVE 1

**Evaluate the effectiveness of the Staged ART Intervention vs Lightest Touch Intervention (primary analysis) and the effectiveness of the Staged ART Intervention vs Intensive Intervention (secondary analysis) on ART initiation within 4-months after enrolment in the trial.**

- **Hypothesis 1.1:** 25% of men will initiate ART with simple reminders compared to 45% with motivational interviewing
- **Hypothesis 1.2:** 65% of men will initiate ART with home-based ART initiation compared to 45% with motivational interviewing

##### 4.3.1. Study design

We will conduct an individually randomized controlled trial at 10 high-burden facilities in Malawi.

##### 4.3.2. Target population

We will enroll 543 HIV-positive men identified through routine Index HIVST strategies and their female partners (1,086 participants total). While men are the primary focus of the study, female partners will be enrolled in order to understand their perception of their male partners use of ART services, acceptability of the intervention, and any unintended outcomes or adverse events.

###### *Eligibility Critiera*

Female ART clients will be enrolled in order to conduct baseline and follow-up surveys to understand their perception of their male partners use of ART services, acceptability of the intervention, and any unintended outcomes or adverse events.

*Female Partner*

- Inclusion criteria include: (1) client and partner are ≥15years of age; (2) partner lives in facility catchment area; (3) partner tested HIV-positive and has not initiated ART; and (4) ART client reports no interpersonal violence (IPV) as defined by WHO with their current sexual partner in the past 12 months.
- Exclusion criteria include: (1) client and partner are <15years of age; (2) partner does not live in facility catchment area; (3) partner has not tested HIV-positive or has testing positive and has initiated ART; and (4) ART client has reported interpersonal violence (IPV) as defined by WHO with their current sexual partner in the past 12 months.

###### Men will be enrolled as the primary recipient of the intervention.

###### *Male partner*

- Inclusion criteria include: (1) self and partner ≥15 years of age; (2) live in the facility catchment area (i.e., in the past 30 days, has spent ≥50% of all nights in the village); (3) has tested HIV-positive and has not initiated ART;
- Exclusion criteria include: (1) self and partner <15 years of age; (2) does not live in the facility catchment area (i.e., in the past 30 days, has spent <50% of all nights in the village); (3) has not tested HIV-positive or has tested HIV-positive and has initiated ART

##### 4.3.3. Sampling techniques and enrollment

Sampling, screening, and enrolling male partners will be embedded within routine Index HIVST strategies.

*Brief description of Routine MOH HIVST Guidelines*

Briefly, routine Index HIVST includes three steps:

- *Identify ART clients with sexual partners in need of Index HIVST:* as outlined in the Malawi guidelines within the health care facility.
- *Distribute Index HIVST:* ART clients with a partner of unknown status will receive standard Index HIVST kit. They will be provided a demonstration, counseling, and an overview of risks and benefits.
- *Follow-Up on Index HIVST Use:* During their next ART appointments, ART clients are asked about HIVST distribution, use, and result of the HIVST kit, along with male partner linkage to a health facility for those who tested HIV-positive (i.e., confirmatory HIV testing and ART initiation). Data are documented in the Index HIVST register.

Once routine Index HIVST activities are completed, study recruitment and enrollment will commence. See Figure 3 for a complete description of particiant enrollment and study activities.

***Figure 3:*** *Study screening and enrollment procedures flow chart*

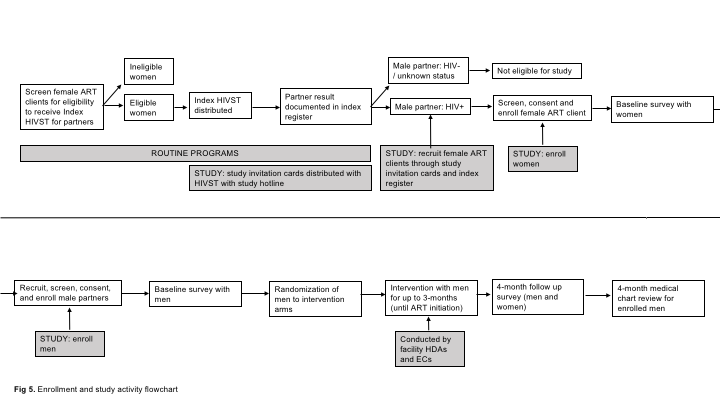

##### 4.3.4. Study Activites

*Female ART client recruitment*

To maximize the number of male partners available for enrollment, the study team will introduce study invitation cards as part of Index HIVST activities and include them with HIVST kits distributed as a part of routine Index HIVST procedures (described above). Study invitation cards will include a ‘hotline’ phone number and will not provide any HIV specific information to ensure confidentiality. (see Appendix A for study invitation card). Female partners will be informed that should their male partner be found HIV-positive, they and their partner may be eligible for a study and should call the hotline number of the invitation card to receive more information.

In addition, the study team will review the Index Testing Register on a regular basis to identify female ART clients who reported HIV-positive male partners through routine Index HIVST procedures. Female ART clients identified will be contact by routine facility staff to be informed about the study and refered to study staff if interested in enrollment.

*Female ART client enrollment*

Female partners interested in the study will provide oral consent to participate in study screening and be screened for eligibility for the study (Appendix B). Study staff will conduct all consent and screening activities. If eligibile and willing to participant, written informed consent will be obtained (Appendix C). Female partners will be enrolled even if their male partner (1) does not consent to study participation (2) cannot be traced.

Upon enrollment in the study, female ART clients will work with study staff to make a plan for inviting male partners to enroll in the study. Baseline surveys with female ART clients will be conducted that same day or on a day convenient for the female ART client. Women whose partners agree to participate in the study will be followed up after 4-months for a follow up survey regarless of the outcome of their partner (ie loss-to-follow-up).

*Male partner recruitment*

Once female ART clients are enrolled, male partners will be recruited, screened, and consented. Study staff will work with the female ART client to establish a recruitment plan that is acceptable and feasible for the women. This may include women referring male partners to the study staff, study staff actively recruiting male partners based on information provided by the female client, or a joint approach where the study staff approaches male partners with the female ART client. Each female ART client enrollled in the study will be allowed to choose the recruitment strategy that best fits her individual situation and the needs of her partner.

*Male partner enrollment*

Men found eligibile and willing to participante, will provide written informed consent (Appendix D) and will be randomized to one of the three study arms. Randomization will occur using an electronic randomization system on tablet devices. A baseline survey will be concuted immediately following consent and the intervention will be carried out over the course of 3 months. Men will be contacted by the study staff 4-months after enrollment for a follow-up survey.

*Randomization*

Randomization will be conducted after completion of the baseline survey with male participants. Study staff will show randomization results as a picture on a pre-programmed tablet, allowing the participant to view the results themselves in order to maximize transparency and study buy-in. Male participants will be randomized to 1 of three arms and will be randomizied 1:1:1.

*Intervention*

Interventions will be offered immediately after study enrollment (that same day or the closest day that is convenient for the male participant). Follow-up interventions will be offered every 14 days after enrollment until 76 days after enrollment in the study or ARTinitiation is reached, whichever comes first. See a full description of the intervention in the Intervention Description section (4.1).

Intervention arms include:

1. ***Lightest touch arm:*** reminders and peer navigation to facility-based ART services
2. ***Staged arm:*** intervention builds in intensity each month for those who have not initiated ART in the previous month. Strategies include reminders and peer navigation, motivational interviewing, and home-based ART initiation
3. ***Intensive arm:*** home-based ART initiation + motivational interviewing + peer navigation to facility-based ART services for their 4-week follow-up appointment

Male expert clients and male nurses will complete all intervention activities to ensure it is as close to real-world implementation as possible (not implemented by Research Assistants). Research Assistants will support facility staff to ensure men are given the appropriate intervention (based on randomization arm), and that all staff activities related to the intervention are documented (i.e., how many reminders were given to each client, ect.). Intervention monitoring and evaluation tools will be developed and incorporated into the facility staff daily routine. Weekly reviews of all intervention monitoring tools and planning for the following week will be completed with study expert clients, nurses, and Research Assistant to ensure adherence to the study protocol. The Study coordinators and PI’s will be highly involved throughout the implementation process to ensure protocol adherence.

##### 4.3.5. Data collection techniques and tools

Data collection tools will include:

- *Baseline Survey:* Research assistants will administer baseline surveys with both female ART clients and male partners immediately following enrollment (before randomization). Surveys will collect data on male and female demographics, sexual partnerships and couple dynamics, and men’s history with health services, and HIV services specifically. (Appendix E & F)
- *Follow-up Survey:* Research assistants will administer follow-up surveys with female ART clients and male participants 4-months after enrolment in the study. Follow-up surveys with men will assess the primary outcome of interest (ART initiation and completion of 4-week follow-up appointment), acceptability of the intervention, and any adverse events (ie., unwanted status disclosure). Men who cannot be reached will be counted as failures for true ART initiation. Follow-up survyes with female ART clients will assess acceptability of the intervention and any adverse events (i.e., IPV, end of the relationship, or unwanted disclosure) associated with intervention procedures. (Appendix G & H)
- *Medical Chart Reviews:* Identifiers will be collected for all men enrolled in the study, including name, age, village and address, and phone number. Identifiers will be used to conduct medical chart reviews at 4-months after enrollment as another measure of ART initiation (attendance to the 4-week follow-up ART appointment). Facility staff (established data clerks employed by Partners in Hope) will review medical records at study facilities and all other Partners in Hope supported facilities within participating districts (61 facilities in total) to account for men who engage in ART outside study facilities. We successfully used this method in other HIVST studies to capture ART initiation.^3^ Male partners who are not found in medical chart reviews will receive a follow-up home-visit to confirm ART outcomes through review of their individual medical record book (health passport) and self-reporting in the event that there are gaps in the record. Men who cannot be reached will be counted as failures for true ART initiation. (Appendix I)
- *Process Implementation Data:* Expert clients, nurses, and research assistants will keep daily logs as part of study monitoring and evaluation tools in order to assess the implementation of the intervention for each participant. Primary events to be recorded in the daily logs are: (1) unable to reach participant (and reason); (2) contacted participant; (3) intervention provided (and notes about the challenges and successes of the interaction; and (4) other comments relevant to intervention implementation. Each event will be recorded with a corresponding date.

Primary and secondary outcomes are measured through medical chart reviews and follow-up surveys (see Table 4).

***Table 4:*** *Study Measures for Objective 1*

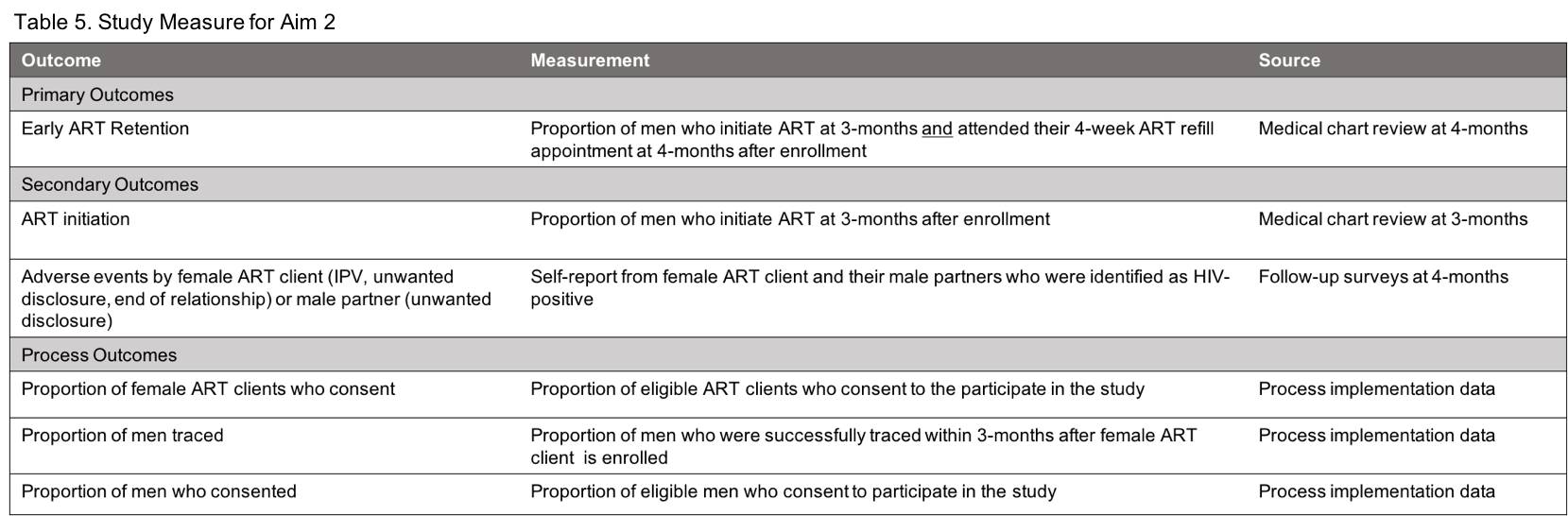

- *Sensitivity analyses for men excluded from the trial***:** We recognize that men’s consent to participate in the study may bias the sample enrolled in the study. There are two groups that we may not be able to include in the main study: (1) men we are unable to trace/contact (herein referred to as “unreachable men”) and (2) men we are able to reach but who refuse to participate in the main trial (herein referred to as “male refusers”). We will take two approaches to address this potential bias
  - Unreachable Men*:* We will collect data on men who are unreachable via their female partner. Female partners for these men will complete a brief survey regarding Surveys will collect data on male and female demographics, sexual partnerships and couple dynamics, and men’s history with health services, and HIV services specifically (as reported by female partners). We successfully used similar methods in the Index HIVST Trial.
  - Male Refusers*:* Men who are contacted but do not consent to the trial will be consented for a one-time survey immediately following refusal for the larger study. The same data will be collected, as described above.

##### 4.3.6. Sample size determination

We powered the study to detect differences in ART initiation between Lightest Touch and Staged Interventions at 4-months after enrollment (primary outcome). We also assured we were powered to detect differences in ART initiation between Staged Interventions and the Intensive Intervention. We Assume that 25% of men in the Lightest Touch arm, 55% in the Staged Intervention arm, and 75% in the Resource Intensive arm initiate ART at 3-months and attend their 4-week follow-up appointment at 4-months. Any man lost to follow-up in any arm will be treated as failures for the outcome evaluation. The sample size needed to detect this difference with the power of 0.8 is 181 men per arm. The calculation is based on asymptotic normality of log odds ratio. We need to enroll and randomize a total of 543 HIV-positive men. Assuming that 25% of women have partners of unknown status, 65% of male partners will use the HIVST kit, 25% of them will be HIV-positive, and 80% of them will enroll in the study, we will need to screen over 3,000 women who were given index HIVST to reach the required sample size.

##### 4.3.7. Data analysis

All randomized men will be included in the analysis of primary outcomes; men with missing outcome assessment due to loss to follow-up will be treated as outcome failures. All primary outcomes are binary; they will be analyzed by logistic regression models with intervention as a predictor, adjusted for baseline socioeconomic and demographic variables. We will conduct sensitivity analyses to account for men who we were never able to contact (unreachable men) and men who refused to participate in the full trial (refusers). We will run several analyses whereby the denominator includes (1) unreachable men and refusers; and (2) refusers.

#### 4.4. OBJECTIVE 2

**Identify individual-, community-, and facility-level factors associated with ART initiation within each intervention arm (Lightest Touch; Staged; and Intensive Interventions).**

- **Hypothesis 2.1***:* In quanitative data, older men, men without strong social support networks, men with high levels of internalized and perceived HIV-related stigma, and men who hold rigid beliefs of gender norms and men’s role as the provider and decision maker of the home will be less likely to initate ART.
- **Hypothesis 2.2***:* In qualitative data, primary factors influencing men’s decision to start ART will be perceptions of feeling healthy, perceptions of one’s ability to continue working and providing for their family without ART initiation, and perceptions of HIV-related stigma within one’s community.

##### 4.4.1. Study design

We will use baseline survey data from the randomized trial (Objective 1) to identify factors associated with ART initiation among men. We will also conduct 200 semi-structured in-depth qualitative interviews with a random sub-set of enrolled men (n=100) and their female partners (n=100) to assess in-depth characteristics of men who fail to engage in care, contextualize decisions around ART initiation and retention, and understand additional strategies that may be needed for male partners to successfully initiate and be retained in ART programs.

##### 4.4.2. Target population

All men and women enrolled in the overarching trial will compelte a baseline survey. Eligibility criteria for study enrollment is described in detail under Objective 1.

A subset of men and women enrolled in the overarching trial will be randomly selected to complete an in-depth interview. Eligibility criteria for in-depth interview are as follows:

*Male partners*

- Inclusion criteria include: (1) randomly selected using electronic random selection techniques; (2) linked to care within 4-months after enrolling in the study (defined as completing the 4-week ART refill appointment) (n=50 respondetns); or (3) did not link to care within 4 months (n=50 respondents)
- Exclusion criteria include: (1) not randomly selected using electronic random selection techniques

*Female partners*

- Inclusion criteria include: (1) randomly selected using electronic random selection techniques; (2) partners could never be traced for study enrollment; or (3) partners were enrolled but were lost to follow up and unable to be reached again
- Exclusion criteria include: (1) not randomly selected using electronic random selection techniques; (2) partners could be traced for study enrollment; or (3) partners were enrolled and were not lost to follow up and were able to be reached again

##### 4.4.3. Sampling techniques and tools

Survey data will include all men and women enrolled in the study (n=1,086; described in detail in Objective 1). 100 men (~ 33 per study arm) and 100 women (~ 33 per study arm) enrolled in the study will be randomly be selected for in-depth interviews.

##### 4.4.4. Data collection techniques and tools

**Surveys:** Baseline and 4-month follow-up surveys will be conducted with men and women enrolled in the trial. They will focus on:

- *Health care system*: perceptions regarding the following aspects of ART services (i) privacy and confidentiality; (ii) availability of services; (iii) wait-time and distance to facility; (iv) quality of care and rude behavior from health care providers, using validated measures.
- *Sociodemographics*: (i) age; (ii) household assets; (iii) work; and (iv) substance use
- *Couple characteristics*: (i) relationship type and length; (ii) sexual activity and risk; (iii) frequency of communication; (iv) disclosure; (v) joint decision making using standard measures from Demographic Health Survey (DHS);^17^ (vi) gender norms using validated Gender Equitable Men (GEM) Scale,^67^ and (v) Revised Conflict Tactics Scale.^68^
- *Knowledge/perceptions and biomedical factors:* (i) knowledge about HIV and ART (treatment as prevention, benefits of early ART); (ii) risk perception (morbidity and mortality); standard DHS measures on (iii) previous use of HIV services and (iv) self-rated health;^17^ and (v) WHO staging at enrollment.

**In-depth interviews:** Table 5 below describes in-depth interview participants and justification. In-depth interview guides for men and women are developed based on existing literature and our extensive experience conducting in-depth interviews with this population (Appendix J and K). Female partners will provide important insight into the circumstances of these men, and potential strategies to more effectively reach them. Men’s qualitative feedback is particularly important for the staged intervention and intensive intervention since these are fairly novel and under explored.

***Table 5.*** *Description of in-depth interview participants and justification*

| **Participant type** | **Number of interviews** | **Justification** |
| --- | --- | --- |
| Women whose male partners were **unreachable** during the study enrollment | 50 (~ 16 per arm) | To understand the couple dynamics and characheristics of men who were never treaced and additional strategies to reaching these men |
| Women whose male partners were **loss-to-follow-up after** study enrollment | 50 (~ 16 per arm) | To understand the couple dynamics and charactersitcs of men who were loss-to-follow-up, and additional strategies to better engage these men in care |
| Male particiapnts who **did not** complete 4-week ART refill appointments by 4-months | 50 (~ 16 per arm) | To understand what they liked and did not like about the intervention, why they did not link to ART, and suggestions on how to improve the intervention |
| Male participants who completed 4-week ART refill appointments by 4-months | 50 (~ 16 per arm) | To understand what they liked and did not like about the intervention, why they linked to ART, and suggestions on how to improve the intervention |

##### 4.4.5. Sample size determination

Survey sample size details provided in Objective 1. The number of interviews required for qualitative data can be challenging to predict. Data should be collected until saturation is reached, meaning that no new themes or relevant information is emerging. The exact number of interviews required to reach saturation differs based on the aim of the study, the diversity in respondents, and the theoretical framework used for analysis.^30^ However, a basic rule of thumb is that no sample size should be under 25 participants in order to reach saturation and identify all relevant themes or new information important to the study.

##### 4.4.6. Data analysis

**Surveys:** Subjects with complete data in outcomes as well as predictors will be included in the analysis. We will calculate descriptive statistics, including mean/median, variation (standard deviation, kurtosis), range, and frequency distributions for the demographic and clinical characteristics, overall and by study arm. Logistic models will be developed for the probability of a positive outcome, with sociodemographic factors included as covariates in a suitable form (linear/spline/factor). Differences in the prevalence of each of the outcomes of interest will be examined by study arm as well as by other factors of interest including demographic characteristics (e.g., age), couple chararisterics, and knowledge/perceptions and biomedical knowledge. The differences will be evaluated using t-tests, Mann-Whitney U test (or other non-parametric tests), chi-square methods, and Fisher’s exact test as appropriate.

**In-depth interviews:** Audio recordings of in-depth interviews will be transcribed and translated to English. A preliminary codebook will be developed for both interview types (male and female). Selected investigators will piloted a codebook by independently reading and coding a randomly-selected subset of transcripts. Through an iterative consultative process, each investigator will revised their respective codebook and repeated this process until there was high interrater reliability among the group. All transcripts will be coded in Atlas.ti v8.3 using constant comparison, and coding disagreements were resolved by consensus.

#### 4.5. **OBJECTIVE 3**

**Determine the cost-effectiveness and scalability of the intervention arms through costing and mathematical modeling.**

- **Hypothesis 3.1:** The staged intervention will be more cost effective at having men initiate ART than both the lightest touch intervention and the intensive intervention.

##### 4.5.1. Study design

We will conduct an incremental cost-effectiveness analysis and mathematical modelling to determine national scale-up potential. The average cost per successful outcome (early ART retention) will be calculated and compared across arms incrementally.

##### 4.5.2. Data collection techniques and tools

Costs will be measured from the health care provider. We will use micro-costing methods by first creating an inventory of all the resources used to achieve the observed study outcomes including:

- Standard counseling interactions (staff cadre, training received, duration of interaction and distance from facility travelled where applicable)
- Motivational interviewing interactions (staff cadre, training received, duration of interaction and distance from facility travelled where applicable)
- Provider interactions (staff cadre, training received, duration of interaction and distance from facility travelled where applicable)
- Cost of reminder messages sent (when messages delivered telephonically instead of in person)

For each study patient, the quantity (number of units) of resources used will be determined. Unit costs of resources, which are not human subject data, will be obtained from external suppliers and the site’s finance and procurement records and multiplied by the resource usage data to provide an average cost per study patient across centers in each study arm.

##### 4.5.3. Data analysis

**Cost-Effectiveness:** Using the average cost per patient as described above, we will then estimate the cost per outcome achieved in each arm. The main measure of effectiveness for the cost-effectiveness analysis will be both the primary study outcome (early ART retention). We will calculate the difference in cost divided by the difference in effectiveness among study arms. Costs will be reported as means (standard deviations) and medians (IQRs) in USD, using the exchange rate prevailing during the follow up period.

**National scale-up modeling**: To determine the budget impact and affordability of the intervention arms, we will parameterize a national scale-up model using the study output. To determine the total cost and impact of the three intervention arms, as well as combinations of interventions, we will model cost and impact out to early ART retention (ART initiation and completion of the 4-week ART refill appointment). The following parameters to be estimated from this trial include:

- Percent of men not linking after HIVST (and thus eligible for this trial)
- Proportion of men that have not linked that could be reached
- Proportion of men that are known HIV-positive and on ART (not disclosed to their partner)
- Proportion of men that initiate ART
- Proportion of men that complete the 4-week ART refill appointment

We will then estimate the expected increase in the number of men linked to ART after index HIVST, adjusted by facility type where possible, by each intervention arm. The number of facility-level HIV tests conducted through index testing at all 652 public healthcare facilities in Malawi from Oct 2019-Sept 2020 will be used for these national calculations. Each intervention will be tested separately in this model, as well as different combination of interventions. Different scenarios will be explored where interventions are used at different facilities (urban versus rural targeting of interventions, geospatial targeting of interventions), or different groups of men within the same facility (where data suggest that different demographics of men resnd differently to the different interventions).

The national-level costs and expected number of men linked to ART, by each intervention and combinations of interventions, will be reported from this model. We will then contextualize the national cost of each intervention with a short-term 3-year budget impact: percent increase (or decrease) of the national HIV treatment budget with the inclusion of one of these interventions

### 5. ETHICAL CONSIDERATIONS

There is minimal risk associated with the above-mentioned procedures. We have extensive experience measuring ART initiation within HIVST studies. We conducted the first trials in the region to objectively measure ART initiation among men after receiving HIVST through the Index HIVST Trial (PI: Dovel) and PASTAL Trial (male partners of antenatal clients; PI: Choko). We draw from lessons learned from our previous trials.

*Informed Consent*

Informed consent will be obtained before any study-specific procedures are performed. The informed consent process will include information exchange, detailed discussion, and assessment of understanding of all required elements of informed consent, including the potential risks, benefits, and alternatives to study participation. The process will emphasize the randomized nature of the study and the differences that participants may experience as part of the study relative to current local standards of care. The study will include children 15 years of age and older. Following Malawian protocol, adolescents <18 years of age will be required to attain assent before completing the survey. Based on prior studies, we anticipate <10% of participants to be under 18 years of age, providing a small sample size to explore the potential impact of facility-based testing for youth.

*Potential Benefits*

Men who participate in the study may have access to additional HIV services not usually provided through routine care, such as appointment reminders, peer navigation, motivational interviewing, and home-based ART initiation. Men can refuse these additional services at any point. Further, both men and women will have the opportunity to discuss their use of HIV services and any concerns with HIV as individuals or as a couple. Information learned in this study may be of benefit to participants and others in the future, particularly information that may lead to optimized testing guidelines.

*Potential risks and discomforts*

Study procedures have minimal risk to the client. For men, maintaining privacy and confidentiality is a potential risk, particularly with home-based ART initiation. In our prior work delivering routine Index HIVST, we have had health workers visit a cluster of homes (not just one) to avoid unwanted questions about the individual’s serostatus. This has worked quite well, with no reports of unwanted disclosure, and we will use this approach in our proposed study to minimize risk of unwanted disclosure. Further, men may refuse any ART service at anypoint if they are uncomfortable.

For women, increased intimate partner violence (IPV) may be a potential risk, particularly if their male partner is prone to violence. To reduse these risks, women who report IPV with their current partner in the past 12 months will be excluded from the study. Female ART clients who report IPV at anypoint of the intervention will be withdrawn from the study, along with their male partner, counseled, and referred to community-based resources for IPV. We also will provide extensive counseling on status disclosure and an IPV hotline to all female participants. Further, Our PASTAL and Index HIVST Trials show no sign of increased IPV and we have published extensively on risk factors for IPV in other settings.^71-73^

Finally, Participation includes completion of a survey that will assess previous use of health services, perceptions of health services received, and sociodemographic and biomedical factors that may be associated with health service utilization. Participants may feel some psychological stress or discomfort from some of the questions, although most questions are not sensitive in nature. Participants may decline to answer any questions that make them uncomfortable and may end participation at any time.

Reimbursement/compensation

Participants will be provided MK 7,500 (equivalent to 10USD) for each survey completed (MK 15,000 / 20USD across the duration of the study). They will receive the above compensation regardless if they use HIV services or not. Those who complete the additional in-depth interview 6-months after study enrollment will receive an additional MK 7,500 (equivalent to 10USD) for their time.

Privacy and confidentiality

All study procedures will be conducted in private, and every effort will be made to protect participant privacy and confidentiality to the extent possible. Participant information will not be released without written permission to do so except as necessary for review, monitoring, and/or auditing. All study-related information will be stored securely. Participant research records will be stored in locked areas with access limited to study staff. All study data will be identified by participant ID (PID) only. Likewise, communications between study staff and protocol team members regarding individual participants will identify participants by PID only. Process evaluation documents, such as intervention monitoring and evaluation tools, will only include PID and will not store PID and identifiers together. All local databases will be encrypted and secured with password-protected access systems. Lists, logbooks, appointment books, and any other documents that link PID numbers to personal identifying information will be stored in a separate, locked location in an area with limited access. For the intervention, home visits will be conducted by health workers who visit a cluster of homes (not just one) at one time in order to avoid unwanted questions about the individual’s serostatus. This has been used in other interventions focused on partner testing and treatment with high success of removing unwanted disclosure to community members.

### 6. DISSMINATION OF RESULTS

This study will set the stage for interventions that combine HIVST with differentiated models for early ART retention in low-resource settings. The study is timely and of high-impact. Findings will establish the effectiveness of home-based ART among male HIVST users, and can directly inform HIV programs throughout the region. The dissemination plan was developed to achieve the most impact while still ensuring dissemination among local stakeholders who may immediately benefit from study findings.

Partners in Hope is already integrated into national technical working groups, so dissemination will follow standard meeting schedules and draw upon Partners in Hope’s longstanding history with the Ministry of Health. Additionally, we will disseminate results through presentations at international scientific meetings and through high-impact peer-reviewed journals. The mentorship team has extensive experience publishing in high-impact journals (e.g., *AJPH, AIDS, BMJ, Lancet HIV, JAIDS*, *PLOS Med*)

### 7. PERSONNEL ROLES AND INSTITUTIONS

The proposed research team includes clinical researchers and implimentation science professionals with substantial experience in HIV testing, HIV prevention and treatment, cost effectiveness, differentiated care model studies, and male-focused studies and programs in Malawi and Sub-Saharan Africa. The study will be implemented in partnership with Partners in Hope Medical Center in Lilongwe, which has years of experience collaborating with Ministry of Health and local health facilities on similar studies, mentoring staff, and running studies embedded within routine clinical care.

- Kathryn Dovel, MPH, PhD, Principle Investigator, Division of Infectious Disease University of California Los Angeles (UCLA) and Research Director for Partners in Hope
- Thomas Coates, PhD, Co-Investigator, Division of Infectious Disease UCLA
- Risa Hoffman, MPH, MD, Co-Investigator, Division of Infectious Disease UCLA
- Brooke Nichols, Co-Investigator, School of Global Health, Boston Univeristy
- Lawrence Long, Co-Investigator, School of Global Health, Boston Univeristy
- Alemayehu Amberbir, Co-Investigator, Partners in Hope
- Augustine Choko, PhD, Site Co-Investigator, Malawi Liverpool Wellcome Trust
- Michal Kulich, Biostatistician, Charles Univeristy in Prauge
- Julie Hubabrd, MSc, Study Coordinator, Partners in Hope
- Kelvin Balakasi, Study Data Manager, Parners in Hope
- Khumbo Phiri, Implimentation Science Manager, Partners in Hope

Dr. Kathryn Dovel, the Principle Investigator, is the Science Director at Partners in Hope and an Assitant Assistant Professor in the Division of Infectious Diseases at UCLA. Dr. Dovel has over ten years of experience in Malawi and collaborating with the study team. She is regularly involved in UNAIDS and WHO workshops and meetings regarding strategies for male engagement, and has been a consultant on two Ministry of Health guidelines on the topic in Malawi.

Dr. Augustine Choko will be responsible with Dr. Dovel for overall adherence to the study protocol and serve as the primary liaison with the local IRB and key stakeholders in Malawi. Dr. Thomas Coates will serve as the community-based trials specialist, with over two decades of experience conducting individual- and cluster-randomized trials in communities with the end goal of engagement in HIV services. Dr. Risa Hoffman is an established clinical investigator and will serve as the MD specializing in differentiated models of ART treatment delivery and HIV care, and ensuring client safety. Brooke Nichols and Lawrence Long will be responsible for reviewing all modeling data, making an analysis plan for the proposed models, and providing modeling for publications. Dr. Michal Kulich is the Chair of the Probability and Statistics Department at Charles University and has extensive experience with the design, conduct, and analysis of clinical trials in the context of HIV prevention research.

Partners in Hope’s staff Kelvin Balakasi (Data Manager) Julie Hubbard (Research Coordinator), Khumbo Phiri (Implimentation Science Manager) and Alemayehu Amberbir (Science Director) will be responsible implimentation and oversight inlcuidng data collection, data management, quality control, and training and certification of data entry personnel. They will also be responsible for ensuring the intervention promotes client safety, meets Ministry of Health guidelines, and is implemented in such a way to promote sustainability and scalability.

CV’s for participating personelle are provided in the Appendix L.

### 8. REGULATORY OVERSIGHT

This study is sponsored by the Bill and Melinda Gates Foundation and implemented through Partners in Hope (PIH), Malawi. PIH staff will perform monitoring visits. As part of these visits, monitors will inspect study-related documentation to ensure compliance with all applicable regulatory requirements.

All health facilities will receive an Initial Registration Notification from PIH that indicates successful completion of the protocol registration process. A copy of the Initial Registration Notification will be retained in the site's regulatory files.

We have developed a trial advisory group. See Table 6 for details about the group members. The group will meet every quarter to review progress, and challenges with study implementation, and provide input on the final interventions to be tested, based on qualitative findings in Aim 1.

***Table 6.*** *Description of trial advisory group*

| **Name** | **Affiliation** | **Expertise** |
| --- | --- | --- |
| Dr. Morna Cornell | University of Cape Town | Epidemiologist, health system barriers to men’s care, men’s HIV services, advocacy and policy change |
| Dr. Heidi van Rooyen | SA Human Sciences Research Council | Social scientist, HIV vulnerability and inequality, interventions for men’s ART initiation |
| Dr. Deborah Donnell | University of Washington, Fred Hutch Vaccine and Infectious Disease Division | Biostatistician, international HIV trials, PI of the HPTN Statistical and Data Management Center |
| Dr. Connie Celum | University of Washington | Infectious disease physician and epidemiologist, implementation science in Africa, HIV prevention trials |
| Dr. Thoko Kalua | Malawi Ministry of Health, Deputy Director at Department of HIV and AIDS | Epidemiologist. Extensive experience in national HIV programs, M&E, and scale-up of interventions on the ground |
| Dr. Sergio Chicumbe | Mozambique National Health Institute (INS), Health System Research Cluster | Clinical trials and implementation science. Extensive experience in national public health programs, methodology for health services research and quality care improvement. |

For any future protocol amendments, upon receiving final IRB/EC and any other applicable regulatory entity approvals, sites should implement the amendment immediately. Sites are required to submit an amendment registration packet to the PIH Protocol Team. PIH key personnel will review the submitted protocol registration packet to ensure that all the required documents have been received.

### 9. STUDY IMPLEMENTATION

Study implementation at each site will be guided site-specific standard operating procedures (SOPs). These SOPs will be updated and/or supplemented as needed to describe roles, responsibilities, and procedures for this study.

### 10.  PROTOCOL DEVIATION REPORTING

All protocol deviations will be documented in participant research records. Reasons for the deviations and corrective and preventive actions taken in response to the deviations will also be documented. Deviations will be reported to site IRBs/ECs and other applicable review bodies in accordance with the policies and procedures of these review bodies. Serious deviations that are associated with increased risk to one or more study participants and/or significant impacts on the integrity of study data must also be reported to the Protocol Team as soon as possible.

### 11.  WORK PLAN TIMELINE

***Table 7:*** *Anticipated workplan timeline of study activities, by year*

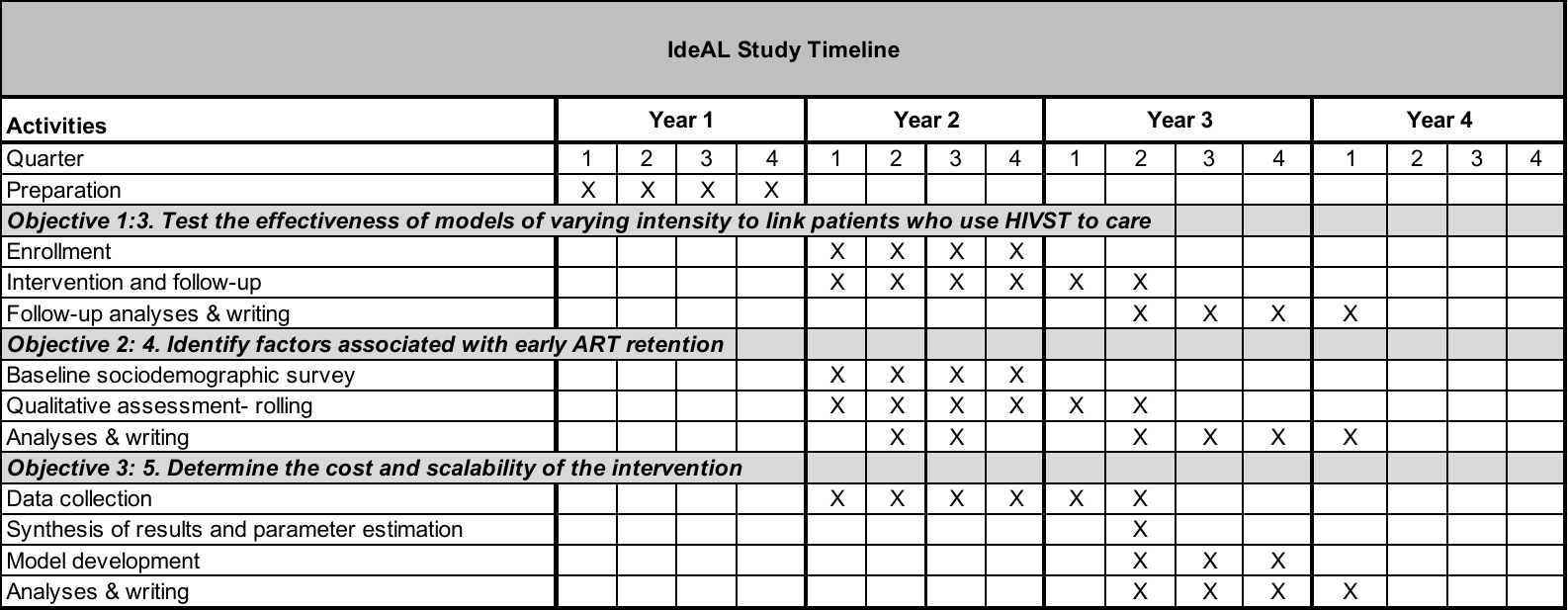

### 12. BUDGET AND JUSTIFICATION

***Table 8:*** *Study budget*

| **Description** | **USD** | **Justification** |
| --- | --- | --- |
| Study Coordinator | 5000 | 25% LOE to coordinate RAs |
| Research Assistant for data collection | 15000 | 5 RAs for 6 months at 500 USD per month |
| Incentive for participants | 21720 | 10USD per study visit, 2 study visits per participant, 1086 participants (543 men and 543 female partners) |
| Expert Client/Nurse lunch allowance | 720 | 6USD lunch allowance for 10 Expert Clients and 10 nurses on a monthly basis to hear from them how the intervention is going |
| Telecomunications | 1000 | Mobile data collection processing by RAs and communication with coordinator |
| NHSRC application fee | 150 | Application fee |
| *Sub Total* | *43,590* |  |
| NHSRC 10% fee | 4,359 | 10% contribution fee of study budget |
| **Grand total** | **47,949** |  |

### 14. APPENDIX

#### 14.1. APPENDIX A – Study Invitation Cards

**STUDY INVITATION CARD**

**Identifying efficient linkage strategies for HIV self-testing (IDEaL)**

English

**Study invitation Card**

Date: ……/……/2020

Dear: __________________________________________

At ____________________________________, we are conducting a study. The study will generate information that will help the Malawi government, Partners in Hope and other stakeholders develop programs that will help your community lead a healthy life. We are therefore inviting you and your partner to come to ____________________________ on____________________________ at_________, so that we can assess if you are eligible for the study. If eligible, you and your partner will each receive MK 7,500 (equivalent to 10USD).

Thank you for your kind attention to this request and we look forward to seeing you. When coming call or flash this number…………………… so that you are met by study personnel.

Sincerely,

[District Health Officer]

#### 14.2. APPENDIX B – Recruitment and Screening Script

**RECRUITMENT AND SCREENING SCRIPT**

**Identifying efficient linkage strategies for HIV self-testing (IDEaL)**

Female

Thank you for speaking with me about the study entitled, *“Identifying efficient linkages strategies for HIVST****”*** conducted by Partners in Hope and the University of California Los Angeles in the United States. You are being approached because you recently reported that your male partner tested HIV-positive using a HIV self-test kit.

The purpose of this is to determine what are the best interventions that can help men who are diagnosed with HIV use other health services, if desired. The study will offer several different strategies for HIV services to see what works best for men who use HIV self-testing kits.

Would you be interested in participating in the eligibility screening to see if you are eligible to participate in the study? Your participation is voluntary and you will not be penalized if you choose not to participate in the screening or the project.

[*If no, thank the person and end the session*]

[*If yes, continue to the screening questions below or make an appointment to complete the screening questions]*

Before we continue with the study, we first need to determine if you are eligible to participate.

1. Are you and your partner 15 years old or older?
2. To the best of your knowledge, did your partner recently test HIV-positive using a HIV self-test kit?
3. To the best of your knowledge, your partner currently NOT taking ART?
4. Does your partner live in the facility catchment area?
5. In the last 30 days, your current partner has NEVER hit, slapped, or kicked you, or forced you to have sexual intercourse with them?

If you answered yes to all these questions, then you are eligible to participate in the study. You may stay here to continue with the study consent, and we will explain how the study will be conducted.

**RECRUITMENT AND SCREENING SCRIPT**

**Identifying efficient linkage strategies for HIV self-testing (IDEaL)**

Male

Thank you for speaking with me about the study entitled, *“Identifying efficient linkages strategies for HIVST****”*** conducted by Partners in Hope and the University of California Los Angeles in the United States. You are being approached because you recently reported testing HIV-positive using a HIV self-test kit.

The purpose of this is to determine what are the best interventions that can help men who are diagnosed with HIV use other health services, if desired. The study will offer several different strategies for HIV services to see what works best for men who use HIV self-testing kits.

Would you be interested in participating in the eligibility screening to see if you are eligible to participate in the study? Your participation is voluntary and you will not be penalized if you choose not to participate in the screening or the project.

[*If no, thank the person and end the session*]

[*If yes, continue to the screening questions below or make an appointment to complete the screening questions]*

Before we continue with the study, we first need to determine if you are eligible to participate.

1. Are you 15 years old or older?
2. You recently test HIV-positive using a HIV self-test kit?
3. You are NOT currently taking ART?
4. Do you live in the facility catchment area?

If you answered yes to all these questions, then you are eligible to participate in the study. You may stay here to continue with the study consent, and we will explain how the study will be conducted.

#### 14.3. APPENDIX C – Written Informed Consent- Female

**WRITTEN INFORMED CONSENT**

**Identifying efficient linkage strategies for HIV self-testing (IDEaL)**

Female

You are asked to participate in a research study entitled *“Identifying efficient linkages strategies for HIVST****”*** conducted by Partners in Hope and the University of California Los Angeles in the United States. You are being requested to take part in the study because you recently reported that your male partner tested HIV-positive using a HIV self-test kit. Your participation in this study is entirely voluntary. You will be read the information below, and you are free to ask questions about anything you do not understand, before deciding whether or not to participate. ***I as the field assistant for this study will take you through this consenting process.***

**Why is this study being done?**

HIV self-testing is very helpful for people who want to know their status but do not usually go to the health facility. However, it can be hard for individuals who test HIV-positive with HIV self-testing to be able to access other health services. Researchers want to determine what are the best interventions that can help men who are diagnosed with HIV use other health services, if desired. The study will offer several different strategies for HIV services to see what works best for men who use HIV self-testing kits.

**What will happen if you take part in this research study?**

There are several steps to this study. if you volunteer to participate in this study you will have the opportunity to participate in the following components:

1. Allow me to trace your male partner, or take me to your male partner in order to invite him to participate in the study as well. Note, you can choose to participate in the study even if your partner refuses or you think your partner would refuse.
2. Complete one or two study visits where a research assistant like myself will interview you and ask you information about yourself, including whether you are married, number of sexual partners, your level of education, information about your experiences with HIV services, and how you feel about HIV testing and treatment services. We will ask you questions today (or a day nearby that is convenient for you) and, if your partner enrolls in the study, we will ask you similar questions again in four months in order to see if anything has changed. Each interview will last about 45minutes. You can refuse a follow-up survey at any point
3. If your partner agrees to participate in the study, he will be randomized to one of three interventions. We will do the randomization together with him so he can see exactly what intervention he will be offered. The potential interventions are:
1) Standard of care where providers may send him reminders about the benefits of health services.
2) Motivational Interviewing where he can talk to someone about his life, challenges he faces, and strategies to make his life better and additional services as needed.
3) Home-based health services whereby a provider will offer him HIV services and NCD screening at your home as a one-time event. He will then be visited after 4-weeks to be escorted to the clinic if desired.

Regardless of what arm your partner is randomized to, he can always refuse health services or refuse talking to a health care provider and still remain in the study. You can remain in the study regardless of what your partner does.

4. Finally, you may be randomly selected to within 6-months of the study to complete a 1-hour in-depth interview so we can learn more about your experiences in the study. Not all participants will be contacted for the interview and you always have the right to decline an in-depth interview – refusal will not affect your participation in the larger study.

**How long will you be in the research study?**

All study activities will be completed within 6-months of today.

**Are there any potential risks or discomforts that you can expect from this study?**

You will be asked a series of questions by a research assistant about your sexual relationship and your perceptions of your partners use of HIV and other health services.  We will NEVER disclose your HIV status to your partner. We will NEVER disclose to your partner that you told us he had tested HIV+. However, you may feel uncomfortable answering some questions asked during the interview or you may feel comfortable having your partner in the study.  You are able to withdraw from the study at any time. During an interview you can say “I don’t want to answer” to any questions that make you uncomfortable.  All questions will be asked in a private place so that no one else will hear your answers.

If you experience distress or adverse events as a result of the study, we will provide you with counseling resources or refer you to resources for assistance.

**Are there any potential benefits to participating?**

You will have the opportunity to discuss information about your well-being, your relationship with your partner, and HIV services for men with a Research Assistant in a confidential, private manner.

**Are there any potential benefits to society?**

Information obtained as part of this work may be of benefit to the larger Malawi program, or similar programs in sub-Saharan Africa, since the work aims to determine if there are better ways to offer HIV services to men who use HIV self-test kits. If researchers better understand what type of programs work better for men, the program in Malawi can be scaled up and strengthened to provide these specific types of increased support.

**Will you receive payment for being part of this study?**

Your participation is entirely voluntary. You will be provided MK 7,500 (equivalent to 10USD) for each survey completed (MK 15,000 / 20USD across the duration of the study). You will receive the above compensation regardless if you use HIV services or not. Those who complete the additional in-depth interview 6-months after study enrollment will receive an additional MK 7,500 (equivalent to 10USD) for their time.

**What is the cost of participating in this study?**

There is no cost to participate in this study.

**Will information about me be kept confidential?**

The study team are the only people who will know about you or any information that you provide in this study. If necessary to protect your rights or welfare (for example, if you are injured and need emergency care) or if required by Malawian law, specific information about you may be made available to providers or officials.

Authorized representatives of the Malawi National Health Sciences Research Council who are responsible for ensuring the rules related to research are followed, may need to review records of study participants. As a result, they may see your name; but they will not to reveal your identity to others.

When the results of the research are published or discussed in meetings, no information will be included that would reveal your identity. Any paperwork related to the study which contains information about you will be kept in a locked cabinet in a locked office.  Only staff members of the study will have access to this information.  A code will be assigned to each individual participating in the study.  This code will be stored on a computer in a locked file. The key to unlock the information will only be known by the research staff.  All data entered into a computer will be entered using this code so information will no longer have any information that can identify you such as your name. Forms containing any identifying information will be destroyed two years after the study is finished.

**Participation and Withdrawal**

Your participation in this research is VOLUNTARY. If you choose not to participate, that will not affect your relationship with the hospital, your health provider or health centre you usually get your medical care from, or your right to health care. If you decide to participate, you are free to withdraw your consent and stop your participation at any time and can still receive future health care at the hospital or health center you go to.

- **Withdrawal of Participation by the Investigator**

The research investigator may stop your participation in this research if he or she feels this is best for you. The investigators will make the decision and let you know if it is not possible for you to continue. The decision may be made to protect your health and safety.

**Who can answer questions I might have about this study?**

In the event of a research related injury or if you experience a problem, please immediately return to the hospital or health centre you go to or contact Khumbo Phiri. The NHSRC Ministry of Health information (Dr. Mitambo) is also provided in case you have questions about your rights as a research participant.

Kusiyitsidwa kutenga nawo mbali mu kafukufuku ndi wakafukufuku

Anthu opangitsa kafukufukuyu akhonza kukuletsani kutenga nawo mbali mukafukufukuyu akaona kuti ndi bwino kuti mutero. Anthu akafukufukuwa azapanga chiganizochi ndikukudziwitsani kuti sizitheka kuti mupitirize. Chiganizochi chitha kupangidwa kuti ateteze thanzi ndi chitetezo chanu.

Investigator

Khumbo Phiri

Partners in Hope Clinic
Area 36, Plot8

M1 Road South
Lilongwe, Malawi

OR

Dr.C. Mitambo
The Secretariate, NHSRC
Ministry of Health
P.O Box 30377
Lilongwe 3
Cell +265888344 443

**SIGNATURE OF RESEARCH SUBJECT [OR LEGAL REPRESENTATIVE]**

I have read (or someone has read to me) the information provided above. I have been given an opportunity to ask questions and all of my questions have been answered to my satisfaction. I have been given a copy of this form.

Ndawerenga (kapena munthu wina wandiwerengera) zonse zalembedwa mwambamu. Ndapatsidwa mwayi wofunsa mafunso ndi mafunso onse ndinafunsa ayankhidwa ndipo ndakhutusidwa. Ndapasidwa pepala ina yangati yomweyi.

**BY SIGNING THIS FORM, I WILLINGLY AGREE TO PARTICIPATE IN THE RESEARCH:**

____________________________________

Name of Subject

____________________________________	Fingerprint

Name of Legal Representative (if applicable)

____________________________________	DATE (DAY/MO/YR): _________________

Signature of Subject or Legal Representative
(may place an X OR fingerprint if unable to sign)

**SIGNATURE OF INVESTIGATOR OR DESIGNEE**

I have explained the research to the subject or his/her legal representative and answered all of his/her questions. I believe that he/she understands the information described in this document and freely consents to participate.

_____________________________

Name of Investigator or Designee

_____________________________	__________________________

Signature of Investigator or Designee	Date (must be the same as subject’s)

#### 14.4. APPENDIX D: Written Informed Consent – Male

**WRITTEN INFORMED CONSENT**

**Identifying efficient linkage strategies for HIV self-testing (IDEaL)**

Male

You are asked to participate in a research study entitled *“Identifying efficient linkages strategies for HIVST****”*** conducted by Partners in Hope and the University of California Los Angeles in the United States. You are being requested to take part in the study because you recently received a self-test kit and reported recently testing HIV-positive. Your participation in this study is entirely voluntary. You will be read the information below, and you are free to ask questions about anything you do not understand, before deciding whether or not to participate. ***I as the field assistant for this study will take you through this consenting process.***

**Why is this study being done?**

HIV self-testing is very helpful for people who want to know their status but do not usually go to the health facility. However, it can be hard for individuals who test HIV-positive with HIV self-testing to be able to access other health services. Researchers want to determine what are the best interventions that can help men who are diagnosed with HIV use other health services, if desired. The study will offer several different strategies for HIV services to see what works best for men who use HIV self-testing kits.

**What will happen if you take part in this research study?**

There are several steps to this study. If you volunteer to participate in this study you will have the opportunity to participate in the following components:

1. Complete two study visits where a research assistant like myself will interview you and ask you information about yourself, including whether you are married, number of sexual partners, your level of education, information about your experiences with HIV services, and how you feel about HIV testing and treatment services. We will ask you questions today (or a day that is convenient for you) and in four months in order to see if anything has changed. Each interview will last about 45minutes. You can refuse a follow-up survey at any point.

2. After completing the first interview you will be randomized to one of three interventions. We will do the randomization together so you can see exactly what you will be offered. Based on what intervention you are randomly selected for, you will also be offered a variety of health services from health facility staff. The potential interventions are:

a) Reminders about the benefits of health services;

b) Motivational Interviewing where you can talk to someone about your life, challenges you face, and strategies to make your life and better, and additional services as needed

c) Home-based health services whereby a provider will offer you HIV services and NCD screening at your home as a one-time event. After four weeks, you will be visited by a health care worker who will escort you to the clinic for continued health services if desired.

Regardless of what arm you are randomized to, you can always refuse health services or refuse talking to a health care provider and still remain in the study. Even if you do not plan to use any additional health services, don’t worry, you can still be in the study and we can still complete the interviews we discussed above.

3. In the first 6 months of the study, we will also review medical records at your local health facility to see if you visited the health facility since enrolling in the study. This does not require interaction with you and will be completely confidential.

4. Finally, we may contact you within 6-months of the study to complete another in-depth interview so we can learn more about your experience with the study, recommendations for future interventions, and what additional services, if any, you would like. Not all participants will be contacted for the interview and you always have the right to decline an in-depth interview – refusal will not affect your participation in the larger study.

**How long will you be in the research study?**

All study activities will be completed within 6-months of today.

**Are there any potential risks or discomforts that you can expect from this study?**

You will be asked a series of questions by a research assistant about your sexual history and your experience receiving and using a self-test kit from your sexual partner.  You may feel uncomfortable answering some of the questions asked by the interviewer.  You can say “I don’t want to answer” to any questions that make you uncomfortable.  All questions will be asked in a private place so that no other patients or staff will hear your answers.

If you agree to participate in the assigned intervention, providers may ask to reach you at home or in the community. As with any health service, if you choose to initiate ART you may be at risk of unwanted status disclosure.

If you experience distress or adverse events, we will provide you with counseling resources or refer you to resources for assistance.

**Are there any potential benefits to participating?**

You will have the opportunity to discuss information about your well-being and HIV services with a Research Assistant and possibly a health care provider and in a confidential, private manner. You will also have the chance to link to HIV care services at the facility of your choosing.

**Are there any potential benefits to society?**

Information obtained as part of this work may be of benefit to the larger Malawi program, or similar programs in sub-Saharan Africa, since the work aims to determine if there are better ways to offer HIV services to men who use HIV self-test kits. If researchers better understand what type of programs work better for men, the program in Malawi can be scaled up and strengthened to provide these specific types of increased support.

**Will you receive payment for being part of this study?**

Your participation is entirely voluntary. You will be provided MK 7,500 (equivalent to 10USD) for each survey completed (MK 15,000 / 20USD across the duration of the study). You will receive the above compensation regardless if you use HIV services or not. Those who complete the additional in-depth interview 6-months after study enrollment will receive an additional MK 7,500 (equivalent to 10USD) for their time.

**What is the cost of participating in this study?**

There is no cost to participate in this study.

**Will information about me be kept confidential?**

Authorized representatives of the Malawi National Health Sciences Research Council who are responsible for ensuring the rules related to research are followed, may need to review records of study participants. As a result, they may see your name; but they will not to reveal your identity to others.

When the results of the research are published or discussed in meetings, no information will be included that would reveal your identity. Any paperwork related to the study which contains information about you will be kept in a locked cabinet in a locked office.  Only staff members of the study will have access to this information.  A code will be assigned to each individual participating in the study.  This code will be stored on a computer in a locked file. The key to unlock the information will only be known by the research staff.  All data entered into a computer will be entered using this code so information will no longer have any information that can identify you such as your name. Forms containing any identifying information will be destroyed two years after the study is finished.

**Participation and Withdrawal**

Your participation in this research is VOLUNTARY. If you choose not to participate, that will not affect your relationship with the hospital, your health provider or health centre you usually get your medical care from, or your right to health care. If you decide to participate, you are free to withdraw your consent and stop your participation at any time and can still receive future health care at the hospital or health center you go to.

- **Withdrawal of Participation by the Investigator**

The research investigator may stop your participation in this research if he or she feels this is best for you. The investigators will make the decision and let you know if it is not possible for you to continue. The decision may be made to protect your health and safety.

**Who can answer questions I might have about this study?**

In the event of a research related injury or if you experience a problem, please immediately return to the hospital or health centre you go to or contact Mike Nyirenda. The NHSRC Ministry of Health information (Dr. Kathyola) is also provided in case you have questions about your rights as a research participant.

Investigator:

Khumbo Phiri

Partners in Hope Clinic
Area 36, Plot8

M1 Road South
Lilongwe, Malawi

OR

Dr.C. Mitambo
The Secretariate, NHSRC
Ministry of Health
P.O Box 30377
Lilongwe 3
Cell +265888344 443

**SIGNATURE OF RESEARCH SUBJECT [OR LEGAL REPRESENTATIVE]**

I have read (or someone has read to me) the information provided above. I have been given an opportunity to ask questions and all of my questions have been answered to my satisfaction. I have been given a copy of this form.

**BY SIGNING THIS FORM, I WILLINGLY AGREE TO PARTICIPATE IN THE RESEARCH:**

____________________________________

Name of Subject

____________________________________

Name of Legal Representative (if applicable)

____________________________________	DATE (DAY/MO/YR): _________________

Signature of Subject or Legal Representative
(may place an X OR fingerprint if unable to sign)

**SIGNATURE OF INVESTIGATOR OR DESIGNEE**

I have explained the research to the subject or his/her legal representative and answered all of his/her questions. I believe that he/she understands the information described in this document and freely consents to participate.

_____________________________

Name of Investigator or Designee

_____________________________	__________________________

Signature of Investigator or Designee	Date (must be the same as subject’s)

#### 14.5. APPENDIX E: Baseline Survey – Male

**BASELINE SURVEY**

**Identifying efficient linkage strategies for HIV self-testing (IDEaL)**

Male

| **Question Name** | **Label** | **Responses** |
| --- | --- | --- |
| INTRODUCTION SECTION | | |
| interviewer | Full Name of Interviewer |  |
| Interview date | Interview date |  |
| Time start | Time survey started |  |
| District | District |  |
| TA | TA |  |
| village | Village |  |
| SECTION A: DEMOGRAPHICS | | |
| Intro Note | Thank you for agreeing to participate. Now I will ask you a few questions about yourself and who you are. Please feel free to answer honestly. There are no right or wrong answers. |  |
| a7 | What is your tribe? | 1. Lomwe 2. Sena 3. Chewa 4. Mang'anja/Nyanja 5. Ngoni 6. Tumbuka 7. Tonga 8. Yao   99. Other, specify |
| a3 | What is the highest level of school you attended? | 1. Primary 2. Secondary 3. Higher |
| a3b | What class did you complete in your highest level of school? | ____________________ |
| a4 | Please think of the past 12months, how would you describe your primary occupation? | 1. Working formally (employed full time) 2. Working informally (ganyu, farming, business) 3. Not working |
| a5 | Are you currently married? | 1. Married 2. Live-in partner 3. Steady Girlfriend/Boyfriend 4. Separated 5. Divorced   99. Other, specify |
| a6 | How many living children do you have? | ______________________ |
| a6b | What is the age of your youngest child? | _______________________ |
| a6bc | What age is the child (in years or months) | _______________________ |
| a4b | How many children currently live with you? | _______________________ |
| a7 | How many sexual partners have you had in the past 12 months? | _______________________ |
| a8 | Have you had sex with someone besides your wife/husband without a condom in the past 12 months? | 1. Yes   0. No  88. Don’t know/ Not sure  89. Refused to answer |
| a8b | Have you had sex without a condom in the past 12 months? | 1. Yes   0. No  88. Don’t know/ Not sure  89. Refused to answer |
| SECTION B: INCOME QUESTIONS | | |
| Intro Note | I will now discuss with you about the valuable items that you or your household possesses. As I will be chatting with you I will also some questions about money you have and activities that you indulge in to find money. |  |
| b1 | Please think of the past 12 months, how would you describe your primary occupation? | 1. Working formally (employed full time) 2. Working informally (ganyu, farming, business) 3. Not working |
| b1b | Think about all the work you have done in the past month. How many days did you normally work this month that gave you pay? | __________________ |
| b2 | Do you have any savings for the future, such as a bank account, savings group or cash? | 1. Yes   0. No |
| Household Assets | | |
| b3 | Does your household have:  *The respondent said that his/her household doesn't have any of the household assets. Please probe and ensure that this is correct before you proceed.* |  |
| b3_1 | Metal Roof? | 1. Yes   0. No |
| b3_2 | Electricity? | 1. Yes   0. No |
| b3_3 | Paraffin lamp with no glass? | 1. Yes   0. No |
| b3_4 | A paraffin lamp? | 1. Yes   0. No |
| b3_5 | A radio? | 1. Yes   0. No |
| b3_6 | A television? | 1. Yes   0. No |
| b3_7 | A cellular phone? | 1. Yes   0. No |
| b3_8 | A bed? | 1. Yes   0. No |
| b3_9 | A sofa set? | 1. Yes   0. No |
| b3_10 | A table? | 1. Yes   0. No |
| b3_11 | A refrigerator | 1. Yes   0. No |
| b3_12 | Mattress? | 1. Yes   0. No |
| b3_13 | Chair(s)? | 1. Yes   0. No |
| b3_14 | Cattle? | 1. Yes   0. No |
| b3_15 | Goat? | 1. Yes   0. No |
| b3_16 | Sheep? | 1. Yes   0. No |
| b3_17 | Pigs? | 1. Yes   0. No |
| b3_18 | Donkey? | 1. Yes   0. No |
| b3_19 | Chickens? | 1. Yes   0. No |
| b3_20 | Other poultry? | 1. Yes   0. No |
| b4 | In the past 30 days, have you drank beer? | 1. Yes   0. No |
| b4b | How many days in the past 30 days have you drank beer? | ______________________ |
| b4c | How much money did you spend on beer the last time you went? | MWK: |
| b4d | In total, approximately how much money did you spend on beer in the past 30 days? | MWK: |
| Relationship | | |
| Intro Note | Now I'd like to talk to you about your current sexual relationship |  |
| f8 | How long have you been/were you in a sexual relationship with your partner? | Days_______________  Months_____________  Years________________ |
| f9 | Do you have children with your partner? How many children? | ______________________ |
| f10 | How often do you currently talk to your partner? | 1. Everyday 2. A couple times a week 3. Once a week 4. A couple times a month 5. Once a month 6. Less than once a month 7. Not at all (never) |
| f10b | In a typical month, who earns more money? You, or your partner? | 1. Myself 2. This partner 3. We earn the same amount   88. Don't know |
| Decision Making | | |
| Intro Note | Now I would like to talk to you about how you and your partner make decisions. |  |
| f11 | Who usually decides how the money you earn will be used? | 1. Yourself (Respondent) 2. Jointly (This partner and you together) 3. Mainly this partner 4. Someone else 5. Do not earn money   88. Refuse to say |
| f11b | (if above question=4 ) Who decides? | ______________________ |
| f12 | Who usually decides how your partner's earnings will be used? | 1. Yourself (Respondent) 2. Jointly (This partner and you together) 3. Mainly this partner 4. Someone else 5. Do not earn money   88. Refuse to say |
| f12b | (if above question=4 ) Who decides? | ______________________ |
| f13 | Who usually makes decisions about health care for yourself? | 1. Yourself (Respondent) 2. Jointly (This partner and you together) 3. Mainly this partner 4. Someone else 5. Not applicable/ Don’t have children   88. Refuse to say |
| f13b | (if above question=4 ) Who decides? | ______________________ |
| f14 | Who usually makes decisions about health care for your child with this partner? | 1. Yourself (Respondent) 2. Jointly (This partner and you together) 3. Mainly this partner 4. Someone else 5. Not applicable/ Don’t have children   88. Refuse to say |
| f14b | (if above question=4 ) Who decides? | ______________________ |
| f15 | Who usually makes decisions about health care for your partner? | 1. Yourself (Respondent) 2. Jointly (This partner and you together) 3. Mainly this partner 4. Someone else 5. Not applicable/ Don’t have children   88. Refuse to say |
| f15b | (if above question=4 ) Who decides? | ______________________ |
| f16 | Who usually makes decisions about making major household purchases? | 1. Yourself (Respondent) 2. Jointly (This partner and you together) 3. Mainly this partner 4. Someone else 5. Not applicable/ Don’t have children   88. Refuse to say |
| f16b | (if above question=4 ) Who decides? |  |
| Note | I would like to ask you questions about [probability/chance/likelihood] that certain things will happen. There are ten beans in this cup. I will ask you to pick some of the beans and put them in the plate. The number of beans that you are going to put in the plate will reflect the probability that something will happen. One bean means there is very little chance that something will happen. If you do not put any bean in the plate it means you are certain that there is no likelihood that something will happen. |  |
| note2 | If you put additional beans in the plate it means the chance that something will happen will also increase. For example, if you put one or two beans in the plate, it means there is little chance that something will happen. Even though there is little chance but it can happen. If you put ten beans it means there is equal chance of something happening or not. If you put six beans it means the chance that something will happen is slightly greater than not happening. If you put all ten beans, it means you are certain that whatever the case something will really happen. There is no wrong or right answer I just want to know what you think. |  |
| note3 | INTERVIEWER: Report for each question the NUMBER OF BEANS put in the PLATE. After each question, replace the beans on the table (unless otherwise noted). |  |
| Practice | | |
| pr1 | Pick the number of beans that reflects how likely you think it is that: | __________________ |
| pr1b | You will go to the market at least once within the next 2 days. | __________________ |
| pr1c | You will go to the market at least once within the next 2 weeks. | __________________ |
| Practice | | |
| pr2 | INTERVIEWER: Did Respondent add any beans between pr1b and pr1c? | 1. Yes   0. No |
| pr3 | Remember, as time goes by, you may find more time to go to the market. Therefore, you should have added beans to the plate. Let me ask you again. Now, add beans in the plate so that the number of beans in the plate reflects how likely you think it is that you will go to the market at least once within 2 weeks.  How likely you think it is that you will go to the market at least once within 2 weeks? | __________________ |
| f17 | Pick the number of beans that reflects how likely you think: |  |
| f17b | You will still be married/with [partner one year from now. | __________________ |
| f17c | You are currently infected with HIV/AIDS | __________________ |
| f17d | You will become infected with HIV/AIDS during the next 12 months | __________________ |
| f17e | You will become infected with HIV/AIDS during their lifetime | __________________ |
| f17c | partner is infected with HIV/AIDS now. | __________________ |
| f17d | partner will become infected with HIV/AIDS during the next 12 months. | __________________ |
| SELF REPORTED HEALTH AND HAPPINESS | | |
| Intro Note | Now I'd like to talk to you about how healthy and happy you feel. |  |
| e1 | I am interested in your general level of well-being or satisfaction with life. How satisfied are you with your life, all things considered? | 1. Very satisfied 2. Somewhat satisfied 3. Neutral 4. Somewhat unsatisfied 5. Very unsatisfied |
| e2 | Do you think that you are more, equally or less satisfied than other persons your age and sex living in your village? | 1. More satisfied 2. Equally satisfied 3. Less satisfied |
| e3 | In general, would you say your health now is: very good, good, poor or very poor? | 1. Very good 2. Good 3. Poor 4. Very poor |
| e4 | How would you compare your health to other people of the same age and sex in your village? | 1. More healthy 2. Equally healthy 3. Less healthy |
| e5 | In the past month, how many days were you too sick to work/go to school/complete household chores? | ___________________ |
| Happiness | | |
| e6 | How true are the following statements for you in the last month? |  |
| e6_1 | I have felt depressed | 1. Strongly Agree 2. Agree 3. Disagree 4. Strongly Disagree |
| e6_2 | I have felt life was not worth living | 1. Strongly Agree 2. Agree 3. Disagree 4. Strongly Disagree |
| e6_3 | I have felt content. | 1. Strongly Agree 2. Agree 3. Disagree 4. Strongly Disagree |
| e6_4 | I have felt lonely | 1. Strongly Agree 2. Agree 3. Disagree 4. Strongly Disagree |
| GENDER EQUITABLE MEN SCALE | | |
| Note | Please tell me if you strongly agree, agree, disagree, or strongly disagree with the following statements: |  |
| j1 | Woman’s most important role is to take care of her home and cook (take care of home is about housekeeping) | 1. Strongly Agree 2. Agree 3. Unsure 4. Disagree 5. Strongly Disagree   88. Refuse to say |
| j2 | Men need sex more than women | 1. Strongly Agree 2. Agree 3. Unsure 4. Disagree 5. Strongly Disagree   88. Refuse to say |
| j3 | Men don’t talk about sex, they just do it. | 1. Strongly Agree 2. Agree 3. Unsure 4. Disagree 5. Strongly Disagree   88. Refuse to say |
| j4 | There are times when a woman deserves to be beaten | 1. Strongly Agree 2. Agree 3. Unsure 4. Disagree 5. Strongly Disagree   88. Refuse to say |
| j5 | Changing diapers, giving kids a bath & feeding kids are mother’s responsibility | 1. Strongly Agree 2. Agree 3. Unsure 4. Disagree 5. Strongly Disagree   88. Refuse to say |
| j6 | It is a woman’s responsibility to avoid getting pregnant | 1. Strongly Agree 2. Agree 3. Unsure 4. Disagree 5. Strongly Disagree   88. Refuse to say |
| j7 | A man should have the final word about decisions in his home | 1. Strongly Agree 2. Agree 3. Unsure 4. Disagree 5. Strongly Disagree   88. Refuse to say |
| j8 | Men are always ready to have sex | 1. Strongly Agree 2. Agree 3. Unsure 4. Disagree 5. Strongly Disagree   88. Refuse to say |
| j9 | A woman should tolerate violence in order to keep her family together | 1. Strongly Agree 2. Agree 3. Unsure 4. Disagree 5. Strongly Disagree   88. Refuse to say |
| j10a | I would be outraged if my wife asked me to use a condom. | 1. Strongly Agree 2. Agree 3. Unsure 4. Disagree 5. Strongly Disagree   88. Refuse to say |
| j10b | Men would be outraged if their wife asked them to use a condom | 1. Strongly Agree 2. Agree 3. Unsure 4. Disagree 5. Strongly Disagree   88. Refuse to say |
| j11 | A man and a woman should decide together what type of contraceptive to use | 1. Strongly Agree 2. Agree 3. Unsure 4. Disagree 5. Strongly Disagree   88. Refuse to say |
| j12 | I would never have a homosexual friend | 1. Strongly Agree 2. Agree 3. Unsure 4. Disagree 5. Strongly Disagree   88. Refuse to say |
| j13a | If someone insults me, I will defend my reputation, with force if I have to. | 1. Strongly Agree 2. Agree 3. Unsure 4. Disagree 5. Strongly Disagree   88. Refuse to say |
| j13b | If someone insults a man, he should defend his reputation, with force if he has to | 1. Strongly Agree 2. Agree 3. Unsure 4. Disagree 5. Strongly Disagree   88. Refuse to say |
| j14 | To be a man you need to be tough. | 1. Strongly Agree 2. Agree 3. Unsure 4. Disagree 5. Strongly Disagree   88. Refuse to say |
| j15 | Men should be embarrassed if unable to get an erection | 1. Strongly Agree 2. Agree 3. Unsure 4. Disagree 5. Strongly Disagree   88. Refuse to say |
| j16 | If a guy gets a woman pregnant, child is the responsibility of both the man and woman | 1. Strongly Agree 2. Agree 3. Unsure 4. Disagree 5. Strongly Disagree   88. Refuse to say |
| j17 | A man should know what his partner likes during sex | 1. Strongly Agree 2. Agree 3. Unsure 4. Disagree 5. Strongly Disagree   88. Refuse to say |
| j18 | The participation of the father is important in raising children | 1. Strongly Agree 2. Agree 3. Unsure 4. Disagree 5. Strongly Disagree   88. Refuse to say |
| j19 | It’s important for men to have friends to talk about their problems | 1. Strongly Agree 2. Agree 3. Unsure 4. Disagree 5. Strongly Disagree   88. Refuse to say |
| j20 | A couple should decide together if they want to have children. | 1. Strongly Agree 2. Agree 3. Unsure 4. Disagree 5. Strongly Disagree   88. Refuse to say |
| STIGMA | | |
| Note | In this next section I'd like to discuss your thoughts about people living with HIV in your community. Please feel free to talk openly, there is no right or wrong answer. I am interested in your own thoughts. |  |
| i3 | I would buy fresh vegetables from a shopkeeper or vendor if I knew that this person had HIV | 1. Strongly Agree 2. Agree 3. Neutral 4. Disagree 5. Strongly Disagree |
| i4 | If a member of my family became sick with AIDS, I would be willing to care for her or him in our own household | 1. Strongly Agree 2. Agree 3. Neutral 4. Disagree 5. Strongly Disagree |
| i5 | In my opinion, if a female teacher has HIV but is not sick, she should be allowed to continue teaching in the school | 1. Strongly Agree 2. Agree 3. Neutral 4. Disagree 5. Strongly Disagree |
| EXPECTATIONS | | |
| Intro Note | I would like to ask you questions about [probability/chance/likelihood] that certain things will happen. There are ten beans in this cup. I will ask you to pick some of the beans and put them in the plate. The number of beans that you are going to put in the place will reflect the probability that something will happen. One beans means there is very little chance that something will happen. If you do not put any bean in the plate it means you are certain that there is no likelihood that something will happen. |  |
| h4 | Pick the number of beans that reflects how likely you think it is that: | ______________________ |
| h4b | You will have to rely on family members for financial assistance in the next 12 months. | ______________________ |
| h4c | You will have to provide some family members with financial assistance in the next 12 months. | ______________________ |
| Note | Next, I would like to ask you a few questions about what you expect in the future. I know that nobody knows for sure what the future may bring, but let`s just talk about your best guess. |  |
| h5 | In the next year how likely is it that you will: |  |
| h5a | You will be enrolled in school one year from now | ______________________ |
| h5b | Start a new business? | ______________________ |
| h5c | Open a bank account? | ______________________ |
| h5d | Buy land? | ______________________ |
| h5e | Save money? | ______________________ |
| h5f | Experience shortage of food? | ______________________ |
| h5g | Have steady work? | ______________________ |
| Tested for HIV | | |
| pd2 | Approximately how many times have you ever been tested for HIV?  *Enter "-99" if client doesn’t remember* | ______________________ |
| pd3 | When was the last time you were tested for HIV? | a. Year______ b. Month________ |
| pd5 | Think about the very first time you received an HIV+ test result. Since that very first HIV+ test result, have you ever tested for HIV again (excluding a confirmatory test)? | 1. Yes   0. No |
| pd6 | Have you ever initiated ART? | 1. Yes   0. No |
| First Initiated ART | | |
| pd6b | When did you first initiate ART? | a. Year_______ b. Month____ |
| pd7 | Have you ever been >14 days late for an ART appointment? | 1. Yes   0. No |
| pd7b | How many times? | _____________________ |
| pd8 | Do you know anyone who is on ART? | 1. Yes   0. No |
| pd8b | Now think about the person on ART who you are closest with.   How often do you talk with them about ART? | 1. Everyday 2. A couple times a week 3. Once a week 4. A couple times a month 5. Once a month 6. Less than once a month 7. Not at all (never) |
| pd9 | Have you disclosed your HIV status to anyone besides your partner? | 1. Yes   0. No |
| pd9b | Who else did you disclose to?  *Mark all that apply* | 1. Sister 2. Brother 3. Father 4. Mother 5. Uncle 6. Aunt 7. Friend 8. Mother-in-Law 9. Father-in-Law 10. My children 11. Employee 12. Other sexual partner   99. Other, specify_____ |
| pd10 | Of those people you disclosed to, who do you talk to most often? | 1. Sister 2. Brother 3. Father 4. Mother 5. Uncle 6. Aunt 7. Friend 8. Mother-in-Law 9. Father-in-Law 10. My children 11. Employee 12. Other sexual Partner   99. Other, specify_____ |
| pd10c | How often do you talk to that person? | 1. Everyday 2. A couple times a week 3. Once a week 4. A couple times a month 5. Once a month 6. Less than once a month 7. Not at all (never) |
| PREVIOUS USE OF HEALTH SERVICES | | |
| Intro Note | Now I'd like to talk to you about your experience with using health services at health facilities. |  |
| h1 | Have you gone to a health facility in the past 12 months (either for yourself or someone else - AKA as a guardian)? | 1. Yes   0. No |
| h2 | How many times have you gone to the health facility in the past 12 months? | ______________________ |
| h3 | Now think about yourself specifically. How many times have you gone to a health facility in the past 12 months for your own health care? | _______________________ |
| h_a1 | When was the last time (the YEAR) you went to a health facility for YOUR OWN health?  NOTE: PUT WHAT YEAR. (i.e., 2015). If DO NOT REMEMBER, help them estimate. IF NEVER GONE, put -99 | ______________________ |
| h4 | What services did you receive at your last health facility visit for your own health? | 1. ANC 2. Family Planning 3. Delivery 4. Post-natal 5. Under Five 6. HTC 7. ART 8. Feeling sick (OPD) 9. Injury (OPD) 10. Dentist 11. None   99. Other specify_____ |
| h_a2 | Now think about the SECOND most recent time you went to a health facility for YOUR OWN health. What year did you go to the health facility? | ______________________ |
| c3 | Now please think about your MOST RECENT visit to a health facility, excluding today. When did you go? | a. Year___________ b. Month_________ |
| c4 | Which facility did you go to? | 1. Current facility  0. Other facility, specify_______________ |
| c5 | What was the main service you went for? | 1. ANC 2. Family Planning 3. Delivery 4. Post-natal 5. Under Five 6. HTC 7. ART 8. Feeling sick (OPD) 9. Injury (OPD) 10. Dentist 11. None   99. Other specify_____ |
| c5b | Who received services? | 1. Myself 2. My child 3. My partner 4. Another family member 5. A friend   99. Other, specify_______ |
| c6a | Did you (or the person you came with) receive another service? | 1. Yes   0. No |
| c6 | What was the second service you went for? | 1. ANC 2. Family Planning 3. Delivery 4. Post-natal 5. Under Five 6. HTC 7. ART 8. Feeling sick (OPD) 9. Injury (OPD) 10. Dentist 11. None   99. Other specify_____ |
| c6b | Who received services? | 1. Myself 2. My child 3. My partner 4. Another family member 5. A friend   99. Other, specify_______ |
| Service Satisfaction | | |
| c10 | Now I would like to talk to you about your satisfaction with the services you received that day. Please tell me whether any of these were problems for you at the VISIT YOU ARE THINKING ABOUT NOW, and if so, whether they were major or minor problems for you. |  |
| c10_1 | Time you waited to see a provider | 1. Major 2. Minor   0. No-problem  88. Not applicable  89. Don’t know |
| c10_2 | Ability to discuss problems or concerns about your pregnancy | 1. Major 2. Minor   0. No-problem  88. Not applicable  89. Don’t know |
| c10_3 | Amount of explanation you received about the problem or treatment | 1. Major 2. Minor   0. No-problem  88. Not applicable  89. Don’t know |
| c10_4 | Privacy from having others see the examination | 1. Major 2. Minor   0. No-problem  88. Not applicable  89. Don’t know |
| c10_5 | Privacy from having others hear your consultation discussion | 1. Major 2. Minor   0. No-problem  88. Not applicable  89. Don’t know |
| c10_6 | Availability of medicines at this facility | 1. Major 2. Minor   0. No-problem  88. Not applicable  89. Don’t know |
| c10_7 | The hours of service at this facility, i.e., when they open and close | 1. Major 2. Minor   0. No-problem  88. Not applicable  89. Don’t know |
| c10_8 | The number of days services are available to you | 1. Major 2. Minor   0. No-problem  88. Not applicable  89. Don’t know |
| c10_9 | The cleanliness of the facility | 1. Major 2. Minor   0. No-problem  88. Not applicable  89. Don’t know |
| c10_10 | How the staff treated you | 1. Major 2. Minor   0. No-problem  88. Not applicable  89. Don’t know |
| c10_11 | Cost for services or treatments | 1. Major 2. Minor   0. No-problem  88. Not applicable  89. Don’t know |
| Satisfaction | | |
| c11 | In general, which of the following statements best describes your opinion of the services you either received or were provided at the facility | 1. I am very satisfied with the services I received 2. I am satisfied with the services I received 3. I am not satisfied with the services I received 4. I am very dissatisfied with the services I received |
| c12 | Did you recommend this health facility to a friend or family member? | 1. Yes   0. No |
| Comment | We have reached the end of the chat. Thank you for your time. Do you have anything else you would like to say? |  |
| End Note | Thank the participant for their time and give them transport reimbursement, if they did not come for an ART appointment |  |
| Comments | Enumerator comments |  |
| End of the survey! | | |

#### 14.6. APPENDIX F: Baseline survey – Female

**BASELINE SURVEY**

**Identifying efficient linkage strategies for HIV self-testing (IDEaL)**

Female

| INTRODUCTION SECTION | |
| --- | --- |
| interviewer | Full Name of Interviewer |
| Interview date | Interview date |
| Time start | Time survey started |
| District | District |
| Facility | Facility |
| ID | ID |

| SECTION A: DEMOGRAPHICS | | |
| --- | --- | --- |
| Intro Note | Thank you for agreeing to participate. Now I will ask you a few questions about yourself and who you are. Please feel free to answer honestly. There are no right or wrong answers. |  |
| a7 | What is your tribe? | 1. Lomwe 2. Sena 3. Chewa 4. Mang'anja/Nyanja 5. Ngoni 6. Tumbuka 7. Tonga 8. Yao   99. Other, specify |
| a3 | What is the highest level of school you attended? | 1. Primary 2. Secondary 3. Higher |
| a3b | What class did you complete in your highest level of school? | ____________________ |
| a4 | Please think of the past 12months, how would you describe your primary occupation? | 1. Working formally (employed full time) 2. Working informally (ganyu, farming, business) 3. Not working |
| a5 | Are you currently married? | 1. Married 2. Live-in partner 3. Steady Boyfriend 4. Separated 5. Divorced   99. Other, specify |
| a6 | How many living children do you have? | ______________________ |
| a6b | What is the age of your youngest child? | _______________________ |
| a6bc | What age is the child (in years or months) | _______________________ |
| a4b | How many children currently live with you? | _______________________ |
| a7 | How many sexual partners have you had in the past 12 months? | _______________________ |
| a8 | Have you had sex with someone besides your husband without a condom in the past 12 months? | 2. Yes   0. No  88. Don’t know/ Not sure  89. Refused to answer |
| a8b | Have you had sex without a condom in the past 12 months? | 2. Yes   0. No  88. Don’t know/ Not sure  89. Refused to answer |
| SECTION B: INCOME QUESTIONS | | |
| Intro Note | I will now discuss with you about the valuable items that you or your household possesses. As I will be chatting with you I will also some questions about money you have and activities that you indulge in to find money. |  |
| b1 | Please think of the past 12 months, how would you describe your primary occupation? | 1. Working formally (employed full time) 2. Working informally (ganyu, farming, business) 3. Not working |
| b1b | Think about all the work you have done in the past month. How many days did you normally work this month that gave you pay? | __________________ |
| b2 | Do you have any savings for the future, such as a bank account, savings group or cash? | 2. Yes   0. No |
| Household Assets | | |
| b3 | Does your household have:  *The respondent said that his/her household doesn't have any of the household assets. Please probe and ensure that this is correct before you proceed.* |  |
| b3_1 | Metal Roof? | 2. Yes   0. No |
| b3_2 | Electricity? | 2. Yes   0. No |
| b3_3 | Paraffin lamp with no glass? | 2. Yes   0. No |
| b3_4 | A paraffin lamp? | 2. Yes   0. No |
| b3_5 | A radio? | 2. Yes   0. No |
| b3_6 | A television? | 2. Yes   0. No |
| b3_7 | A cellular phone? | 2. Yes   0. No |
| b3_8 | A bed? | 2. Yes   0. No |
| b3_9 | A sofa set? | 2. Yes   0. No |
| b3_10 | A table? | 2. Yes   0. No |
| b3_11 | A refrigerator | 2. Yes   0. No |
| b3_12 | Mattress? | 2. Yes   0. No |
| b3_13 | Chair(s)? | 2. Yes   0. No |
| b3_14 | Cattle? | 2. Yes   0. No |
| b3_15 | Goat? | 2. Yes   0. No |
| b3_16 | Sheep? | 2. Yes   0. No |
| b3_17 | Pigs? | 2. Yes   0. No |
| b3_18 | Donkey? | 2. Yes   0. No |
| b3_19 | Chickens? | 2. Yes   0. No |
| b3_20 | Other poultry? | 2. Yes   0. No |
| b4 | In the past 30 days, have you drank beer? | 2. Yes   0. No |
| b4b | How many days in the past 30 days have you drank beer? | ______________________ |
| b4c | How much money did you spend on beer the last time you went? | MWK: |
| b4d | In total, approximately how much money did you spend on beer in the past 30 days? | MWK: |
| Relationship | | |
| Intro Note | Now I'd like to talk to you about your current sexual relationship |  |
| f8 | How long have you been/were you in a sexual relationship with your partner? | Days_______________  Months_____________  Years________________ |
| f9 | Do you have children with your partner? How many children? | ______________________ |
| f10 | How often do you currently talk to your partner? | 1. Everyday 2. A couple times a week 3. Once a week 4. A couple times a month 5. Once a month 6. Less than once a month 7. Not at all (never) |
| f10b | In a typical month, who earns more money? You, or your partner? | 1. Myself 2. This partner 3. We earn the same amount   88. Don't know |
| Decision Making | | |
| Intro Note | Now I would like to talk to you about how you and your partner make decisions. |  |
| f11 | Who usually decides how the money you earn will be used? | 1. Yourself (Respondent) 2. Jointly (This partner and you together) 3. Mainly this partner 4. Someone else 5. Do not earn money   88. Refuse to say |
| f11b | (if above question=4 ) Who decides? | ______________________ |
| f12 | Who usually decides how your partner's earnings will be used? | 1. Yourself (Respondent) 2. Jointly (This partner and you together) 3. Mainly this partner 4. Someone else 5. Do not earn money   88. Refuse to say |
| f12b | (if above question=4 ) Who decides? | ______________________ |
| f13 | Who usually makes decisions about health care for yourself? | 1. Yourself (Respondent) 2. Jointly (This partner and you together) 3. Mainly this partner 4. Someone else 5. Not applicable/ Don’t have children   88. Refuse to say |
| f13b | (if above question=4 ) Who decides? | ______________________ |
| f14 | Who usually makes decisions about health care for your child with this partner? | 1. Yourself (Respondent) 2. Jointly (This partner and you together) 3. Mainly this partner 4. Someone else 5. Not applicable/ Don’t have children   88. Refuse to say |
| f14b | (if above question=4 ) Who decides? | ______________________ |
| f15 | Who usually makes decisions about health care for your partner? | 1. Yourself (Respondent) 2. Jointly (This partner and you together) 3. Mainly this partner 4. Someone else 5. Not applicable/ Don’t have children   88. Refuse to say |
| f15b | (if above question=4 ) Who decides? | ______________________ |
| f16 | Who usually makes decisions about making major household purchases? | 1. Yourself (Respondent) 2. Jointly (This partner and you together) 3. Mainly this partner 4. Someone else 5. Not applicable/ Don’t have children   88. Refuse to say |
| f16b | (if above question=4 ) Who decides? |  |
| Note | I would like to ask you questions about [probability/chance/likelihood] that certain things will happen. There are ten beans in this cup. I will ask you to pick some of the beans and put them in the plate. The number of beans that you are going to put in the plate will reflect the probability that something will happen. One bean means there is very little chance that something will happen. If you do not put any bean in the plate it means you are certain that there is no likelihood that something will happen. |  |
| note2 | If you put additional beans in the plate it means the chance that something will happen will also increase. For example, if you put one or two beans in the plate, it means there is little chance that something will happen. Even though there is little chance but it can happen. If you put ten beans it means there is equal chance of something happening or not. If you put six beans it means the chance that something will happen is slightly greater than not happening. If you put all ten beans, it means you are certain that whatever the case something will really happen. There is no wrong or right answer I just want to know what you think. |  |
| note3 | INTERVIEWER: Report for each question the NUMBER OF BEANS put in the PLATE. After each question, replace the beans on the table (unless otherwise noted). |  |
| Practice | | |
| pr1 | Pick the number of beans that reflects how likely you think it is that: | __________________ |
| pr1b | You will go to the market at least once within the next 2 days. | __________________ |
| pr1c | You will go to the market at least once within the next 2 weeks. | __________________ |
| Practice | | |
| pr2 | INTERVIEWER: Did Respondent add any beans between pr1b and pr1c? | 2. Yes   0. No |
| pr3 | Remember, as time goes by, you may find more time to go to the market. Therefore, you should have added beans to the plate. Let me ask you again. Now, add beans in the plate so that the number of beans in the plate reflects how likely you think it is that you will go to the market at least once within 2 weeks.  How likely you think it is that you will go to the market at least once within 2 weeks? | __________________ |
| f17 | Pick the number of beans that reflects how likely you think: |  |
| f17a | You will still be married/with [partner one year from now. | __________________ |
| f17b | Your partner will become sick during the next 12 months | __________________ |
| f17c | Your partner will start ART treatment in the next 3 months | __________________ |
| f17d | Your partner will disclose your HIV status to your close friends/family in the next 3 months | __________________ |
| SELF REPORTED HEALTH AND HAPPINESS | | |
| Intro Note | Now I'd like to talk to you about how healthy and happy you feel. |  |
| e1 | I am interested in your general level of well-being or satisfaction with life. How satisfied are you with your life, all things considered? | 1. Very satisfied 2. Somewhat satisfied 3. Neutral 4. Somewhat unsatisfied 5. Very unsatisfied |
| e2 | Do you think that you are more, equally or less satisfied than other persons your age and sex living in your village? | 1. More satisfied 2. Equally satisfied 3. Less satisfied |
| e3 | In general, would you say your health now is: very good, good, poor or very poor? | 1. Very good 2. Good 3. Poor 4. Very poor |
| e4 | How would you compare your health to other people of the same age and sex in your village? | 1. More healthy 2. Equally healthy 3. Less healthy |
| e5 | In the past month, how many days were you too sick to work/go to school/complete household chores? | ___________________ |
| Happiness | | |
| e6 | How true are the following statements for you in the last month? |  |
| e6_1 | I have felt depressed | 1. Strongly Agree 2. Agree 3. Disagree 4. Strongly Disagree |
| e6_2 | I have felt life was not worth living | 1. Strongly Agree 2. Agree 3. Disagree 4. Strongly Disagree |
| e6_3 | I have felt content. | 1. Strongly Agree 2. Agree 3. Disagree 4. Strongly Disagree |
| e6_4 | I have felt lonely | 1. Strongly Agree 2. Agree 3. Disagree 4. Strongly Disagree |
| GENDER EQUITABLE MEN SCALE | | |
| Note | Please tell me if you strongly agree, agree, disagree, or strongly disagree with the following statements: |  |
| j1 | Woman’s most important role is to take care of her home and cook (take care of home is about housekeeping) | 1. Strongly Agree 2. Agree 3. Unsure 4. Disagree 5. Strongly Disagree   88. Refuse to say |
| j2 | Men need sex more than women | 1. Strongly Agree 2. Agree 3. Unsure 4. Disagree 5. Strongly Disagree   88. Refuse to say |
| j3 | Men don’t talk about sex, they just do it. | 1. Strongly Agree 2. Agree 3. Unsure 4. Disagree 5. Strongly Disagree   88. Refuse to say |
| j4 | There are times when a woman deserves to be beaten | 1. Strongly Agree 2. Agree 3. Unsure 4. Disagree 5. Strongly Disagree   88. Refuse to say |
| j5 | Changing diapers, giving kids a bath & feeding kids are mother’s responsibility | 1. Strongly Agree 2. Agree 3. Unsure 4. Disagree 5. Strongly Disagree   88. Refuse to say |
| j6 | It is a woman’s responsibility to avoid getting pregnant | 1. Strongly Agree 2. Agree 3. Unsure 4. Disagree 5. Strongly Disagree   88. Refuse to say |
| j7 | A man should have the final word about decisions in his home | 1. Strongly Agree 2. Agree 3. Unsure 4. Disagree 5. Strongly Disagree   88. Refuse to say |
| j8 | Men are always ready to have sex | 1. Strongly Agree 2. Agree 3. Unsure 4. Disagree 5. Strongly Disagree   88. Refuse to say |
| j9 | A woman should tolerate violence in order to keep her family together | 1. Strongly Agree 2. Agree 3. Unsure 4. Disagree 5. Strongly Disagree   88. Refuse to say |
| j10a | I would be outraged if my wife asked me to use a condom. | 1. Strongly Agree 2. Agree 3. Unsure 4. Disagree 5. Strongly Disagree   88. Refuse to say |
| j10b | Men would be outraged if their wife asked them to use a condom | 1. Strongly Agree 2. Agree 3. Unsure 4. Disagree 5. Strongly Disagree   88. Refuse to say |
| j11 | A man and a woman should decide together what type of contraceptive to use | 1. Strongly Agree 2. Agree 3. Unsure 4. Disagree 5. Strongly Disagree   88. Refuse to say |
| j12 | I would never have a homosexual friend | 1. Strongly Agree 2. Agree 3. Unsure 4. Disagree 5. Strongly Disagree   88. Refuse to say |
| j13a | If someone insults me, I will defend my reputation, with force if I have to. | 1. Strongly Agree 2. Agree 3. Unsure 4. Disagree 5. Strongly Disagree   88. Refuse to say |
| j13b | If someone insults a man, he should defend his reputation, with force if he has to | 1. Strongly Agree 2. Agree 3. Unsure 4. Disagree 5. Strongly Disagree   88. Refuse to say |
| j14 | To be a man you need to be tough. | 1. Strongly Agree 2. Agree 3. Unsure 4. Disagree 5. Strongly Disagree   88. Refuse to say |
| j15 | Men should be embarrassed if unable to get an erection | 1. Strongly Agree 2. Agree 3. Unsure 4. Disagree 5. Strongly Disagree   88. Refuse to say |
| j16 | If a guy gets a woman pregnant, child is the responsibility of both the man and woman | 1. Strongly Agree 2. Agree 3. Unsure 4. Disagree 5. Strongly Disagree   88. Refuse to say |
| j17 | A man should know what his partner likes during sex | 1. Strongly Agree 2. Agree 3. Unsure 4. Disagree 5. Strongly Disagree   88. Refuse to say |
| j18 | The participation of the father is important in raising children | 1. Strongly Agree 2. Agree 3. Unsure 4. Disagree 5. Strongly Disagree   88. Refuse to say |
| j19 | It’s important for men to have friends to talk about their problems | 1. Strongly Agree 2. Agree 3. Unsure 4. Disagree 5. Strongly Disagree   88. Refuse to say |
| j20 | A couple should decide together if they want to have children. | 1. Strongly Agree 2. Agree 3. Unsure 4. Disagree 5. Strongly Disagree   88. Refuse to say |
| Comment | We have reached the end of the chat. Thank you for your time. Do you have anything else you would like to say? |  |
| End Note | Thank the participant for their time and give them transport reimbursement, if they did not come for an ART appointment |  |
| Comments | Enumerator comments |  |
| End of the survey! | | |

#### 14.7. APPENDIX G: Follow-up Survey - Male

**FOLLOW-UP SURVEY**

**Identifying efficient linkages strategies for HIVST (IDEaL)**
Male

**Complete this form for men who enrolled in the study 4-months ago**

**Date of Interview: _______________________
Site Code: ______________________________**

**Full Name of Interviewer: __________________________________**

**Participant Study ID#: _____________**

| **#** | **Question** | **Response** |
| --- | --- | --- |
| c1a | Please think about your primary partner in the last 4 months.  Has your relationship changed in the last 4 months? How? | 🞏 Nothing changed (1)  🞏 Married (2)  🞏 Steady Girlfriend (3)  🞏 Moved out of the house (4)  🞏 Became an infrequent  partner (5)  🞏 Separated (6)  🞏 Divorced (7)  🞏 Other (8) |
| c4a | Have you disclosed your HIV status to this partner? | 🞏 Yes (1)  🞏 No (0) |
| c5a | Have you had any new children with this partner since we last spoke? | 🞏 Yes (1)  🞏 No (0) |
| c18a | Have you started ART in the past 4months? | 🞏 Yes (1)  🞏 No (0)  🞏 No, but I plan to link |
| c18b | IF YES: When did you start ART? | Day/Month/Year |
| **Unintended Consequences** | | |
| c19a | Were unwantedly pressured to initiate ART? | 🞏 Yes (1)  🞏 No (0) |
| c20a | After enrolling in the study, did anyone find out your HIV status against your will (unwanted disclosure)? | 🞏 Yes (1)  🞏 No (0) |
| C20b | After enrolling in the study, did anyone find out your partners’ HIV status against her will (unwanted disclosure)? | 🞏 Yes (1)  🞏 No (0) |
| c21a | After enrolling in the study, did your partner….  Threaten to hurt or harm you or someone you cared about. | 🞏 Yes (1)  🞏 No (0)  🞏 Refused to respond (99) |
| c22a | Insulted you or made you feel bad about yourself. | 🞏 Yes (1)  🞏 No (0)  🞏 Refused to respond (99) |
| c23a | Hit, slapped, kicked or did anything else meant to physically hurt you. | 🞏 Yes (1)  🞏 No (0)  🞏 Refused to respond (99) |
| c21a | After enrolling in the study, did you ever do the following to your partner….  Threaten to hurt or harm her or someone she cared about. | 🞏 Yes (1)  🞏 No (0)  🞏 Refused to respond (99) |
| c22a | Insulted her or made her feel bad about herself. | 🞏 Yes (1)  🞏 No (0)  🞏 Refused to respond (99) |
| c23a | Hit, slapped, kicked or did anything else meant to physically hurt her. | 🞏 Yes (1)  🞏 No (0)  🞏 Refused to respond (99) |
| c24 | Slept with another woman | 🞏 Yes (1)  🞏 No (0)  🞏 Refused to respond (99) |
| c25a | *Now, I am going to ask you a series of questions about who makes within this relationship. Please think about the last 4 months*  Who usually decides how the money you earn will be used? | 🞏 Yourself (respondent)  🞏 Jointly (This partner and you together)  🞏 Mainly this partner  🞏 Someone else  🞏 Do not earn money  🞏 Refuse to say |
| c26a | Who usually decides how your partner’s earnings will be used? | 🞏 Yourself (respondent)  🞏 Jointly (This partner and you together)  🞏 Mainly this partner  🞏 Someone else  🞏 Refuse to say |
| c27a | Who usually makes decisions about health care for yourself? | 🞏 Yourself (respondent)  🞏 Jointly (This partner and you together)  🞏 Mainly this partner  🞏 Someone else  🞏 Refuse to say |

|  | **D. Additional Questions** | |
| --- | --- | --- |
| **#** | **Question** | **Response** |
| d1 | Would you recommend the ART intervention you were part of to other male friends or family? | 🞏 Yes (1)  🞏 No (0) |
| d2 | Are you happy that you participated in the ART intervention? | 🞏 Yes (1)  🞏 No (0) |

Thank the participant for their time and end the survey

#### 14.8. APPENDIX H: Follow-up Survey – Female

**FOLLOW-UP SURVEY**

**Identifying efficient linkages strategies for HIVST (IDEaL)**
Female

**Complete this form for women whose:**

**(1) Partners consented to be in the study 4-months ago**

**Date of Interview: _______________________
Site Code: _____________________________**

**Full Name of Interviewer: __________________________________**

**Participant Study ID#: _____________**

| **#** | **Question** | **Response** |
| --- | --- | --- |
| c1a | Please think about your primary partner in the last 4 months.  Has your relationship changed in the last 4 months? How? | 🞏 Nothing changed (1)  🞏 Married (2)  🞏 Steady Boyfriend (3)  🞏 Moved out of the house (4)  🞏 Became an infrequent partner (5)  🞏 Separated (6)  🞏 Divorced (7)  🞏 Other (8) |
| c4a | Have you disclosed your HIV status to this partner? | 🞏 Yes (1)  🞏 No (0) |
| c5a | Have you had any new children with this partner since we last spoke? | 🞏 Yes (1)  🞏 No (0) |
| c18a | To your knowledge, did your partner start ART? | 🞏 Yes (1)  🞏 No, they do not plan to link (2)  🞏 No, but they plan to link (3)  🞏 Unsure (88) |
| **Unintended Consequences** | | |
| c19a | Did you pressure your partner to initiate ART? | 🞏 Yes (1)  🞏 No (0) |
| c20a | After enrolling in the study, did anyone find out your HIV status against your will (unwanted disclosure)? | 🞏 Yes (1)  🞏 No (0) |
| C20b | After enrolling in the study, did anyone find out your partners’ HIV status against his will (unwanted disclosure)? | 🞏 Yes (1)  🞏 No (0) |
| c21a | After enrolling in the study, did your partner….  Threaten to hurt or harm you or someone you cared about. | 🞏 Yes (1)  🞏 No (0)  🞏 Refused to respond (99) |
| c22a | Insulted you or made you feel bad about yourself. | 🞏 Yes (1)  🞏 No (0)  🞏 Refused to respond (99) |
| c23a | Hit, slapped, kicked or did anything else meant to physically hurt you. | 🞏 Yes (1)  🞏 No (0)  🞏 Refused to respond (99) |
| C23b | Forced sexual intercourse and other forms of sexual coercion | 🞏 Yes (1)  🞏 No (0)  🞏 Refused to respond (99) |
| C23c | Slept with another woman. | 🞏 Yes (1)  🞏 No (0)  🞏 Refused to respond (99) |
| c24a | Ended the relationship | 🞏 Yes (1)  🞏 No (0)  🞏 Refused to respond (99) |
| c25a | *Now, I am going to ask you a series of questions about who makes within this relationship*  Who usually decides how the money you earn will be used? | 🞏 Yourself (respondent)  🞏 Jointly (This partner and you together)  🞏 Mainly this partner  🞏 Someone else  🞏 Do not earn money  🞏 Refuse to say |
| c26a | Who usually decides how your partner’s earnings will be used? | 🞏 Yourself (respondent)  🞏 Jointly (This partner and you together)  🞏 Mainly this partner  🞏 Someone else  🞏 Refuse to say |
| c27a | Who usually makes decisions about health care for yourself? | 🞏 Yourself (respondent)  🞏 Jointly (This partner and you together)  🞏 Mainly this partner  🞏 Someone else  🞏 Refuse to say |

|  | **Additional Questions** | |
| --- | --- | --- |
| d1 | Would you recommend the ART intervention to other male friends or family? | 🞏 Yes (1)  🞏 No (0) |
| d2 | Are you happy your partner was in the ART intervention? | 🞏 Yes (1)  🞏 No (0) |

Thank the participant for their time and end the survey

#### 14.9. APPENDIX I: Data Extraction Tool

**DATA EXTRACTION TOOL**

**Identifying efficient linkages strategies for HIVST (IDEaL)**

English only

**INSTRUCTIONS:**

The Medical Chart Review will be used to link the male study participant with the facilities ART records and to document their facility visits over the 4-months of study participation. Please follow the instructions to prepare for data collection (1) gather all ART registers that were active between DAY MONTH YEAR up to today (2) enter and re-enter the participant ID’s who have reached the 4-month follow up period into the tablet (3) once you have re-entered, the tablet will provide you with identifying information about the male study participant. (5) match the participants information with information provided by the ART register to see if the participant initiated ART or not. If a participant did not initiate care (i.e. you cannot find him in the ART register), still enter the initial data points and indicate that the participant did not

| **Code** | **Question** | **Responses** |
| --- | --- | --- |
| pid | Please enter the Participant ID |  |
| district | District | ☐ Chickwawa  ☐ Nkhotakota  ☐ Lilongwe  ☐ Kasungu |
| site | Facility name | ☐ Chickwawa District Hospital  ☐ St. Montford Mission Hospital  ☐ Kalemba Community Hospital  ☐ Kasungu District Hospital  ☐ Nkhoma Community Hospital  ☐ Mponela Rural Hospital  ☐ Deayang Luke Hospital  ☐ Nkhotakota District Hospital  ☐ Nsanje District Hospital  ☐ Ngabu Rural Hospital |
|  | *We want to know if this participant initiated ART. Please look at the ART register used at the clinic between DATE MONTH YEAR up to today*  *Instructions: Look for the below information in the ART register, matching the below participant with a name in the ART register.*  *Sometimes it is hard to find an exact match in the ART register. Consider it a match if 3 of the 4 data points match. For example, someone’s name may be different, but the age, and village/residence matches. Consider this the same person.*  *CLIENT NAME:*  *AGE:*  *Ta:*  *Village:* |  |
| found_art | Was the participant found in the ART register? | ☐ Yes (1) – proceed to next question  ☐ No (0) – end survey |
| art_number | What is the participant assigned ART number? |  |
| art_date | What is the clients ART start date? | ___ ___/___ ___/___ ___  Day Month Year |
|  | *Use the ART number to find the paper Mastercard OR look up the participant in the Baobab system* |  |
| mastercard_found | Did you find the mastercard/baobab record? | ☐ Yes (1) – proceed to next question  ☐ No (0) – end survey |
| tb | At initiation: TB | ☐ Yes (1)  ☐ No (0) |
| ks | At initiation: KS | ☐ Yes (1)  ☐ No (0) |
| pillcount | Number of pills given |  |
| nextapp_date | Date of next appointment | ___ ___/___ ___/___ ___  Day Month Year |
| nextconsult_date | Date of next consultation visit | ___ ___/___ ___/___ ___  Day Month Year |

**END OF SURVEY**

#### 14.10. APPENDIX J: In-Depth Interview Guide – Female

**IN-DEPTH INTERVIEW GUIDE**

**Identifying efficient linkages strategies for HIVST (IDEaL)**
Female

**BEGIN RECORDING**

Original Study ID: ______________________

Repeat Original Study ID:

State if male or female respondent _______________________

Health Facility: _______________________

Date of Interview: _______________________

Full Name of Interviewer: __________________________________

District where respondent Lives: __________________________________

**Open- Ended Questions**

*Note: The in-depth interview will be open-ended and guided by the respondent’s answers. This outline reflects a general guide for the in-depth interviews.*

*The interviews are meant to help us understand barriers and facilitators to ART initiation. We are also interested in their thoughts on new interventions we are developing to help men start ART.* *The following questions are meant to guide interviewers. Actual questions asked during the interview will vary based on participant responses.*

**DEMOGRAPHICS:**

Demographics on the female participant not collected. Already in the original study. Just make sure the Original Study ID is documented correctly.

1. What type of job/work does your partner do?
   a. If he does not work, why?
   b. Does he do anything else to earn money?
2. What time of day/week is your partner usually busy?
3. Where does he spend most of his time when he is not working?
   a. *Probe: At the bar, watching football, at church, at home…*
4. How long have you been in a relationship with your sexual partner (whom you gave the HIVST kit to)

**HIV TESTING:**

Now I’d like to talk to you about you and your partner’s experience with HIV services

1) To your knowledge, has your partner tested for HIV before you gave him the HIV-self testing kit?
   a) IF YES, did he tell you the result? What was his result?
   b) IF HIV+: At that time, did he start ART? IF NO: Why not?
2) Think about when you gave the HIV self-test kit to your sexual partner (show the kit).
   a) How did your partner feel about his HIV positive test result? How did the result affect him?
   b) How did you feel after your partner received an HIV positive status on the HIV self-test kit? How did the result affect you?
   c) Did your relationship change at all after he received an HIV-positive status? How?
   d) Did he talk to anyone about it?/ who?
   e) Did he disclose to anyone besides you? Why/why not?

**ART UPTAKE:**

1) After he tested HIV-positive, did he initiate ART? Why/Why not?
   a) IF INITIATED ART: How long did it take him to initiate ART? (several weeks, several months?). Why did you initiate ART so quickly/slowly? /

We know starting ART is difficult.

1) What do you think is the most difficult thing about starting ART for your partner right now?
   a) Do you think this is the same for men and for women? How is it different?
   b) How does taking ART affect one’s daily activities? (schedule/routine) In bad ways? In good ways? Is it different for men? How?
   c) How does taking ART affect relationships? In bad ways? In good ways? Is it different for men? How?
   d) Out of all these things you mentioned, what do you think is the biggest concern/problem for your partner?

FOR MEN WHO ARE CONSIDERED LOSS-TO-FOLLOW UP

1. Since we saw you last, we haven’t been able to follow up with your partner. From your perspective, why do you think this is?
2. Since your partner was enrolled in our study, what kind of things did you notice or did he mention to you related to HIV health services?
   a. *Probe: phone calls/SMS or speak one-on-one to a health care worker? Was your partner visited by a health care professional within your home or in the community?*
3. Has your partner mentioned any of these interactions to anyone?
   a. *Probe: friends, family, community members*
4. How did your partner react to these things? Did he like/dislike them? What did he do when they happened?
   a. *Probe: phone call, SMS, in-person visit, home-based ART*
5. How did you react to these things? Did you like/dislike them?
   a. *Probe: Was there any reaction amongst your family, friends or the community to these things?*
   b. *If so, how did this affect your partner and/or you?*

**SUGGESTIONS FOR ART SERVICES**

Thank you for all the information. We would like to develop ART services that meet the needs of individuals in your community. We understand that men may face different challenges than women. I would like to know your opinion about what is needed in order to make ART services easy to access and use.

1) Could you describe the ideal way ART services would be given to your partner? If the clinic could do anything …
   a) When would he want to pick up ART?
      i) Probe: day of week, time of day?
      ii) *Why do you say this?*
   b) Where would he want to pick up ART*?*
      i) Probe: clinic near you, clinic far from you, somewhere in the community (WHERE SPECIFICALLY), at your home?
      ii) *Why do you say this?*
2) Do you think your partner needs more HIV-related information? About the benefits of ART, how to keep his status a secret, or how to disclose his status, about other aspects of his health?
   a) IF YES: How would he want to get this information? In person (one on one, pamphlet, radio, phone call, …)
      i) Probe: IF YES: WHO would he like to talk to about this information? (Provider, expert client, other community member)
3) Does he need support? For example, reminders to go to the health facility, someone to talk to regularly about what he is going through with ART, help disclosing his status, anything else).
   a) *Probe: IF YES: How would he want to get this support? In person (one on one, pamphlet, radio, phone call)*
   b) *Probe: IF YES: WHO would he like to talk to about this support? (Provider, expert client, other community member)*
4) Out of time of pick up, location, more information, or more support, what are the most important factors for your partner to use ART services?
   a) *PROBE: Think about other males. What do you think they would say is the most important factor for men to use ART services?*

**SPECIFIC SUGGESTIONS ON CURRENT INTERVENTIONS**

FOR MEN WHO ARE CONSIDERED LOSS-TO-FOLLOW UP

1. As discussed earlier, your partner received [phone calls, texts, in person counseling, home-based ART].
   a. *Do you feel that this was enough to encourage him to start ART?*
      i. *If NO, what would you have done differently? (frequency, content, location)*
      ii. *If YES, why do you think they were sufficient?*
   b. Are there any other ideas/services we should think about doing beside the ones we just talked about (appointment reminders, in-depth counseling, community/home ART)?/
      i. PROBE: What is it?
      ii. Why do you think this could work?

**CONCLUSION**

Thank you for your time.

1. Is there anything else you would like to say about men’s use of ART?
2. Is there anything else you would like to say about your own use of ART and how we can help make your experience better?

**STOP THE RECORDER AND MAKE SURE RECORDING IS SAVED.**

**THANK YOU FOR YOUR PARTICIPATION IN THIS INTERVIEW. LET ME ENCOURAGE YOU THAT ARVS CAN HELP YOU LIVE LONG AND HEALTHY.**

*The following general education messages should be conveyed to all male and female participants:*

- All people who have been tested HIV positive should start ART as soon as possible for their own health and to prevent passing the virus on to others.
- Serious diseases can occur even in patients with high CD4 count (>500), without any previous symptoms. Immediate ART greatly reduces this risk.
- People that start ART and continue lifelong without interruptions can remain healthy and live as long as people without HIV.
- Even though you may not feel sick, ART is still important to keep you healthy for the rest of your life.
- ART reduces the amount of virus in your body and therefore can reduce the chance that HIV is passed to your sex partners.
- Current ART regimens are easy to take and rarely cause serious side-effects. Some people have side effects in the first few weeks of treatment and these almost always go away. IF there are persistent side effects, an alternative HIV regimen can be given.

*NOTE: Be careful not to give any specific medical advice but rather refer respondents back to the clinic to speak to a provider.*

#### 14.11. APPENDIX K: In-Depth Interview Guide – Male

**IN-DEPTH INTERVIEW GUIDE**

**Identifying efficient linkages strategies for HIVST (IDEaL)**
Male

**BEGIN RECORDING**

Original Study ID: ______________________

Repeat Original Study ID:

State if male or female respondent _______________________

Health Facility: _______________________

Date of Interview: _______________________

Full Name of Interviewer: __________________________________

District where respondent Lives: __________________________________

**DEMOGRAPHICS:**

| 1. What is your current age in years? | Age in Completed Years |
| --- | --- |
| 2. How would you rate your health today on a scale from 1-5 with 1 being excellent health and 5 being the very poor health? | 🞏 Excellent (1)  🞏 Very good (2)  🞏 Good (3)  🞏 Fair (4)  🞏 Poor (5) |
| 3. Now I’d like to ask about your relationships. Are you currently in a sexual relationship? | 🞏 Yes (1) 🞏 No (0)  **If NO, skip to QUALITATIVE** |
| 4. Does your partner know your HIV status? | 🞏 Yes (1)  🞏 No (0) |
| 5. How many times have you tested for HIV? |  |
| 6. When was the first time you tested HIV-positive? |  |

**Open- Ended Questions**

*Note: The in-depth interview will be open-ended and guided by the respondent’s answers. This outline reflects a general guide for the in-depth interviews.*

*The interviews are meant to help us understand barriers and facilitators to ART initiation. We are also interested in their thoughts on new interventions we are developing to help men start ART.* *The following questions are meant to guide interviewers. Actual questions asked during the interview will vary based on participant responses.*

SCRIPT: Now I’d like to talk to you about your experience with HIV services.

**HIV TESTING**

1. Think about when you used an HIV self-test kit (show the kit). When did you use it?
2. How did you feel after receiving the HIV positive status?
3. Can you talk to me about what happened after you tested HIV positive? Walk me through it so I can see the picture in detail?.
   a. Did you talk to anyone about it?
   b. Has your HIV status changed your daily activities at all? (Your schedule/routine)
   c. Has your HIV status impacted your relationships? How?
   d. Have you disclosed your status to anyone besides your partner? Why/why not

**INTERVENTION**

1. After you tested positive, we approached you to be a part of our study. Since you were enrolled in the study (ie in the last 3 months), can you walk me through what has happened related to HIV health services?
   a. Probe: What kinds of interactions have you had with expert clients or health personelle (phone call, SMS, in-person visits, home based ART).
   b. What was the frequency of these interactions (weekly, every other week, monthly)

**ART INITIATION**

2. Since you have been enrolled in the study, have you initiated ART?

INITIATED

3. How long did it take you to initiate ART? (several weeks, several months?). Where did you initiate?
   a. Why did you initiate ART so quickly/slowly?
4. Since you initiated treatment, have you continued to take you medication?
   a. Have you returned to the clinic for another refill of ART? Have there been challenges to staying on treatment – how have you overcome them?

NOT INITIATED

1) Since you tested HIV-positive, have you been to a health facility?
   a) When did you attend? (year)
   b) Why did you attend? (guardian vs client; HIV vs OPD)
   c) Why did you not initiate ART during this visit?

**ART UPTAKE**

2) We know starting ART is difficult. What do you think is most the most difficult thing about starting ART?
   a) PROBE: Think about your male friends. What do you think they would say is the most difficult part about starting ART for men in your village?

INITITATED ART

1) How has taking ART affected your daily activities at all? (your schedule/routine) In bad ways? In good ways?
2) How has taking ART affected your relationships? In bad ways? In good ways?
3) Does being on ART change if you are able to hide/keep your HIV status from other people/ If you are able to hide/keep your status other people, does that make it easier to be on ART? How?

NOT INITITATED ART

1) How do you think taking ART would affect your daily activities at all? (your schedule/routine) In bad ways? In good ways?
2) How do you think taking ART would affect your relationships? In bad ways? In good ways?
3) Would being able to hide/keep your HIV status from other people make it easier for you to be on ART? How?
4) What do you think is most the SECOND most difficult thing about starting ART?
   a) PROBE: Think about your male friends. What do you think they would say is the SECOND most difficult part about starting ART for men in your village?

**SUGGESTIONS FOR ART SERVICES**

Thank you for all the information. We would like to develop ART services that meet the needs of men in your community. We understand that men are busy and may face different challenges than women. I would like to know your opinion about what is needed in order to make ART services easy to use for men in your community

1) Could you describe the ideal way ART services would be given to you? If the clinic could do anything
   a) When would you want to pick up ART?
   b) Probe: day of week, time of day?
   c) Why do you say this?
2) Where would you want to pick up ART?
   a) Clinic near you, clinic far from you, somewhere in the community (WHERE SPECIFICALLY), at your home?
   b) Why do you say this?

I would like to learn about how you felt about each of the interactions we talked about earlier (remind participant of what they mentioned – ex: phone calles, texts, in person visits, home based ART)

1) What did you like about them?
   a) Why? (ex: individual follow-up, sense of support, not having to travel to the clinic for homebased ART)
2) What did you dislike about them? What were challenges?
   a) Why? (ex: difficulty maintaining privacy with contact or visits, doesn’t want to start ART for other reasons)
3) Do you feel like these things helped encourage you to seek health services?
   a) If YES, why?
   b) If NO, why?
4) Do you think these things would help other men in your community if they were to test positive for HIV?

INITIATED

1) Do you feel that these things helped to encourage you to initiate and stay on ART?
2) Do you think you would have started treatment without them?

NOT INITIATED

1) Why do you think these things failed to help you start/stay on treatment?
   a) Why?
2) If you could change anything about these interactions that you have listed, what would you change?
   a) Probe: Type of contact, frequency of contact, personelle, location, topics covered
3) We understand that everyone is different. Beyond what you have experienced, do you still have problems related to seeking health services for HIV? (i.e. are there still things that you need?)
   a) What are these unmet needs?
   b) What do you feel would be the best solution to meet those needs?
4) Are there any other ideas/services we should think about doing beside the ones we just talked about (appointment reminders, in-depth counseling, community/home ART)?
   a) What is it?
   b) Why do you think this could work?
5) Is there anything else that you would like to add as we are towards the end of the interview?

**STOP THE RECORDER AND MAKE SURE RECORDING IS SAVED.**

**THANK YOU FOR YOUR PARTICIPATION IN THIS INTERVIEW. LET ME ENCOURAGE YOU THAT ARVS CAN HELP YOU LIVE LONG AND HEALTHY.**

*The following general education messages should be conveyed to all male and female participants:*

- All people who have been tested HIV positive should start ART as soon as possible for their own health and to prevent passing the virus on to others.
- Serious diseases can occur even in patients with high CD4 count (>500), without any previous symptoms. Immediate ART greatly reduces this risk.
- People that start ART and continue lifelong without interruptions can remain healthy and live as long as people without HIV.
- Even though you may not feel sick, ART is still important to keep you healthy for the rest of your life.
- ART reduces the amount of virus in your body and therefore can reduce the chance that HIV is passed to your sex partners.
- Current ART regimens are easy to take and rarely cause serious side-effects. Some people have side effects in the first few weeks of treatment and these almost always go away. IF there are persistent side effects, an alternative HIV regimen can be given.

*Be careful not to give any specific medical advice but rather refer respondents back to the clinic to speak to a provider.*

APPENDIX L: Personel CV’s

Kathryn L. Dovel

Department of Medicine, Division of Infectious Diseases

Los Angeles, CA 90095

POSITIONS HELD

2017-	Adjunct Assistant Professor, Division of Infectious Disease
 Department of Medicine, David Geffen School of Medicine - UCLA

2017-	Research Director - Partners in Hope

Lilongwe, Malawi

2016-17	Postdoctoral Fellow in Global HIV Prevention Research

David Geffen School of Medicine - UCLA, 2016

2012-15	International Programs Director - 31Bits International

Gulu, Uganda

EDUCATION

PhD	Health and Behavioral Sciences - University of Colorado Denver, 2016

*Outstanding CLAS Ph.D. Student for the University of Colorado Denver*

*Outstanding Dissertation Award for the University of Colorado Denver*

Dissertation: “Shifting Focus from Individuals to Institutions: The Role of Gendered Health Institutions on Men’s use of HIV Services”

MPH	Community Health Sciences - UCLA, 2010

*Certificate in Global Health*

BA	Sociology & Anthropology (dual major), minor in Biology - Vanguard University, 2007

*Magna Cum Laude*

**HONORS AND AWARDS**

2019	5th place in the 2019 Department of Medicine Research Day poster competition, Department of Medicine, University of California Los Angeles

2019	Invited panelist, IAS 2019, Sticky and durable linkage: Latest evidence and new strategies

2019	Panel Chair, IAS Pre-Conference 2019, Men and HIV: What we know and what we don’t know

2019	Top 5 abstracts at CROI, 2019

2019	Invited participant, Technical Consultation on HIV Linkage, International AIDS Society

2019	Invited participant, Differentiated Service Delivery Think Tank, Gates Foundation

2018	Joep Lange Award (best abstract at INTEREST, 2018)

2018	Female Global Scholar, The Women in Global Health Research Initiative (Weill Cornell Medicine, Cornell University)

2016	Outstanding Dissertation Award (UCDenver)

2016	Outstanding CLAS Ph.D. Student  (UCDenver)

2012-16	Deans Travel Grant (UCDenver)

2007	Delta Kappa Honor Society (Vanguard University)

2007	Alpha Kappa Delta Honor Society (Vanguard University)

2007	Lambda Alpha Honor Society (Vanguard University)

2007	Anthropology Student of the Year (Vanguard University)

**EXTERNAL GRANTS**

2019-2023 Principle Investigator, Bill and Melinda Gates Foundation. (001423) “Identifying Effective Linkage Strategies for HIVST (IDEaL)”

2019-2024 Principle Investigator, Fogarty International Center. International Research Scientist Development Award (K01), K01TW011484. “Innovative strategies to increase ART

Initiation and viral suppression among HIV+ men in Malawi”.

2019-2020 Principle Investigator, Clinton Health Access Foundation. “The impact of facility HIV self-test scale up in Malawi: a mixed methods study”

2018-2021	Co-Investigator, The Conrad N. Hilton Foundation. Delivery of childhood development services as part of HIV treatment services in Malawi. Project implemented by UCLA and Partners in Hope.

2017-19	Principal Investigator, USAID. **Use of HIV self-test kits to increase identification of HIV-infected individuals and their partners: a Cluster Randomized Control Trial.** (sub-study within a large PEPFAR-USAID grant; PI: Risa Hoffman).

2017-19	Principle Investigator, Clinical Research Scholar, National Institutes of Health Loan Repayment Program.

2016-18	Co-Principle Investigaor, USAID. Test and Start: Tracking Uptake and Retention in Care using Standard Registry Data. (sub-study within a large PEPFAR-USAID grant; PI: Risa Hoffman).

2015	Principal Investigator, 31Bits International. “Evaluating the impact of a couple’s livelihoods program on power dynamics and economic attainment among couples in northern Uganda.” Project implemented by 31Bits International.

2014-16	Principal Investigator, NIMH National Research Service Award Predoctoral Individual Fellowship, F31-MH103078-01A1, “Gender Disparities in High-Risk PITC: The Role of Policy on Provider Practices”, Impact Score: 14; Percentile: 2.0

2013-16	Principal Investigator, Stop AIDS Now!. “Evaluation of the ‘Quality HIV and reproductive maternal and neonatal health services for women and young women in Africa through good clinical governance and community-driven accountability’”. Project implemented by the Clinton Health Access Initiative.

**INTERNAL GRANTS**

2016-18	Principal Investigator, UCLA Center for AIDS Research Seed Grant, University of California Los Angeles, “The Gendered Dynamics of ART Uptake and Retention under Universal Treatment Policies. Examining trends and ART barriers in Central Malawi”

2014	Principal Investigator, Calvin L Wilson Scholarship, University of Colorado Denver, “Gender and the provision of HIV testing: Examining how models of care influence men’s use of testing services in southern Malawi”

2013-15	Principal Investigator, Dissertation Grant, University of Colorado Denver, “Gender Disparities in High-Risk PITC: The Role of Policy on Provider Practices”

2013-14	Principal Investigator, Robinson Durst Scholarship, University of Colorado Denver, “Gender disparities in high-risk PITC: Exploring the influence of feminized policy on provider practices in Malawi”

2009	Principal Investigator, Drabkin and Bixby International Scholarship, UCLA, “Evaluating barriers and facilitators of a nutrition program in the Bateyes of Dominican Republic”

2009	Principal Investigator, Global Health Grant, UCLA, “Evaluating a Nutrition Program in the Bateyes of Dominican Republic”

**PUBLICATIONS**

***** represents MPH, PhD or medical students I mentored

2020	Cornell, Morna, Katherine Horton, Christopher Colvin, Andrew Medina-Marino, **Kathryn Dovel.** Raising the profile of men’s health: the role of the research community: Letter to the editor. *Lancet*. Ahead of Print.

2020	**Dovel, Kathryn**, Mike Nyirenda, Frackson Shaba*, O. Agatha Offorjebe*, Kelvin Balakasi, Brooke Nichols, Khumbo Phiri*, Khumbo Ngona, Sundeep K Gupta, Risa Hoffman. “Facility-based HIV self-testing for outpatients dramatically increases HIV testing in Malawi: a cluster randomized trial.” *Lancet Global Health*. Ahead of print

2020	**Dovel, Kathryn,** Khumbo Phii, Misheck Mphande, Deborah Mindry, Esnart Sanudi, McDaphton Bellos, Risa Hoffman. Optimizing Test and Treat in Malawi: Health care worker perspectives on barriers and facilitators to ART initiation among healthy clients. Global Health Action. Ahead of Print.

2020	Hubbard, Julie, Khumbo Phiri*, Corrina Moucheraud, Kaitlyn McBride, Ashley Bardon, Kelvin Balakasi, Eric Lungu, **Kathryn Dovel**, Gift Kakwesa, Risa Hoffman. A qualitative assessment of provider and client experiences with three- and six-month dispensing of antiretroviral therapy in Malawi. Global Health: Science and Practice. Ahead of Print.

2020	Hoffman, Risa M, Kelvin Balakasi, Ashley Bardon, Zumbe Siwale, Julie Hubbard, Gift Kakwesa, Mwiza Haambokoma, Thoko Kalua, Pedro Pisa, Crispin Moyo, **Kathryn Dovel**, Thembi Xulu, Ian Sanne, Matt Fox, Sydney Rosen. Eligibility for differentiated models of HIV treatment service delivery: an estimate from Malawi and Zambia. *AIDS.* 1;34(3):475-9.

2019	McBride, Kaitlyn, Julie Parent, Kondwani Mmanga, Mackenzie Chivwala, Mike H. Nyirenda, Alan Schooley, James B. Mwambene, **Kathryn Dovel**, Eric Lungu, Kelvin Balakasi, Risa M. Hoffman, Corrina Moucheraud. "ART Adherence Among Malawian Youth Enrolled in Teen Clubs: A Retrospective Chart Review." *AIDS Behav.* (2019): 1-5.

2019	Frackson Shaba*, Ogechukwu Offorjebe*, Phiri Khumbo, Lungu Eric, Kalande Pericles, Nyirenda Mike, Hoffman M Risa, Gupta Sundeep, **Dovel Kathryn**. Perceived Acceptability of a Facility-Based HIV Self-Test Intervention in Outpatient Waiting Spaces Among Adult Outpatients in Malawi: A Formative Study. *JAIDS*. 1;81(3):e92-4.

2019	Magaço Amílcar, **Dovel Kathryn**, Cataldo Fabian, Nhassengo Pedroso, Nuera Lucas, Tique José, Saide Mohomed, Couto Aleny, Mbofana Francisco, Gudo E Eduardo, Cuco Rosa Marlene, Chicumbe Sérgio. “Good health as a barrier and facilitator to ART initiation: a qualitative study in the era of Test and Treat in Mozambique.” *Cult Health Sex. 11:1-5.*

2018	Cornell M, **Dovel K**. Reaching key adolescent populations. *Cur Opinion HIV AIDS.* 1;13(3):274-80.

2018	Sara, Yeatman, Stephanie Chamberlin*, **Kathryn Dovel.** Women’s (health) work: A population-based, cross-sectional study of gender differences in time spent seeking health care in Malawi. *PLoS ONE.* 13(12): e0209586

2018	Nhassengo, Pedroso Fabian Cataldo, Amílcar Magaço, Risa Hoffman, Lucas Nuera, José Tique, Mohomed Saide, Aleny Couto, Francisco Mbofana, Eduardo Gudo, Rosa Marlene Cuco, Sérgio Chicumbe, **Kathryn Dovel**. “Barriers and facilitators to the uptake of universal treatment in Mozambique: a qualitative study on patient and provider perceptions.” *PLoS ONE.* 13(12): e0205919

2018	Hubbard, Julie, Gift Kakwesa, Mike Nyirenda, James Mwambeneb, Ashley Bardona, Kelvin Balakasi, **Kathryn Dovel**, Thokozani Kaluac, Risa Hoffman. Towards the third 90: improving viral load testing with a simple quality improvement program in health facilities in Malawi. *International Public Health.* Ahead of print.

2018	**Dovel, Kathryn,** Frackson Shaba*, Ogechukwu Offorjebe*, Kelvin Balakasi, Khumbo Phiri*, Brooke Nichols, Chi-Hong Tseng, Ashley Bardon, Khumbo Ngona, Risa Hoffman. “Evaluating the integration of HIV self-testing into low-resource health systems: study protocol for a cluster randomized trial from EQUIP Innovations” *Trials.* 19:498.

2018	Moucheraud, Corrina, Dennis Chasweka, Mike Nyirenda, Alan Schooley, **Kathryn Dovel**, Risa Hoffman. “A simple screening tool may help identify high-risk children for targeted HIV testing in Malawian inpatient wards.” *JAIDS.* 79:352-7.

2018	Cornell Morna, **Dovel Kathryn**. “Reaching key adolescent populations.” *Current opinion in HIV and AIDS.* 13(3):274-80.

2016	**Dovel, Kathryn**, Sara Yeatman, Joep Vanoosterhout, Adrienne Chan, Alfred Matengeni, Megan Landes, Richard Bedell, and Sumeet Sodhi. “Trends in ART Initiation among Men and Non-Pregnant/Non-Breastfeeding Women before and after Option B+ in Southern Malawi.” *PLoS ONE*. (12): e0165025.

2016	Poulin, Michelle, **Kathryn Dovel** and Susan Watkins. “Men with money and the 'vulnerable women' client category in an AIDS epidemic.” *World Development*. 85; 16-30.

2016	**Dovel, Kathryn**, Susan Watkins, Sara Yeatman, and Michelle Poulin. "Prioritizing strategies to reduce AIDS-related mortality for men in sub-Saharan Africa: Author’s reply.” *AIDS*. 30(1); 158-9.

2015	**Dovel, Kathryn,** Sara Yeatman, Susan Watkins, and Michelle Poulin. “Men’s heightened risk of AIDS-related death: the legacy of gendered HIV testing and treatment strategies.” *AIDS.* 29; 1123–5.

2015	**Dovel, Kathryn** and Kallie Thomson. “Financial obligations and economic barriers to antiretroviral therapy experienced by HIV positive women participating in a job-creation program in northern Uganda.” *Culture, Health, and Sexuality*. 18(6).

2015	Krueger, Patrick, **Kathryn Dovel** and Justin Denney. “Democracy and self-rated health across 67 countries: A multilevel analysis.*”* *Social Science and Medicine.* 143; 137-44.

2013	Conroy, Amy, Sara Yeatman and **Kathryn Dovel**. “The social construction of HIV/AIDS during a time of evolving access to antiretroviral therapy in rural Malawi.” *Culture, Health and Sexuality.* 15(8); 924-37.

2012	Yeatman, Sara, **Kathryn Dovel**, Amy Conroy and Hazel Namadingo. “The predictors of HIV treatment optimism and its relationship with sexual risk behavior among a population-based sample of young adults in southern Malawi.” *AIDS Care.* 25(8);1018-25.

**TECHNICAL MANUSCRIPTS**

2019	Hopkins, John, Laura Pascoe, Dean Peacock and Kathryn Dovel. “Accelerating Men’s HIV service delivery and uptake in Eastern and Southern Africa UNAIDS Literature Review, Eastern and Southern Africa Regional Focus.” UNAIDS, Johannesburg, South Africa.

2018	Masina, Tobias, **Kathryn Dovel**, Reuben Mwenda on behalf of the Malawi Ministry of Health. “National Guidelines for HIV self-testing.” Malawi Ministry of Health Lilongwe Malawi.

2017	Pascoe, Laura, Dean Peacock and **Kathryn Dovel**. “To Get to Zero, We Must Also Get to Men – UNAIDS Literature Review, Eastern and Southern Africa Regional Focus.” UNAIDS, Geneva.

2016	Macharia, Faith, Job Akuno, Faith Wanji, Julius Nguku, **Kathryn Dovel**, Caroline Ngare, Fred Nyagah, and Daniel Mwisunji. “National Guidelines for Male Engagement in HIV Services.” Kenya Ministry of Health. Nairobi, Kenya.

2016	**Kathryn Dovel**, James Mkandawire, Susan Watkins, Nancy Mulauzi and Sydney Rodney Lungu. “Evaluation of the Good Clinical Governance Project: improving HIV and reproductive health services in Lilongwe, Malawi.” Stop AIDS Now!. Lilongwe, Malawi.

**OTHER PUBLICATIONS**

2019	Kathryn Dovel, Stephanie Chamberlin, Sara, Yeatman. Malawi’s Health System Puts Women First. This Isn’t Always a Good Thing. *The Conversation: Africa.* Published February 19, 2019. Found at https://theconversation.com/malawis-health-system-puts-women-first-this-isnt-always-a-good-thing-111277

2016	**Dovel, Kathryn**, Sara Yeatman, and Susan Watkins. **Dying from a treatable disease: HIV and the men we neglect. *Huffington Post.* Published February 23, 2016. Found at** http://www.huffingtonpost.com/the-conversation-africa/dying-from-a-treatable-di_b_9295620.html

**WORK IN PREPARATION**

**Dovel, Kathryn**. “The gendered organization of HIV services and men’s poor use of testing in southern Malawi: consequences of hegemonic masculinity within health institutions.” (Revise & Resubmit, JIAS)

Offorjebe , Ogechukwu*, Frackson Shaba*, Kelvin Balakasi, Mike Nyrienda, Risa Hoffman, **Kathryn** **Dovel**. “Partner-delivered HIV self-testing increases the perceived acceptability of index partner testing among HIV-positive clients in Malawi.” (Revise & Resubmit, PLoS ONE)

**Dovel, Kathryn**, Kelvin Balakasi, Khumbo Phiri*, Frackson Shaba*, O. Agatha Offorjebe*, Sundeep K Gupta, Vincent Wong, Eric Lungu, Brooke Nichols, Mike Nyirenda, Ngona K, Anteneh Worku, Risa Hoffman. ^“^A randomized trial on index HIV self-testing for sexual partners of ART clients in Malawi.” (Under Review)

Nichols, Brooke; Offorjebe, O. Agatha; Cele, Refiloe; Shaba , Frackson ; Balakasi, Kelvin; Chivwara, Mackenzie; Hoffman, Risa; Long, Lawrence; Rosen, Sydney; **Dovel, Kathryn**. “Economic evaluation of facility-based HIV self-testing among adult outpatients in Malawi. " (Under Review)

**Dovel, Kathryn,** Gladies Orobmi, Melanie Beagly*, Kallie Thomson. “Including men without sidelining women: the feasibility of male involvement within resource-strained gender equality programs in sub Saharan Africa.” (Under Review)

**Dovel, Kathryn** and Kallie Thomson. “Evaluating the impact of a couple’s livelihoods program on power dynamics and economic attainment among couples in northern Uganda.” (In preparation)

**Dovel, Kathryn**. “Men in global HIV policy: examining discourses of blame and vulnerability.” (In preparation)

**SELECT PEER-REVIEWED PRESENTATIONS**

2020	Moucheraud, Corrina, Samuel W. Lewis, Misheck Mphande, Ben Allan Banda, Hitler Sigauke, Paul Kawale, Aubrey Dkangoma, **Kathryn Dovel**, Alemayehu Amberbir, Agnes Moses, Sundeep Gupta, Risa M. Hoffman. Cervical cancer knowledge and attitudes among HIV-positive men in Malawi.” Paper accepted for poster presentation. Conference on Retroviruses and Opportunistic Infections (CROI). Boston, Massachusetts, USA

2019	**Dovel, Kathryn**, Kelvin Balakasi, Khumbo Phiri*, Frackson Shaba*, O. Agatha Offorjebe*, Sundeep K Gupta, Vincent Wong, Eric Lungu, Brooke Nichols, Mike Nyirenda, Ngona K, Anteneh Worku, Risa Hoffman. ^“^Index HIV self-testing among male partners in Malawi: predictors of self-testing within a randomized controlled trial”. Paper accepted for poster presentation. International AIDS Society. Mexico City, Mexico

2019	**Dovel Kathryn**, Salem Ejigu, Pericles Kalande, Evelyn Udedi, Chipawiru Mbalanga, Lauri Bruns, Thomas Coates. ^“^Beyond the Caregiver: Diffusion of early childhood development knowledge and practices within the social networks of HIV-positive mothers in Malawi”. Paper accepted for poster discussion. International AIDS Society. Mexico City, Mexico

2019	**Dovel, Kathryn**, Kelvin Balakasi, Khumbo Phiri*, Frackson Shaba*, O. Agatha Offorjebe*, Sundeep K Gupta, Vincent Wong, Eric Lungu, Brooke Nichols, Mike Nyirenda, Ngona K, Anteneh Worku, Risa Hoffman. ^“^A randomized trial on index HIV self-testing for sexual partners of ART clients in Malawi.” Paper accepted for oral presentation. Conference on Retroviruses and Opportunistic Infections (CROI). Seattle, Washington, USA

2019	Ogechukwu Offorjebe, **Kathryn Dovel**, Frackson Shaba, Kelvin Balakasi, Risa Hoffman, Sydney Rosen, Brooke Nichols, for the EQUIP Health team. Cost-effectiveness and national impact of index HIV self-testing in Malawi. Paper accepted for poster presentation. Conference on Retroviruses and Opportunistic Infections (CROI). Seattle, Washington, USA

2018	**Dovel, Kathryn**, Mike Nyirenda, Frackson Shaba*, Ogechukwu Offorjebe*, Kelvin Balakasi, Brooke Nichols, Khumbo Phiri*, Khumbo Ngona, Alan Schooley, Risa Hoffman on behalf of EQUIP Innovation for Health. “Facility-based HIV self-testing for outpatients dramatically increases HIV testing in Malawi: a cluster randomized trial.” Paper accepted for oral presentation. International AIDS Society. Amsterdam, Netherlands

2018	Shaba, Frackson*, Kelvin Balakasi, Ogechukwu Offorjebe*, Mike Nyirenda, Risa Hoffman, **Kathryn Dovel** on behalf of EQUIP Innovation for Health. “Facility-based HIV self-testing in Malawi: an assessment of characteristics and concerns among clients who opt-out of testing.” Paper accepted for poster presentation. International AIDS Society. Amsterdam, Netherlands

2018	**Dovel, Kathryn**, Mike Nyirenda, Frackson Shaba, Ogechukwu Offorjebe*, Kelvin Balakasi, Brooke Nichols, Khumbo Phiri*, Khumbo Ngona, Alan Schooley, Risa Hoffman on behalf of EQUIP Innovation for Health. “Facility-based HIV self-testing for outpatients dramatically increases HIV testing in Malawi: a cluster randomized trial.” Paper accepted for oral presentation. INTEREST. Kigali, Rwanda - awarded the Joep Lange INTEREST award

2018	Offorjebe , Ogechukwu*, Frackson Shaba, Kelvin Balakasi, Mike Nyrienda, Risa Hoffman, **Kathryn** **Dovel** on behalf of EQUIP Innovation for Health. “Partner-delivered HIV self-testing increases the perceived acceptability of index partner testing among HIV-positive clients in Malawi.” Paper accepted for mini-oral presentation. INTEREST. Kigali, Rwanda

2018	Stephanie Chamberlin*, Misheck Mphande, Pericles Kalande, **Kathryn** **Dovel** on behalf of EQUIP Innovation for Health. “Barriers and facilitators to consistent engagement in HIV care under Test and Treat in Malawi.” Paper accepted for poster presentation. INTEREST. Kigali, Rwanda

2017	**Dovel Kathryn,** Khumbo Phiri*, Alan Schooley, Misheck Mphande, Mackenzie Chivwara, Risa Hoffman. “Facility-level barriers to antiretroviral therapy experienced by men in Malawi.” Paper accepted for poster presentation. International AIDS Society. Paris, France

2017	Misheck Mphande, Khumbo Phiri*, Mackenzie Chivwara, Mike Nyirenda, Alan Schooley, Rachel Thomas, Risa Hoffman, **Kathryn Dovel**. “Examining Malawi’s Rollout of Universal Treatment: Policy Implementation and Provider Perceptions.” Paper accepted for poster presentation. International AIDS Society. Paris, France

2016	**Dovel, Kathryn**. “Factors influencing the implementation of provider-initiated testing and counseling (PITC) among STI clients in southern Malawi: A mixed methods study.” Paper accepted for poster presentation. International AIDS Society. Durbin, South Africa

2016	Westerhof, Nienke, Dzowela M, **Kathryn Dovel**, E. Banda, J. Chikonda. “Community-driven accountability through advocacy committees: a vehicle for improving HIV and reproductive health services for women living with HIV.” Paper accepted for poster presentation. International AIDS Society. Durbin, South Africa

2016	**Dovel, Kathryn**, Patrick Krueger, Shari Dworkin. “Predictors of men’s use of HIV testing services in low-income countries: the role of masculinity.” Paper accepted for poster presentation. Population Association of America. Washington D.C.

2014	**Dovel, Kathryn**. "Gender in HIV Policy: Examining how gender shapes the dissemination of HIV policies in southern Malawi." Paper accepted for roundtable presentation. American Public Health Association. New Orleans.

2014	**Dovel, Kathryn**. “Gendered care: examining how clinic experiences influence HIV testing decisions among STI patients in southern Malawi.” Paper accepted for oral presentation. National Women’s Studies Association, San Juan, Puerto Rico.

2013	**Dovel, Kathryn**. “HIV policies and their influence on men's use of care.” Paper accepted for oral presentation. International HIV Social Science and Humanities Conference, Paris, France.

2007	**Dovel, Kathryn**. “Social and structural impediments that limit proper healthcare in rural southern Kurdistan.” Paper accepted for oral presentation. The Anthropology and Sociology Research Conference, Santa Clara, CA.

**INVITED PRESENTATIONS**

2019	“Index HIV Self-Testing in Malawi”. World Health Organization webinar

2019	“Men’s (lack of) access to the health system”. UNAIDS. Regional meeting on Accelerating Men’s HIV service delivery and uptake in Eastern and Southern Africa.

2019	“Index HIVST in Malawi: a Randomized Control Trial. World Health Organization. Webinar

2019	“*Reaching men and engaging them in HIV care – lessons from Malawi*”. Men and HIV forum. International AIDS Society. Mexico City, Mexico

2018	“The impact of HIV self-testing on HIV testing among outpatients in high burden facilities in Malawi: preliminary findings from a cluster randomized control trial” USAID Washington. Washington D.C

2017	“Who benefits from Test and Treat? Understanding gender dimensions of universal treatment policies and gender-specific barriers to care” Malawi Ministry of Health, HIV Treatment Technical Working Group. Lilongwe, Malawi.

2016	“Facility-based barriers to HIV testing among men in Malawi: a systems approach” Malawi Ministry of Health, HIV Treatment Technical Working Group. Lilongwe, Malawi.

2015	“Men’s heightened risk of AIDS-related death: the legacy of gendered HIV testing and treatment strategies” United Nations Meeting on Male Engagement. Geneva, Switzerland

2015	“Facility-based barriers to men’s use of HIV testing: recommendations for male engagement guidelines.” National AIDS Control Council Meeting for the Development of the Male Engagement Guidelines. Nairobi, Kenya

2014	“Gendered care: examining who ‘does gender’ in clinical settings and its influence on HIV services for men in southern Malawi.” Health Working Group, UCLA. Los Angeles, CA

2013	“From questions to methods: mixed methods approach to disparities research.” Course in Qualitative Methods (Doctoral Students). University of Colorado Denver. Denver, C

2010	“Lost in translation: examples of why best-practice nutrition programs fail in rural Dominican Republic.” Drabkin and Bixby International Conference, UCLA. Los Angeles, CA

**RELEVANT EMPLOYMENT ACTIVITIES**

2015-	Consultant for Mixed Methods, Invest in Knowledge, Zomba, Malawi

Activities: Oversaw data analysis and write-up for studies implemented by Invest in Knowledge. I focused on qualitative and mixed methods analysis and write-up.

2009-10	Research Specialist, Korean Resource Center, Los Angeles, CA

Activities: Managed data entry and data cleaning and led in data analysis and write-up of a study assessing use of non-communicable disease services among first- and second-generation Korean populations in Los Angeles.

2009	Program Evaluation Fellow, Bataye Relief Alliance, Santo Domingo, Dominican Republic

Activities: Led the assessment of a nutritional program aimed to improve child health outcomes in Haitian populated bateyes in Dominican Republic. I led tool development, training enumerators, data analysis, and write-up

2007-08	Program Coordinator, Orange County Department of Public Health, Santa Ana, CA

Activities: Conducted literature reviews and assisting in the development of interventions to address Alcohol and Drug abuse among young adults in Orange County. Assisted in the protocol development and implementation of interventions.

**SERVICE**

2019	Committee Member of the Men’s HIV Forum at the International AIDS Conference, Mexico City

2018-	Member of the Malawi Ministry of Health HIV Self-Testing Guidelines Task Force

2017-	Member of the Malawi Ministry of Health HIV Testing Services Technical Working Group

2017-	Member of the EQUIP HIV Self-Testing Technical Working Group

2016-	Member of the UNAIDS Working Group “Engaging men in solutions for the HIV epidemic: Health systems.”

2016-	Member of the “Men and HIV Global Working Group”

2016	Reviewer for the APHA 2016 Annual Meeting & Expo

2012-13	Editor of the Health and Behavioral Sciences Peer-Reviewed Journal, University of

Colorado, Denver

2011-12	Student Advisory Council Member, University of Colorado, Denver

**COURSES TAUGHT**

**Adjunct Professor**

Social determinants of health in the context of HIV services in sub-Saharan Africa – Field Rotation Series (UCLA)

Health, Disease & Globalization: Foundations of Epidemiology (Vanguard University)

Human Sexuality (co-taught, Vanguard University)

Cultural Anthropology (Vanguard University)

Applied Anthropology (Vanguard University)

Qualitative Methods (Vanguard University)

**Teaching Assistant**

AIDS and Other Sexually Transmitted Diseases (UCLA)

Global Health Issues (UCLA)

Social Determinants of Health (University of Colorado Denver)

Statistical Analysis (University of Colorado Denver)

**MENTORSHIP**

University of California Los Angeles. David Geffen School of Medicine. Medical Student. Kate Coursey. “Examining characteristics of women who engage in an integrated Early Childhood Development and PMTCT program in Malawi: endline evaluation.” 2019-

University of California Los Angeles. David Geffen School of Medicine. Medical Student. “Provider acceptability of interventions to increase ART initiation among men who test HIV-positive through index HIV self-testing.” 2019-

University of California Los Angeles. David Geffen School of Medicine. Medical Student. Tijana Temelkovska. “Examining the successes and challenges of implementing an early childhood development intervention with HIV-positive women in Malawi: a process evaluation.” 2018-

University of California Los Angeles. Internal Medicine Residency, Global Health Track. Resident Physician. Marguerite Thorp. “Can a brief screening tool identify ART clients at risk of defaulting from treatment? a prospective study in Malawi.” 2018-

University of California Los Angeles. Internal Medicine Residency, Global Health Track. Resident Physician. Adrian Mayo. “Predictors of early ART retention among adults who initiated under Universal Treatment policies in Malawi.” 2018-

University of Colorado Denver. Health and Behavioral Sciences. Doctoral Student. Stephanie Chamberlin. “Exploring the association between education and ART retention in rural Malawi.” 2017-

University of California Los Angeles. Fogarty GloCal Fellow. Medical Student. Ogechukwu Offorjebe. “Examining the feasibility and acceptability of HIV self-test kits for index testing among HIV+ clients and their partners in Malawi: A mixed methods study.” 2017-18

College of Medicine, Malawi. MPH Student. Khumbo Phiri. “The role of lay cadre in ART initiation and retention under Test and Treat in Malawi.” 2017-18

Brandeis University. Elisa Morales, Becca Sliwosk, and Melanie Morris (capstone project). “Developing a funding proposal for Men-to-Men, a gender-transformation and income-generating program for men in northern Uganda.” 2015 (with 31Bits International)

Vanguard University. Medical Anthropology Honors Thesis. Joanna Takegami. “Barriers to Women’s use of Antiretroviral Therapy in Northern Uganda: Exploring the Role of Structural Violence.” 2011

**AD HOC REVIEWER**

AIDS, JAIDS, JIAS, BMC Public Health, Global Health Action, Culture, Health and Sexuality

**PROFESSIONAL MEMBERSHIPS**

Member, American Public Health Association (APHA), Present

Member, American Sociology Association (ASA), Present

Member, American Anthropological Association (AAA), Present

OMB No. 0925-0001 and 0925-0002 (Rev. 10/15 Approved Through 10/31/2018)

**THOMAS J. COATES**

eRA COMMONS USER NAME (credential, e.g., agency login): **TCOATS**

POSITION TITLE: **Professor Emeritus, Division of Infectious Diseases, Department of Medicine UCLA David Geffen School of Medicine**

EDUCATION/TRAINING (Begin with baccalaureate or other initial professional education, such as nursing, include postdoctoral training and residency training if applicable. Add/delete rows as necessary.)

| INSTITUTION AND LOCATION | DEGREE  (if applicable) | Completion Date  MM/YYYY | FIELD OF STUDY |
| --- | --- | --- | --- |
| San Luis Rey College, San Luis Rey, California | BA | 06/1968 | Philosophy |
| San Jose State University, San Jose, California | MA | 01/1971 | Psychology |
| Stanford University, Stanford, California | PhD | 06/1977 | Counseling Psychology |

A.	Personal Statement

I am Director of the system-wide University of California Global Health Institute (founded in 2008) and was the Founding Director of the UCLA Center for World Health (founded in 2012) until 2018. In 1986 I co-founded the Center for AIDS Prevention Studies (CAPS) at UCSF and directed it from 1991 to 2003. I was also the founding Director of the UCSF AIDS Research Institute, leading it from 1996 to 2003.

I have substantial expertise in research on HIV prevention among heterosexual men and women in the HIV epidemic in sub-Saharan Africa and in the HIV testing and treatment trials in sub-Saharan Africa, especially Malawi through PEPFAR funding. As Distinguished Research Professor of Medicine, I continue with two NIH and two foundation grants focused in southern Africa. I also continue as a co-investigator on the UCLA-based Center for HIV Identification, Prevention and Treatment Studies (CHIPTS).

I have had extensive experience with large-scale, community-based, multi-site research and implementation projects spanning HIV prevention, care and treatment, and policy. I currently have funding to test and evaluate innovative strategies for bring men in South Africa into HIV testing and treatment, as well as for providing early childhood development training for HIV-infected mothers and their babies in Malawi through support from the Conrad N. Hilton Foundation. We are also in the first year of a 5-year NIH-funded grant to study pre-exposure prophylaxis for pregnant and post-partum women in South Africa.

B.	Positions and Honors

1984 - 2003	Member, Medical Attending Staff, UCSF Hospitals and Clinics

1990 - 2003	Professor, Department of Medicine, UCSF

1991 - 2003	Director, Center for AIDS Prevention Studies, UCSF

1996 - 2003	Director, AIDS Research Institute, UCSF

2000	Elected to the Institute of Medicine (now the National Academy of Medicine)

2010 - 2014	Member, Institute of Medicine Board on Global Health

2003 - 2006	Professor Step VII, Division of Infectious Diseases, Department of Medicine, David Geffen School of Medicine, UCLA

2003 - Present	Joint Appointment, Department of Medicine, UCSF; Member, Executive Committee, UCLA AIDS Institute

2003 - 2011	Director, UCLA Program in Global Health

2004 - Present	Joint Appointment, Department of Epidemiology, UCLA School of Public Health

2006 – 2009	Professor Step IX, Division of Infectious Diseases, Department of Medicine, David Geffen School of Medicine, UCLA

2006 - 2018	Michael & Sue Steinberg Endowed Professor of Global AIDS Research, Division of Infectious Diseases, Department of Medicine, David Geffen School of Medicine, UCLA

2006 - 2018	Director, Global Capacity Building Core Center for HIV Identification, Prevention, and Treatment Services, UCLA Semel Neurosciences Institute

2006 - 2018	Associate Director for International and Policy Research UCLA AIDS Institute

2009 - 2016	Co-director, University of California Global Health Institute

2009 –2018	Distinguished Professor, Division of Infectious Diseases, Department of Medicine, David Geffen School of Medicine,	University of California, Los Angeles

2011 – 2018	Director, UCLA Center for World Health at the David Geffen School of Medicine and UCLA Health

2016-Present	Director, University of California Global Health Institute

2018-Present	Distinguished Research Professor, Division of Infectious Diseases, UCLA David Geffen School of Medicine

C.	Contribution to Science

1. **Combination HIV Prevention including Pre-Exposure Prophylaxis:** I have written extensively and conducted research on combination HIV prevention for MSM in the United States and Latin America and with a variety of populations in sub-Saharan Africa. My writing and research have been influential in shaping thinking about combination prevention, and in demonstrating the importance of considering HIV prevention as a combination of factors, as opposed to any single kind of program.

Joseph Davey D, Bekker LG, Gorbach P, **Coates T**, Myer L. Delivering PrEP to pregnant and breastfeeding women in sub-Saharan africa: The implementation science frontier. AIDS. 2017 Jul 18. doi: 10.1097/QAD.0000000000001604. PubMed PMID: 28723709.

Richter L, Komárek A, Desmond C, Celentano D, Morin S, Sweat M, Chariyalertsak S, Chingono A, Gray G, Mbwambo J, **Coates T**; Reported physical and sexual abuse in childhood and adult HIV risk behaviour in three African countries: findings from Project Accept (HPTN-043). AIDS and behavior. 2014; 18(2):381-9. PMCID: PMC3796176

**Coates TJ**; An expanded behavioral paradigm for prevention and treatment of HIV-1 infection. Journal of acquired immune deficiency syndromes (1999). 2013; 63 Suppl 2:S179-82. PMCID: PMC3943341

**Coates TJ**, Richter L, Caceres C. Behavioural strategies to reduce HIV transmission: how to make them work better. Lancet. 2008; 372(9639):669-84. PMCID: PMC2702246

2. **HIV Counseling and Testing (HTC):** I have conducted many significant and influential studies in HTC, beginning first with observational studies of the effect of HTC on risk behavior among men who have sex with men (MSM) in San Francisco. I was Principal Investigator for the first randomized controlled trial of HTC in Eastern Africa and the Caribbean, examining the effect of HTC on individual males and females, as well as couples presenting for HTC in Kenya, Tanzania, and Trinidad and Tobago, and these results were reported in *The Lancet* in 2000. I was the Principal Investigator for Project Accept, a cluster randomized trial conducted in South Africa, Zimbabwe, Tanzania, and Thailand, and these results were reported in *Lancet Global Health* in 2015. I also was the Principal Investigator of a randomized trial at Mulago Hospital in Uganda examining the effect of short vs. elaborated counseling on males and females presenting for care, and these results were reported in *Lancet Global Health* in 2014.

**Coates TJ**, Kulich M, Celentano DD, Zelaya CE, Chariyalertsak S, Chingono A, Gray G, Mbwambo JK, Morin SF, Richter L, Sweat M, van Rooyen H, McGrath N, Fiamma A, Laeyendecker O, Piwowar-Manning E, Szekeres G, Donnell D, Eshleman SH; NIMH Project Accept (HPTN 043) study team; Effect of community-based voluntary counselling and testing on HIV incidence and social and behavioural outcomes (NIMH Project Accept; HPTN 043): a cluster-randomised trial. The Lancet. Global Health. 2014; 2(5):e267-77. PMCID: PMC4131207

van Rooyen H1, McGrath N, Chirowodza A, Joseph P, Fiamma A, Gray G, Richter L, **Coates T**. Mobile VCT: reaching men and young people in urban and rural South African pilot studies (NIMH Project Accept, HPTN 043). AIDS and behavior. 2013; 17(9):2946-53. PMCID: PMC3597746

Wanyenze RK, Kamya MR, Fatch R, Mayanja-Kizza H, Baveewo S, Szekeres G, Bangsberg DR, **Coates T**, Hahn JA; Abbreviated HIV counselling and testing and enhanced referral to care in Uganda: a factorial randomised controlled trial. The Lancet. Global Health. 2013; 1(3):e137-45. PMCID: PMC4129546

Mhlongo S, Dietrich J, Otwombe KN, Robertson G, **Coates TJ**, Gray G.Factors associated with not testing for HIV and consistent condom use among men in Soweto, South Africa. PloS one. 2013; 8(5):e62637. PMCID: PMC3656000

3. **Global Health:** I have contributed to the literature on global health, especially from the perspective of engaging multiple disciplinary perspectives to attend to a variety of global health issues around the world.

Debas HT, **Coates TJ**; The University of California Global Health Institute opportunities and challenges.

Infectious disease clinics of North America. 2011; 25(3):499-509, vii. PubMed [journal]PMID: 21896355

Duber HC, **Coates TJ**, Szekeras G, Kaji AH, Lewis RJ; Is there an association between PEPFAR funding and improvement in national health indicators in Africa? A retrospective study. Journal of the International AIDS Society. 2010; 13:21. PMCID: PMC2895577

Maman S, Abler L, Parker L, Lane T, Chirowodza A, Ntogwisangu J, Srirak N, Modiba P, Murima O, Fritz K.A comparison of HIV stigma and discrimination in five international sites: the influence of care and treatment resources in high prevalence settings. Social science & medicine (1982). 2009; 68(12):2271-8. PMCID: PMC2696587

Collins C, Coates TJ, Szekeres G; Accountability in the global response to HIV: measuring progress, driving change. AIDS (London, England). 2008; 22 Suppl 2:S105-111. PMCID: PMC2879260

**Complete List of Published Work in MyBibliography:** <http://www.ncbi.nlm.nih.gov/sites/myncbi/thomas.coates.1/bibliography/40839346/public/?sort=date&direction=descending>

D.	Research Support

**Ongoing Research Support**

R01MH105534-01A1 (Coates)	07/07/15 – 04/30/20

NIH/NIMH

**Bringing South African Men into HIV Counseling and Testing (HCT) and Care**

The objective of this project is to provide evidence-based strategies to improve treatment of HIV+ men through a three-step process: (1) Testing a significant proportion of the population, (2) linkage to care, and (3) maintaining in care a significant proportion of HIV+ individuals to the point of viral suppression. My role is as the Principal Investigator.

UM1 AI068619 (El Sadr)	07/01/14 – 11/30/20

Family Health International

NIH-NIAID

**HIV Prevention Trials Network (HPTN) Leadership Group**

The goals of this project are: 1) to develop the HPTN research agenda; 2) to review SWG research plans; 3) to review and approve concept plans; 4) to oversee the discretionary fund; 5) to review and revise HPTN policies and procedures; and 6) to evaluate the performance of the HPTN. My role is as Chair of the Manuscript Review Committee

P30 MH058107 (Shoptaw)	03/01/2017-02/28/2022

NIMH/NIH

**Center for HIV Identification, Prevention, and Treatment Services**

This project is a P30 and provides center grant services to HIV investigators at UCLA. I am a Co-Investigator in this center.

The Conrad N. Hilton Foundation	01/01/2018-12/31/2020

**Delivery of Childhood Development Services as Part of HIV Treatment Services in Malawi**

This grant supports the integration of early childhood development services within pre- and post-natal care for HIV+ mothers in Malawi.

R01 MH116771-01A1	09/30/2018-09-29-2023

NIMH/NIH

**Evaluating the Prep-PP Cascade in HIV-negative Pregnant and Breastfeeding Women in South Africa.**

The goal of this project is to test innovative models for delivering PrEP to pregnant and breastfeeding women age 16 and above in South Africa.

**Entertainment Industry Foundation-Charlize Theron Africa Outreach Project** 06/01/2018-05/30/2021

The goal of this project is to create a Youth Leaders Scholarship Fund to support promising young South Africans to attend South African tertiary education institutions.

Bill and Melinda Gates Foundation **(**Dovel)	12/4/2019-12/3/2023

**Identifying Effective Linkage Strategies for HIVST (IDEaL)**

This grant tests the effect of a staged intervention for ART initiation among men in Malawi, whereby additional interventions are added each month for individuals who have not yet initiated ART.

**Completed Research Support**

20150025 (Coates)	09/01/15 – 08/31/17

Conrad N. Hilton Foundation

**Delivery of Early Childhood Development Services as a Part of HIV Treatment Services in Malawi**

Pilot grant to assess the feasibility and acceptability, as well as initial outcomes, of supporting Option B+ mothers in Malawi to increase their responsiveness to their children and have positive impacts on early childhood development (ECD).

P30 MH58107 (Rotheram-Borus)	02/01/07 - 01/31/17

NIH/NIMH

**Center for HIV Identification, Prevention, and Treatment Services (CHIPTS)**

The mission of the Center for HIV Identification, Prevention, and Treatment Services (CHIPTS) is to promote collaborative research and education on effective HIV detection, prevention, and treatment programs for HIV at the societal, community, provider, and individual levels. My role is as the Director for International Care.

**RISA MICHELLE HOFFMAN**

CURRICULUM VITAE

**PERSONAL HISTORY**

David Geffen School of Medicine at UCLA

Division of Infectious Diseases

10833 Le Conte Ave 37-121 CHS

Los Angeles, CA 90095

Tele: (310) 825-7225

**EDUCATION**

Stanford University 1994, BA

University of California Los Angeles 2000, MD

Harvard School of Public Health 2000, MPH

Internship 2000-2001: Harvard Combined Medicine/Pediatrics Residency Program

Residency 2001-2004: Harvard Combined Medicine/Pediatrics Residency Program

Fellowship Infectious Diseases: 2005-2008: University of California, Los Angeles

**LICENSURE**

California, A85173, 01/31/2021

**BOARD CERTIFICATION/OTHER CERTIFICATION**

2004 & 2014 American Board of Internal Medicine

2007 & 2017 American Board of Internal Medicine, Infectious Diseases

2005 Certification in Travel Medicine from the London School of Hygiene and Tropical Medicine

**PROFESSIONAL EXPERIENCE**

*Present Position*

2016-present	Associate Clinical Professor, Division of Infectious Diseases, UCLA

2010-2016	Assistant Clinical Professor, Division of Infectious Diseases, UCLA Medical Center, Los Angeles, California

2008-2010	Clinical Instructor, Division of Infectious Diseases, UCLA Medical Center, Los Angeles, California

*Previous Positions*

2005-2008	Fellow in Infectious Diseases, UCLA Medical Center, Los Angeles, California

2001-2004	Resident Physician, Internal Medicine, Brigham and Women’s Hospital, Boston, Massachusetts

2001-2004	Resident Physician, Pediatrics, Boston Children’s Hospital and Massachusetts General Hospital, Boston, Massachusetts

2000-2001	Intern, Internal Medicine, Brigham and Women’s Hospital, Boston, Massachusetts

2000-2001	Intern, Pediatrics, Boston Children’s Hospital and Massachusetts General Hospital, Boston, Massachusetts

**PROFESSIONAL ACTIVITIES & MEMBERSHIPS**

2018-present	Interim Director, Global Health Education and Research Program, David Geffen School of Medicine at UCLA

2016-present	Co-Director UCLA AIDS Institute/CFAR International Health Services and Policy Research Program Section

2015-present	Associate Program Director, UCLA Infectious Diseases Fellowship Training Program

2013-present	Advisory Board Member for the University of California Global Health Institute GloCal Health Fellowship

2009-present	Research Co-Director, Partners in Hope Malawi and UCLA Research Collaboration

2009-present	Investigator, AIDS Clinical Trials Group (ACTG) and Maternal Child Adolescent Network (IMPAACT)

2009-present	HIV Clinical Consultant, To Help Everyone Clinic in Los Angeles, California

2009-present	Ad hoc Peer Reviewer (AIDS Care, International Journal of STD and AIDS, BMC Women’s Health, American Society of Tropical Medicine and Hygiene, Journal of Infectious Diseases, International Health, JIAS)

2007-present	Member, Infectious Diseases Society of America (IDSA)

2016-2018	Committee Member, Antiretroviral Therapy Strategies (ARTs), AIDS Clinical Trials Group

2008-2016	Founder/Program Director, Sustainable Nutrition for Orphans and Vulnerable Children in Malawi, Central Africa: Provides education on nutrition and sustainable food sources for families caring for orphans in northern Malawi

2014-2016	Committee Lead, Infectious Diseases Quality Improvement M&M Program

2011-2016	Committee Member, AIDS Clinical Trials Group *Women’s Health Inter-network Scientific Committee (WHISC)*

2007-2013	Founder/Program Co-Director, UCLA resident physician elective training program in Malawi, Africa

2010-2013	Co-Director, UCLA Program in Global Health and Global Health Education Program for the David Geffen School of Medicine at UCLA

2008-2012	Faculty for ‘Multidisciplinary Approach to Global Health’ elective course for first and second year medical students at UCLA

2007-2012	Committee Member, American Society of Tropical Medicine and Hygiene Education Committee

2005-2012	Advisory Board Member, UCLA Medicine/Pediatrics Residency Training Advisory Board

2005-2011	Interviewer, UCLA Medicine/Pediatrics Residency Training Program

2006-2008	Faculty Group Leader, Problem Based Learning Microbiology Block for second year medical students at UCLA

2006-2008	Creator/Organizer, UCLA Infectious Diseases Core Curriculum Program

**HONORS AND AWARDS**

2012	David Geffen School of Medicine Award for Excellence in Education

2011	Nomination for the Consortium of Universities for Global Health Early Career Award

2009	Nomination for UCLA Faculty Teaching Award

2007	Nomination for UCLA Fellow Teaching Award

2006	Nomination for UCLA Fellow Teaching Award

2000	Elected to the UCLA chapter of the Alpha Omega Alpha Honor Society

2000	Janet M. Glasgow Memorial Achievement Citation for Academic Achievement at the UCLA School of Medicine

2000	John M. Adams Award for Excellence in Pediatrics, UCLA School of Medicine

2000	Edith and Carl Lasky Memorial Award for Outstanding Research Achievement, UCLA School of Medicine

1999	Longmire Surgical Medal for outstanding performance in surgical clerkships, awarded by the Department of Surgery, UCLA School of Medicine

1999	Charles Wacker Summer Research Fellowship, UCLA School of Medicine

1994	Elected to the Stanford Chapter of Phi Beta Kappa

1994	Elected to the Stanford Cap and Gown Women’s Honor Society

1994	Recipient of the Joshua Lederberg Award for Outstanding Academic Achievement in Human Biology

**LECTURES AND PRESENTATIONS**

“Management of Febrile Neutropenia” Presented as part of the UCLA Division of Infectious Diseases Core Curriculum Program. Los Angeles, California, November 2013

“Challenges and Successes of EQUIP Malawi” Presented at UCLA Infectious Diseases Grand Rounds. Los Angeles, California, April 2014

“Primary Care Issues in HIV Care” Presented at the UCLA Department of Medicine housestaff curriculum. Los Angeles, California, May 2014

“A Case of Multi-Class HIV Resistance” Presented at the UCLA HIV/Hepatitis C Case Conference Series, Los Angeles, California, May 2014

“Evidence Based Managed of Osteomyelitis” Presented at the UCLA Division of Infectious Diseases Case Conference Series, Los Angeles, California, June 2014

“Responding to Viral Load: A Primer for Malawi Clinical Mentors” Presented at a PEPFAR EQUIP Clinical Training Meeting in Malawi, Africa, January 2015

“Clinical Pearls in the Management of HIV/AIDS”. Presented at the UCLA Internal Medicine Resident Core Curriculum Conference Series, Los Angeles, California, February 2015

“Quality Improvement in ID Care at UCLA: Lessons Learned from M&M” Presented at the UCLA Division of Infectious Diseases Core Conference Series, Los Angeles, California, April 2015

“Health & Safety Overseas: An orientation for medical students” Presented at the UCLA Global Health Education Medical Student Orientation Program, Los Angeles, California, April 2015

“ID Mimics”. Presented at the UCLA ID Fellow Core Curriculum Series, Los Angeles, California, May 2015

“Quality Improvement on the Infectious Diseases Service: Transition of Care.” Presented at the UCLA Division of Infectious Diseases Case Conference, Los Angeles, California, June 2015

“Quality Improvement on the Infectious Diseases Service: Notes and Documentation.” Presented at the UCLA Division of Infectious Diseases Case Conference, Los Angeles, California, December 2015

“Quality Improvement on the Infectious Diseases Service: HIV Care”. Presented at the UCLA Division of Infectious Diseases Case Conference, Los Angeles, California, February 2016

“Multi-month scripting to achieve improved outcomes in EQUIP”. Presented at the EQUIP annual meeting, Johannesburg, South Africa March 2016

“Clinical Management of HIV/AIDS for the Primary Care Resident”. Presented at the UCLA Internal Medicine Resident Core Curriculum Conference Series, Los Angeles, California, April 2016

“Update on Option B+ in Malawi”. Presented to the Women’s Health Committee of the AIDS Clinical Trials Group, Los Angeles, California, April 2016

EQUIP Malawi: A Partnership for HIV Care in Malawi. Presented at Harbor UCLA Infectious Diseases Grand Rounds, Los Angeles, California, July 2016

Speaker, Infectious Diseases Career Panel for Medical Students at the David Geffen School of Medicine. Los Angeles, September 2016

Systemwide Case Conference Faculty Discussant for the MultiCampus Infectious Diseases Fellowship Program. Presented at the VA Hospital, Los Angeles, California, December 2016

“Introduction to Global HIV Treatment in Resource Poor Settings,” Lecturer for the UCLA School of Public Health, February 2018, Los Angeles

UCLA Division of Infectious Diseases, Journal Club Faculty Discussant, MDR TB Treatment, March 2018, Los Angeles

Faculty Panelist. Global Health Career Night for the David Geffen School of Medicine. November 2018, Los Angeles

West LA VA Internal Medicine Grand Rounds Speaker: “The Intersection of HIV and Non-Communicable Diseases in Resource-Limited Settings” April 2019, Los Angeles

“Qualitative Client and Provider Experiences with Multi-Dispensing for HIV in Malawi and Zambia”. Presented as part of the CQUIN Consortium. Webinar, April 2019

“Introduction to the Global Health Program”. Presented as part of the DGSOM Global Health Selective, September 2019, Los Angeles

**PUBLICATION/BIBLIOGRAPHY**

**RESEARCH PAPERS**

**RESEARCH PAPERS (PEER REVIEWED)**

**Hoffman RM**, Umeh OC, Garris C, Givens N, Currier JS. Evaluation of Sex Differences of Fosamprenavir (With and Without Ritonavir) in HIV-infected Men and Women. HIV Clin Trials. 2007;8(6):371-380.

**Hoffman RM**, AboulHosn J, Child JS, Pegues DA. Bartonella Endocarditis in Complex Congenital Heart Disease. Congenit Heart Dis. 2007;2(1):79-84.

Black V, **Hoffman RM**, Sugar CA, Menon P, Venter FWD, Currier JS, Rees H. Safety and Efficacy of Initiating Highly Active Antiretroviral Therapy in an Integrated Antenatal and HIV Clinic in Johannesburg, South Africa. J Acquir Immune Defic Syndr. 2008;49(3):276-81. PMC2893046.

**Hoffman RM**, Black V, Technau K, van der Merwe KJ, Currier JS, Coovadia A, Chersich M. Effects of Highly Active Antiretroviral Therapy Duration and Regimen on Risk for Mother-to-Child Transmission of HIV in Johannesburg, South Africa. J Acquir Immune Defic Syndr. 2010;54(1):35-41. PMC2880466.

Pilotto JH, Velasque L, Khalili R, Ismerio R, Veloso VG, Grinsztejn B, Morgado MG, Watts DH, Currier JS, **Hoffman RM**. Maternal Outcomes after HAART for Prevention of Mother-to-Child Transmission in HIV-infected Women in Brazil. Antivir Ther. 2011;16(3):349-56. PMC3437753.

Hoffman RM, Jamieson BD, Bosch RJ, Currier JS, Kitchen CMR, Schmid I, Zhu Y, Bennett K, Mitsuyasu R. Baseline Immune Phenotypes and CD4+ T Lymphocyte Responses to Antiretroviral Therapy in Younger versus Older HIV-infected Individuals. J Clin Immunol. 2011;31(5):873-81. PMC3194061.

Van der Merwe J, Hoffman RM, Black V, Chersich M, Coovadia A, Rees H. Birth outcomes in South African Women Receiving Highly Active Antiretroviral Therapy: a Retrospective Observational Study. J Int AIDS Soc. 2011;14:42. PMC3163172.

Mindry D, Wagner G, Lake JE, Smith A, Linnemayr S, Quinn M, **Hoffman RM**. Fertility Desires Among HIV-infected Men and Women in Los Angeles County: Client Needs and Provider Perspectives; Matern Child Health J. 2013 May;17(4):593-600. PMC N/A.

Burke Z, Chen J, Conceicao C, **Hoffman R**, Miller L, Taela A, DeUgarte DA. Evaluation of Preoperative and Intraoperative RBC Transfusion Practices in Maputo Central Hospital, Mozambique. Transfusion. 2013 May 21. doi: 10.1111/trf.12252. PMC3751985.

**Hoffman RM**, Leister E, Kacanek D, Shapiro DE, Read JS, Bryson Y, Currier JS. Biomarkers from late pregnancy to six weeks postpartum in HIV-infected women who continue versus discontinue antiretroviral therapy after delivery. JAIDS. 2013 May 8. PMC3868443.

Jaganath D, Mulenga C, **Hoffman R**, Hamilton J, Boneh G. This is My Story: Participatory Performance for HIV and AIDS Education at the University of Malawi. Health Education Research. 2013 Sep 18. PMC4155417.

Kawale P, Mindry D, Stramotas S, Chilikoh P, Phoya A, Henry K, Elashoff D, Jansen P, **Hoffman R**. Factors associated with desire for children among HIV-infected women and men: A quantitative and qualitative analysis from Malawi and implications for the delivery of safer conception counseling. AIDS Care. 2013 Jun;26(6). PMC3943633.

1. Iroezi N, Mindry D, Kawale P, Chikowi G, Jansen P, **Hoffman R**. A qualitative analysis of the barriers and facilitators to receiving care in a prevention of mother-to-child program in Nkhoma, Malawi. Afr JReprod Health. 2013 Dec;17(4). PMC4361063.
2. Russell E, Mohammed T, Smeaton L, Jorowe B, MacLeod I, **Hoffman R**, Currier JS, Moyo S, Essex M, Lockman S. Immune activation markers in peripartum women in Botswana: association with feeding strategy and maternal morbidity. PLoS One. 2014 Mar 21. PMC3962339.
3. Reddy D, Njala J, Stocker P, Schooley A, Flores M, Tseng C-H, Pfaff C, Jansen P, Mitsuyasu RT, **Hoffman RM**. High-risk human papillomavirus in HIV-infected women undergoing cervical cancer screening in Lilongwe, Malawi: A pilot study. International Journal of STDS and AIDS, 2014 Jun 13. PMC4363075.
4. Kamuyango A, Hirschhorn L, Wang W, Jansen P, **Hoffman R**. One-Year Outcomes of Women Started on Antiretroviral Therapy during Pregnancy before and after the Implementation of Option B+ in Malawi: A Retrospective Chart Review from Three Facilities. World Journal of AIDS. 2014 Sept;4(3). PMC4356991.
5. Shull H, Tymchuk C, Grogan T, Hamilton J, Friedman J, **Hoffman RM**. Evaluation of the UCLA Department of Medicine Malawi Global Health Clinical Elective: Lessons from the First Five Years. Am J Trop Med Hyg. 2014 Sep 15. PMC4228879.
6. Hoffman JC, Anton PA, Baldwin GC, Elliott J, Anisman-Posner D, Tanner K, Grogan T, Elashoff D, Sugar C, Yang OO, **Hoffman RM**. Seminal Plasma HIV-1 RNA Concentration is Strongly Associated with Altered Levels of Seminal Plasma Interferon Gamma, Interleukin-17, and Interleukin-5. AIDS Res Hum Retroviruses. 2014 Oct 2. PMC4208556.
7. Menon P, **Hoffman R**, Black V. Characteristics of HIV-infected women on antiretroviral therapy who develop preeclampsia in South Africa: A Case Series. The Journal of Global Health. 2014;4(2). PMC N/A.
8. Cheng Q, Engelage E, Grogan T, Currier JS, and **Hoffman RM**. Who provides primary care? A cross-sectional survey of HIV patients and providers in a Los Angeles clinic. AIDS Clinical Research. 27 Oct 2014;5(11). PMC4409003.
9. Kawale P, Mindry D, Phoya A, Jansen P**, Hoffman RM**. Provider attitudes about childbearing and knowledge of safer conception at two HIV clinics in Malawi. Reproductive Health. 2015 Mar 7;12(1):17. PMC4355153.
10. Yeatman S, **Hoffman RM,** Chilungo A, Lungu S, Namadingo H, Chimwaza A, and Trinitapoli JA. Health-seeking behavior and symptoms associated with early HIV infection: Results from a population-based cohort in southern Malawi. JAIDS. 2015 May 1;69(1):126-30. PMC4422188.
11. **Hoffman RM**, Jaycocks A, Vardavas V, Wagner G, Lake JE, Mindry D, Currier JS, Landovitz R. Benefits of PrEP as an adjunctive method of HIV prevention during attempted conception between HIV-uninfected women and HIV-infected male partners. Journal of Infectious Diseases, J Infect Dis. 2015 Jun 19. PMC4621256.
12. Lake JE, **Hoffman RM**, Tseng CH, Wilhalme HM, Currier JS. Success of Standard Dose Vitamin D Supplementation in Treated HIV Infection. Open Forum Infect Dis. 2015 May 15;2(2). PMC4462892.
13. Chipungu C, Veltman JA, Jansen P, Chiliko P, Lossa C, Namarika D, Benner B, **Hoffman RM**, Bristow CC, Klausner JD. Feasibility and Acceptability of Cryptococcal Antigen Screening and Prevalence of Cryptocococcemia in Patients Attending a Resource-limited HIV/AIDS clinic in Malawi. Journal of the International Association of Providers of AIDS Care, J Int Assoc Provid AIDS Care. 2015 Jul 2. PMC N/A.
14. Coelho L, Cardoso SW, Luz PM, **Hoffman RM**, Mendonca L, Veloso VG, Currier JS, Grinsztejn B, Lake JE. Vitamin D3 supplementation in HIV infection: effectiveness and associations with antiretroviral therapy. Nutr J. 2015 Aug 18: 14. PMC4538921.
15. **Hoffman RM**, Lake JE, Wilhame H, Tseng CH, Currier JD. Vitamin D levels and markers of inflammation and metabolism in HIV-infected individuals on suppressive antiretroviral therapy. AIDS Res and Human Retroviruses 2016 March;32(3). PMC4779972.
16. Herbst de Cortina S, Arora G, Wells T, **Hoffman RM**. Evaluation of a Structured Predeparture Orientation at the David Geffen School of Medicine’s Global Health Education Programs. Am J Trop Med Hyg. 2016 Mar 2;94(3). PMC4775891.
17. Jordan J, **Hoffman R**, Arora G, Coates W. Activated learning: providing structure in global health education at the David Geffen School of Medicine at the University of California, Los Angeles – a pilot study. BMC Medical Education. 2016 Feb 16. PMC4755030.
18. Wagner GJ, Linnemayr S, Ghosh-Dastidar B, Currier JD, **Hoffman R**, Schneider S. Supporting Treatment Adherence Readiness through Training (START) for patients with HIV on antiretroviral therapy: a study protocol for a randomized controlled trial. Trials. 2016 March 24;17(1). PMC4806419.
19. Arora G, Perkins K, **Hoffman RM**. Optimizing global health electives through partnerships: A pilot study of pediatric residents. Academic Pediatrics. 2015 Sept-Oct;15(5). PMC N/A.
20. Chien E, Phiri K, Schooley S, Chivwala M, Hamilton J, **Hoffman RM**. Successes and Challenges of HIV Mentoring in Malawi: The Mentee Perspective. PLoS One. 2016 Jun 18;11(6). PMC4924818.
21. Schooley A, Kamudumuli P, Vangala S, Tseng CH, Soko C, Parent J, Phiri K, Jahn A, Namarika D, **Hoffman RM**. CD4 Variability in Malawi: Implications for Use of a CD4 Threshold of 500 Cells/mm3 Versus Universal Eligibility for Antiretroviral Therapy. Open Forum Infectious Diseases. 2016 Aug 29;3(3). PMC5047419.
22. Hemarajata P, Baghdadi JD, **Hoffman R**, Humphries RM. Burkholderia pseudomallei: Challenges for the Clinical Microbiology Laboratory. J Clin Microbiol 2016 Dec;54(12):2866-2873. PMC5121373.
23. Pfaff C, Scott V, **Hoffman R**, Mwagomba B. You can treat my HIV – But can you treat my blood pressure? Availability of integrated HIV and non-communicable disease care in northern Malawi. Afr J Prim Health Care Fam Med. 2017 Feb 15;9(1):e1-e8. PMC5320467.
24. Gibb J, Chitsulo J, Chipungu C, Chivwara M, Schooley A, **Hoffman RM**. Supporting Quality Data Systems: Lessons Learned from Early Implementation of Routine Viral Load Monitoring at a Large Clinic in Lilongwe, Malawi. Clinical Research in HIV AIDS and Prevention. 2017 Mar 14;2(4). PMC5502771
25. Arora G and **Hoffman RM**. Development of an HIV Postexposure Prophylaxis (PEP) Protocol for Trainees Engaging in Academic Global Health Experiences. Acad Med. 2017 Apr 25. PMC28445222.
26. **Hoffman RM**, Phiri K, Parent J, Grotts J, Elashoff D, Kawale P, Yeatman S, Currier JS, Schooley A. Factors associated with retention in Option B+ in Malawi: a case control study. J Int AIDS Soc. 2017 Apr 27;20(1):1-8. PMC28453243.
27. Currier JS, Britto P, **Hoffman RM**, Brummel S, Masheto G, Joao E, Santos B, Aurpibul L, Losso M, Pierre MF, Weinberg A, Gnanashanmugam D, Chakhtoura N, Klingman K, Browning R, Coletti A, Mofenson L, Shapiro D, Pilotto J. Randomized Trial of Stopping or Continuing ART among Postpartum Women with Pre-ART CD4 ≥ 400 cells/mm3 (PROMISE 1077HS). PLoS One. 2017 May 10;12(5):e0176009. PMC28489856.
28. **Hoffman R**, Bardon A, Rosen S, Fox M, Kalua T, Xulu T, Taylor A, Sanne I. Varying intervals of antiretroviral medication dispensing to improve outcomes for HIV patients (The INTERVAL Study): study protocol for a randomized controlled trial. Trials. 2017 Oct 13;18(1):476. PMC29029644.
29. Judson SD, LeBreton M, Fuller T, **Hoffman RM**, Njabo K, Brewer TF, Dibongue E, Diffo J, Kameni JF, Loul S, Nchinda GW, Njouom R, Nwobegahay J, Takuo JM, Torimiro JN, Wade A, Smith TB. Translating Predictions of Zoonotic Viruses for Policymakers. Ecohealth. 2017 Dec 11. PMC29230614.
30. Fatti G, Ngorima-Mabhena N, Chirowa F, Chirwa B, Takarinda K, Tafuma TA, Mahachi N, Chikodzore R, Nyadundu S, Ajayi CA, Mutasa-Apollo T, Mugurungi O, Mothibi E, **Hoffman RM**, Grimwood A. The effectiveness and cost-effectiveness of 3- vs. 6-monthly dispensing of antiretroviral treatment (ART) for stable HIV patients in community ART-refill groups in Zimbabwe: study protocol for a pragmatic, cluster-randomized trial. Trials. 2018 Jan 29;19(1):79. PMC29378662.
31. Caplan MR, Phiri K, Parent J, Phoya A, Schooley A, **Hoffman RM**. Provider Perspectives on Barriers to Reproductive Health Services for HIV-Infected Clients in Central Malawi. Clinical Obstetrics, Gynecology and Reproductive Medicine. 2018 Feb 12;4(1). PMC6391881.
32. Canan T, **Hoffman RM**, Schooley A, Boas Z, Schwab K, Kahn D, Shih R, Phiri K, Parent J, Banda BA, Chagoma R, Chipungu C, Pool K-L. Training Course in Focused Assessment with Sonography for HIV/TB in HIV Prevalent Medical Centers in Malawi. Journal of Global Radiology. 2018 March;4(1). PMC N/A.
33. Rough K, Seage GR, Williams PL, Hernandez-Diaz S, Huo Y, Chadwick EG, Currier JS, **Hoffman RM**, Barr E, Shapiro DE, Patel K; PHACS and the IMPAACT P1025 Study Teams. Birth Outcomes for Pregnant Women with HIV Using Tenofovir–Emtricitabine. New England Journal of Medicine. 2018 April 26;378(17):1593-1603. PMC5984044
34. Dovel K, Shaba F, Nyirenda M, Offorjebe OA, Balakasi K, Phiri K, Nichols B, Tseng C-H, Bardon A, Ngona K, **Hoffman R**. Evaluating the Integration of HIV Self-Testing Into Low-Resource Health Systems: Study Protocol for a Cluster-Randomized Control Trial from EQUIP Innovations. Trials. 2018 Sept 17;19(1). PMC6142354
35. Schwab K, **Hoffman RM**, Phiri L, Kahn D, Longwe L, Banda BA, Gama K, Chimombo M, Shih R, Schooley A, Pool K-L. Remote Training and Oversight of Sonography for Human Immunodeficiency Virus–Associated Tuberculosis in Malawi. Journal of the American College of Radiology. 2018 Sept 26. PMC6540803
36. Hubbard J, Kakwesa G, Nyirenda M, Mwambene J, Bardon A, Balakasi K, Dovel K, Kalua T, **Hoffman RM**. Towards the Third 90: Improving Viral Load Testing with a Simple Quality Improvement Program in Health Facilities in Malawi. Int Health. 2018 Nov 1. PMC Pending
37. Moucheraud C, Chasweka D, Nyirenda M, Schooley A, Dovel K, **Hoffman RM**. Simple Screening Tool to Help Identify High-Risk Children for Targeted HIV Testing in Malawian Inpatient Wards. J Acquir Immune Def Syndr. 2018 Nov 1; 79(3). PMC Pending
38. **Hoffman RM**, Brummel SS, Britto P, Pilotto JH, Masheto G, Aurpibul L, Joao E, Purswani MU, Buschur S, Flore Pierre M, Coletti A, Chakhtoura N, Klingman KL, Currier JS for the 1077HS PROMISE Team. Adverse Pregnancy Outcomes among Women who Conceive on Antiretroviral Therapy. Clin Infect Dis 2018 June 1. PMC6321847
39. Nhassengo P, Cataldo F, Magaco A, **Hoffman RM**, Nerua L, Saide M, Cuco R, Hoek R, Mbofana F, Couto A, Gudo E, Chicumbe S, Dovel K. Barriers and facilitators to the uptake of Test and Treat in Mozambique: a qualitative study on patient and provider perceptions. PLOS One. 2018 Dec 26; 13(12). PMC6306230
40. Magaço A, Dovel K, Cataldo F, Nhassengo P, **Hoffman RM**, Nerua L, Tique J, Saide M, Couto A, Mbofana F, Gudo E, Cuco R, Chicumbe S. ‘Good health’ as a barrier and facilitator to ART initiation: a qualitative study in the era of test-and-treat in Mozambique. Culture, Health and Sexuality. 2018. PMC Pending
41. **Hoffman RM**, Konstantia A, Brummel S, Saidi F, Violari A, Dula D, Mave V, Fairlie L, Gerhard T, Kamateeka M, Chipato T, Chi B, Stranix-Chibanda L, Nematadzira T, Moodley D, Bhattacharya D, Gupta A, Coletti A, McIntyre JA, Klingman KL, Chakhtoura N, Shapiro DE, Folwer MG, Currier JS. Maternal health outcomes among HIV-infected breastfeeding women with high CD4 counts: results of a treatment strategy trial. HIV Clinical Trials 2018 Dec;19 (6). PMC6428202 [Available on 2019-12-01]
42. Murnane PM, Bacchetti P, Currier JS, Brummel S, Okochi H, Phung N, Louie A, Kuncze K, **Hoffman RM**, Nematadzira T, Soko DK, Owor M, Saidi F, Flynn PM, Fowler MG, Gandhi M. Tenofovir concentrations in hair strongly predict virologic suppression in breastfeeding women. AIDS 2019 Apr 22. [Epub ahead of print]. PMC Pending
43. Shaba F, Offorjebe OA, Phiri K, Lungu E, Kalande, P, Nyirenda M, Gupta S, **Hoffman RM**, Dovel K. Perceived acceptability of a facility-based HIV self-test intervention in outpatient waiting spaces among adult outpatients in Malawi: A formative study. J Acquir Immune Defic Syndr. 2019 Jul 1;81(3):e92-e94. PMC Pending
44. Weinberg A, Huo Y, Kacanek D, Patel K, Watts DH, Wara D, **Hoffman RM**, Klawitter J, Christians U; for IMPAACT P1025 Team. Markers of spontaneous preterm delivery in women living with HIV: relationship with protease inhibitors and Vitamin D. J Acquir Immune Defic Syndr. 2019 May 28. [Epub ahead of print]. PMC Pending
45. McBride K, Parent J, Mmanga K, Chivwala M, Nyirenda MH, Schooley A, Mwambene JB, Dovel K, Lungu E, Balakasi K, **Hoffman RM**, Moucheraud C. ART Adherence Among Malawian Youth Enrolled in Teen Clubs: A Retrospective Chart Review. AIDS Behav. 2019 July 10. PMC Pending
46. Le C, Britto P, Brummel SS, **Hoffman RM**, Li JZ, Flynn PM, Taha TE, Coletti A, Fowler MG, Bosch RJ, Gandhi RT, Klingman KL, McIntyre JA, Currier JS. Time to Viral Rebound and Safety after Antiretroviral Treatment Interruption in Postpartum Women Compared to Men. In Press AIDS 2019, August 1. PMC Pending

OMB No. 0925-0001 and 0925-0002 (Rev. 09/17 Approved Through 03/31/2020)

**MICHAL KULICH**

eRA COMMONS USER NAME (credential, e.g., agency login):

POSITION TITLE: Associate Professor of Statistics

EDUCATION/TRAINING (Begin with baccalaureate or other initial professional education, such as nursing, include postdoctoral training and residency training if applicable. Add/delete rows as necessary.)

| INSTITUTION AND LOCATION | DEGREE  (if applicable) | Completion Date  MM/YYYY | FIELD OF STUDY |
| --- | --- | --- | --- |
| Charles University, Prague | M.S. | 9/1991 | Math. Statistics |
| Limburgs Universitair Centrum, Diepenbeek | M.S. | 9/1992 | Biostatistics |
| University of Washington, Seattle | M.S. | 9/1995 | Biostatistics |
| University of Washington, Seattle | Ph.D. | 10/1997 | Biostatistics |

**A.	Personal Statement**

I have an extensive past experience with design, conduct and analysis of clinical trials, especially community randomized trials, in the context of HIV prevention research. I served as the Lead Statistician for the Behavioral Working Group within the HPTN in 2000–2003 and as the Protocol Statistician and Steering Committee member for Project ACCEPT (HPTN043) in 2003-2013. I have been also participating in protocol review groups in the HPTN. I was involved in the design of HPTN043, development of data collection procedures, and development and application of data quality control measures. I am a coauthor of 7 research papers on methodology and results of HPTN043. Since 2015, I am a protocol statstician on another community-randomized trial, Zwakala Ndoda Study: Diagnosing, Linking and Maintaining Men in Antiretroviral Treatment in Vulindlela and Greater Edendale Area, KwaZulu-Natal. The current application builds on my past experience with HIV prevention trials.

**B.	Positions and Honors**

**Professional Positions**

1998–2000	Dept. of Probability and Statistics, Charles University, Prague, Czech Rep., Assistant Professor
2000–2003	Dept. of Biostatistics, University of Washington, Seattle, Research Assistant Professor
2004–2009	Dept. of Probability and Statistics, Charles University, Prague, Czech Rep., Assistant Professor
2010–2013	Dept. of Probability and Statistics, Charles University, Prague, Czech Rep., Associate Professor
2014–	Dept. of Probability and Statistics, Charles University, Prague, Czech Rep., Chair

**Professional Memberships**

1995-	Member, American Statistical Association

1998-	Member, Czech Statistical Society

2004-	Member, International Biometric Society

2005-	Member, International Society for Clinical Biostatistics

**Honors**

1995	Donovan J. Thompson Award, University of Washington, Seattle, WA.

1997	Best Written Paper, International Biometric Society, Park City, UT.

2001	Bolzano Award, Bernard Bolzano Foundation, Prague, Czech Republic.

**C.	Contributions to Science**

**Design and conduct of HIV prevention trials**

I have an expertise in design, conduct and analysis of large randomized HIV prevention trials. I was a protocol statistician in Project ACCEPT (HPTN043), a community-randomized trial conducted in five African and Asian sites, with HIV incidence calculated from cross-sectional blood samples as the primary endpoint. I designed methods for obtaining population samples by household-probability sampling, participated in data verification, performed analyses and collaborated on publications.

Genberg, B., **Kulich, M.,** Kawichai, S., Modiba, P., Chingono, A., Kilonzo, G., Richter, L., Pettifor, A., Sweat, M. & Celentano, D. HIV risk behaviors in Sub-Saharan Africa and Northern Thailand: Baseline behavioral data from Project Accept. *Journal of AIDS* 2008, 49(3):309-319. PMID: 18845954

Sweat, M., Morin, S., Celentano, D., Mulawa, M., Singh, B., Mbwambo, J., Kawichai, S., Chingono, A., Khumalo-Sakutukwa, G., Gray, G., Richter, L., **Kulich, M.,** Sadowski, A., Coates, T., and the Project Accept study team. Community-based intervention to increase HIV testing and case detection in people aged 16-32 years in Tanzania, Zimbabwe, and Thailand (NIMH Project Accept, HPTN 043): a randomised study. *The Lancet Infectious Diseases* 2011, 11(7), 525-532. PMID: 21546309

Coates, T.J., **Kulich, M.**, Celentano, D.D., Zelaya, C.E., Chariyalertsak, S., Chingono, A., Gray, G., Mbwambo, J.K.K., Morin, S.F., Richter, L., Sweat, M., van Rooyen, H., McGrath, N., Fiamma, A., Laeyendecker, O., Piwowar-Manning, E., Szekeres, G., Donnell, D., Eshleman, S.H. (2014) Effect of community-based voluntary counselling and testing on HIV incidence and social and behavioural outcomes (NIMH Project Accept; HPTN 043): A cluster-randomised trial. *The Lancet Global Health* 2014, 2 (5), e267-e277. PMID: 25103167

Salazar-Austin, N., **Kulich, M.**, Chingono, A., Chariyalertsak, S., Srithanaviboonchai, K., Gray, G., Richter, L., van Rooyen, H., Morin, S., Sweat, M., Mbwambo, J., Szekeres, G., Coates, T., Celentano, D. (2017) Age-Related Differences in Socio-Demographic and Behavioral Determinants of HIV Testing and Counseling in HPTN 043/NIMH Project Accept. *AIDS and Behavior* 2018, 22(2) 569-579. PMID: 28589504

**Methods for cross-sectional incidence estimation**

I participated in the development of laboratory and statistical methods for estimating HIV incidence from cross-sectional blood samples. These methods were needed for successful evaluation of the primary outcome in Project ACCEPT.

Laeyendecker, O., Piwowar-Manning, E., Fiamma, A., **Kulich, M.**, Donnell, D., Bassuk, D., Mullis, C. E., Chin, C., Swanson, P., Hackett, Jr, J., Clarke, W., Marzinke, M., Szekeres, G., Gray, G., Richter, L., Alexandre, M. W., Chariyalertsak, S., Chingono, A., Celentano, D. D., Morin, S. F., Sweat, M., Coates, T., Eshleman, S. H. Estimation of HIV Incidence in a Large, Community-Based, Randomized Clinical Trial: NIMH Project Accept (HIV Prevention Trials Network 043), *PLoS ONE* 2013, 8:7, e68349. PMID: 23874597

Laeyendecker, O., **Kulich, M.**, Donnell, D., Komárek, A., Omelka, M., Mullis, C. E., Szekeres, G., Piwowar-Manning, E., Fiamma, A., Gray, R. H., Lutalo, T., Morrison, C. S., Salata, R. A., Chipato, T., Celum, C., Kahle, E. M., Taha, T. E., Kumwenda, N. I., Karim, Q. A., Naranbhai, V., Lingappa, J. R., Sweat, M. D., Coates, T., Eshleman, S. H. Development of Methods for Cross-Sectional HIV Incidence Estimation in a Large, Community Randomized Trial. *PLoS ONE* 2013, 8:11, e78818. PMID: 2423605

Fogel, J.M., Piwowar-Manning, E., Donohue, K., Cummings, V., Marzinke, M.A., Clarke, W., Breaud, A., Fiamma, A., Donnell, D., **Kulich, M.**, Mbwambo, J., Richter, L., Gray, G., Sweat, M., Coates, T., Eshleman, S. Determination of HIV status in African adults with discordant HIV rapid tests. *Journal of Acquired Immune Deficiency Syndromes* 2015, 69, 430-438. PMID: 25835607

Fogel, J.M., Clarke, W., **Kulich, M.**, Piwowar-Manning, E., Breaud, A., Olson, M.T., Marzinke, M.A., Laeyendecker O., Fiamma, A., Donnell, D., Mbwambo, J., Richter, L., Gray, G., Sweat, M., Coates, T.J., Eshleman, S.H. Antiretroviral drug use in a cross-sectional population survey in Africa: NIMH Project Accept (HPTN 043). *Journal of Acquired Immune Deficiency Syndromes* 2017, 74, 158-165. PMID: 27828875

**AUGUSTINE T CHOKO**

**Institution:** Malawi Liverpool Wellcome Trust Clinical Research Programme

**General Medical Council (or equivalent) registration number** N/A

**Do you currently have personal medical malpractice insurance? (if so, name of insurer)**

N/A

**Project role**

Principal Investigator

**Qualifications**

| **Degree** | **Year** | **Subject** | **Awarding Institution** |
| --- | --- | --- | --- |
| PhD | 2018 | Epidemiology | LSHTM |
| MSc | 2012 | Epidemiology | LSHTM |
| BSc | 2009 | Statistics & Computing | University of Malawi |

**Positions held (last ten years)**

| **Start** | **End** | **Organisation** | **Position title, brief description of responsibilities** |
| --- | --- | --- | --- |
| **2020** | **2024** | Malawi Liverpool Wellcome Trust (MLW) | Wellcome Trust & National Institute for Health Research International Intermediate Fellow |
| 2019 | 2020 | MLW | Protocol Lead; leading design, implementation and write up of a complex primary health clinic randomized trial. |
| 2015 | 2018 | MLW | Wellcome Trust Fellow in Public Health and Tropical Medicine   - PhD student |
| 2013 | 2015 | MLW | Research Assistant (Epidemiology)   - Data analysis and publication |
| 2012 | 2013 | MLW | Trial Manager   - Leading implementation of a community-based cluster randomized trial (HIV/TB) |
| 2009 | 2012 | MLW | Data Manager/Statistician   - Designing and administering study databases - Preparing data for analysis and data analysis |

**GCP training: course provider, date**

NIDA Clinical Trials Network, Obtained 11 November 2016, expires 2019

**Please list any institutions with which you are affiliated to**

Malawi Liverpool Wellcome Trust Clinical Research Programme, London School of Hygiene & Tropical Medicine

**Ethics training: details of training, course provider, date**

**Clinical trials experience: title of trial, role, dates (if multiple, restrict to most recent and most relevant)**

Secondary distribution of HIV self-tests through antenatal and HIV testing services: a pragmatic cluster-randomized trial (STAR-ANC); 2018-2019.

Partner-Provided Self-Testing and Linkage (PASTAL) adaptive multi-arm multi-stage cluster randomized trial; 2016-2017.

Intensified HIV / TB prevention linking home-based HIV testing, including the option of self-testing, with HIV care: a cluster-randomised trial in Blantyre, Malaw, Research Fellow, 2011-2015.

**Other professional experience relevant to role in project**

Experience in handling and analyzing large epidemiological datasets.

**Relevant recent publications (maximum 5)**

**Khumbo Phiri Nyirenda**

**CONTACTS**

Partners in Hope

PO Box 302, Lilongwe**.**

Cell: 265882400721/ 265999 840 946

****

**ACADEMIC QUALIFICATIONS**

I. MPH, University of Malawi, College of medicine, anticipating graduation in 2020
II. BSOC, Economics, University of Malawi, Chancellor College- February, 2006
III. Malawi School Certificate of Education (MSCE) Phwezi Girls Sec School-June, 2000

**COURSES/ TRAININGS**

- Qualitative research synthesis, university of cape town, faculty of health sciences February 2020
- Qualitative data analysis university of cape town, faculty of health sciences January 2020
- Biomedical Researchers and Staff modules (CITI program _May 2016)
- Certificate of Attendance in Value chain analysis training by Ron Black from CNFA’s farmer to farmer USAID funded program, Washington DC (February 23-27, 2009)
- Output marketing training in grain grading by CNFA/RUMARK facilitated by a North Carolina Agriculture Department officer, held at Natural Resources College (August 25 -28, 2009).
- Certificate of Attendance in a Leadership workshop facilitated by Engineers without Boarders (October 26 – 28, 2009).
- Training of trainers course in Business management and technical knowledge by COMESA’s ACTESA and IFDC in Lusaka, Zambia (September 6-15, 2010)

**WORK EXPERIENCE**

**PARTNERS IN HOPE**

**Position**: Implementation Science Manager

**Period:** September 2017 to date

Summary:

The Projects Research Coordinator is responsible for overseeing and implementing all research related activities at Partners in Hope (PIH) and in all program-supported sites. He/she is in charge of monitoring and evaluating projects and ensuring that PIH is accountable to research donors. This person works hand-in-hand with the University of California in Los Angeles (UCLA), Partners in Hope (PIH), the Ministry of Health (MoH) and other partners.

Responsibilities

**1. Overseeing and implementing all research projects at Partners in Hope and all EQUIP-supported sites**

**2. Research applications, Reviews and Reports.**

- Oversee applications for ethical review for the Malawi NHSRC and/or COMREC, including initial and renewal applications, as well as closeout of completed projects.
- Serve as the first line of direct communication with the NHSRC and/or COMREC to advocate for submitted applications.
- Work with UCLA, PIH, MoH and other partners to ensure all research is performed to the highest ethical standards and that data is securely managed.
- Make sure appropriate reporting is provided to the governing bodies (final reports, publications, etc.).
- Oversee submission of abstracts to research meetings.

**3. Monitoring, Evaluation and Accountability to Donors**

- Monitor all research projects and develop donor communications in collaboration with senior leadership, especially the M&E Team.
- Ensure timely production and submission of donor experts.
- Participate in development of strategies for expansion of research.
- Ensure continuous evaluation of projects and staff, including hiring and regular appraisals.

**Period**: December 2012 to March 2016

**Organization:** Partners in Hope

**Position:** Research coordinator

**Description**

- Coordinates and administers research study associated activities. Assists in project planning and ensures that pre-established work scope, study protocol and regulatory (ethical review in Malawi and at UCLA) requirements are followed. Oversees and coordinates research staff. Develops and maintains record keeping systems and procedures. My job as Research Coordinator involves these main tasks
- Assistance developing research proposals, data collection forms, and spreadsheets for organization of data. Develops and maintains record keeping systems and procedures.
- Ensures the smooth and efficient day to day operation of research and data collection
- activities; acts as the primary administrative point of contact for EQUIP research staff.
- Supervision of team of research assistants
- Assistance with recruitment and coordination of research subjects as appropriate.
- Supervision and assistance with quality control Data.
- Monitors the progress of research activities; develops and maintains records of research activities and prepares periodic and ad hoc reports as required by investigators, administrators and funding agencies(USAID quarterly reports) and regulatory bodies (NHSRC,UCLA,IRB)
- Assistance with preparing ethical review applications (HSRC) including frequent communication with NHSRC about status of pending applications.

**I. CARANA COOPERATION**

**Position:** M&E/MIS Assistant

**Period:** November 2010–September 2011

**Description**

Market Linkages Initiative was a project funded by **USAID** and implemented by ACDI/VOCA and CARANA Corporation. The two key objectives of the project are to strengthen and expand grain bulking systems and to integrate farmers to national and regional markets. My job as an M&E/MIS Specialist involved these main tasks:

- Assisting the M&E specialist in tracking MLI indicators, collecting and verifying information and maintaining PMP reports, work plans and reports(weekly, monthly updates, quarterly and annual) for Malawi activities
- Administering data collection tools to GBC/VACs and capacity building of grantees to keep relevant records and generate M&E reports as stipulated by the Grant agreement.
- Collating , analyzing and reporting in usable forms all data collected form GBC/VAC
- Supporting in the coming up of GIS map for MLI supported GBCs and its associated VACs in Malawi
- Administering M&E data collection tools and supervising M&E data collectors and ensure quality data collection
- Maintaining records of all source documents from grantees and other sources including filled questionnaires and interview reports
- Keeping records of field trip reports and monitor and updating field trip tracker for Malawi based MLI staff
- Undertaking case studies and documenting most significant change stories for selected GBCs/VACs/Farmers to monitor impacts of MLI work
- Maintaining an up to date filing system including project photos
- Ensuring that quality control procedures are met in terms of market data.
- Facilitating dissemination of market information to farmers on a timely and reliable basis using the E-platform
- Providing technical assistance on the E- platform to strategic partners
- Working alongside the new company and assisting/participating in development and deployment team to design and roll out a web to phone MIS platform
- Conducting weekly data checks on approved prices inside E-platform’s price flagging module
- Manage a user, market and commodities database

II. **CNFA/RUMARK**

**Position:** Monitoring and Evaluation Coordinator

**Period:** January 2009 to October 2010

CNFA/RUMARK implemented the Malawi Agrodealer Strengthening Program funded by AGRA. Its main objective was to develop rural-based, commercially-viable agrodealer networks and to work with agrodealers to improve the management, technical and financial capacity of their enterprises, thereby creating a rural market driven economic environment specifically designed to meet the unique needs of smallholder farmers. My position of as M&E coordinator involved the following tasks

- Monitoring progress of the project activities by doing surveys which included development of survey tools which mostly use participatory methods.
- Monitoring and evaluating the agrodealers performance in terms of sales as well as their financial status.
- Analysis of information on Agrodealer performance
- Verifying and identifying operational and potential agrodealers and recommending them for training to enable them get registered with CNFA
- Organizing promotional activities i.e. lottery competitions with the intention of creating customer database for surveys
- Playing a facilitating role in managing relationships between RUMARK and input supply companies to ensure cordial relationships and partnerships.
- Consolidating and analyzing results across CNFA’s programs.
- Conducting training needs assessment for different categories of agrodealers to ensure equal treatment so that their specific needs are taken on board.
- Organizing the Agrodealers Annual Convention.
- Production of monthly as well as interim semi-annual reports for the Project
- Involved in advocating for policies which are conducive for agrodealers’ business growth and sustainability through Private–public partnerships which involves working with various stakeholders including Government and civil society organizations.

**Research Abstracts**

**Provider perspectives on barriers to reproductive health services for HIV-infected clients in Central Malawi**: Khumbo Phiri, Margaret R Caplan , Julie Parent , Ann Phoya , Alan Schooley, and Risa M. Hoffman, Poster presentation at Interest 2017, Malawi

**Barriers to ART uptake experienced by healthy clients in Malawi under Test and Treat:** Dovel, Kathryn, Khumbo Phiri, Alan Schooley, McDaphton Bellos, Esnart Sanudi, Denis Chasweka, Risa Hoffman, poster exhibition at the 9th IAS Conference on HIV Science (IAS 2017, in Paris, France, 23-26 July 2017 and Interest 2017 in Malawi).

**Facility-level barriers to antiretroviral therapy experienced by men in Malawi:** Dovel, Kathryn, Khumbo Phiri, Alan Schooley, Misheck Mphande, Mackenzie Chivwara, Risa Hoffman(poster presentation at interest 2017, Malawi)

**Examining Malawi’s Rollout of Universal Treatment: Policy Implementation and Provider Perceptions**: Misheck Mphande, Khumbo Phiri, Mackenzie Chivwara, Mike Nyirenda, Alan Schooley, Rachel Thomas, Risa Hoffman, Kathryn Dovel, (Poster presentation at IAS 2017 in Paris, France)

**Low rates of successful defaulter tracing and re-engagement in care in Option B+ women in Central Malawi.** K. Phiri, J. Parent, T. Mulitswa, A. Schooley, R. Hoffman. Poster presentation at the International AIDS Society (IAS) conference (Durban, 2016).

**The successes and Challenges of collaborating with Health Surveillance Assistants (HSAs) to trace Option B+ defaulters.** Khumbo Phiri Nyirenda, Julie Parent, Risa Hoffman, Alan Schooley, Temwanani Mulitswa

**The Option B+ cascade: Characterizing uptake and retention in a USAID-PEPFAR program in rural Malawi.** Khumbo Phiri, Alan Schooley, Mackenzie Chivwala, Joseph Njala, Judy Currier, Andreas Jahn, Anteneh Worku, Perry Jansen, Risa Hoffman

**Improvements and on-going challenges in exposed infants care at rural sites in Malawi.** Alan Schooley, Khumbo Phiri, Mackenzie Chivwala, Peter Chilikoh, Antenneh worku, Risa Hoffman

**Health Surveillance Assistants Can Successfully Perform Defaulter Tracing In Rural Malawi.** Mackenzie Chivwala, Khumbo Phiri, Risa Hoffman, Jimmy Chitsulo, Alan Schooley

**Assessing the Potential Impact of Health Surveillance Assistants on HIV Care At The Facility And Community Level.** Mackenzie Chivwala, Khumbo Phiri, Weston Njamwaha, Peter Chilikoh, Risa Hoffman, Alan Schooley

**Mentee Perspectives on Factors Associated with a Successful HIV Mentorship Program** Mike Nyrienda, Chiulemu Kussen, Savior Mwandira, Khumbo Phiri, Chiukepo Longwe, Peter Chilikoh, Risa Hoffman, Weston Njamwaha, Alan Schooley

**Rapid Rollout of Viral Load Testing at Rural Health Facilities in Malawi.** Alan Schooley, Risa Hoffman, Mike Nyirenda, Savior Mwandira, Weston Njamwaha, Khumbo Phiri, Chifundo Chipungu, Mackenzie Chivwala, James Kandulu

**Increased HIV testing after implementation of an innovative CD4 results reporting system in rural Malawi.** Alan Schooley, Mackenzie Chivwala, Reynier Ter Haar, George Mtonga, Doreen Suwande, Kelvin Rambiki, Chiulemu Kussen, John Hamilton, Khumbo Phiri, Peter Chilikoh, Risa Hoffman, Perry Jansen. Accepted for poster presentation at the 6th South African AIDS Conference, Durban, South Africa, 18-21 June 2013.

**Barriers to Adherence to ART in the Prevention of Mother-to-Child Transmission of HIV: Option B+ in Nkhoma, Malawi.** Paul Kawale, Alan Schooley, Virginia Tancioco, Danielle Wickman, Khumbo Phiri, Ella Bwanausi, Risa Hoffman. Accepted for poster presentation at the 6th South African AIDS Conference, Durban, South Africa, 18-21 June 2013.

**MANUSCRIPTS ACCEPTED/PUBLISHED**

**Successes and Challenges of HIV Mentoring in Malawi: The Mentee Perspective.** E. Chien, K. Phiri, A. Schooley, M. Chivwala, J. Hamilton, R. Hoffman. PLoS One. 2016 Jun;11(6).

**CD4 variability in Malawian adults and implications for universal eligibility.** A.L. Schooley, P.S. Kamudumuli, S. Vangala, C.H. Tseng, C. Soko, J. Parent, K. Phiri, A. Jahn, D. Namarika, R. Hoffman. Open Forum Infect Dis. 2016 Aug;3(3).

**Provider perspectives on barriers to reproductive health services for HIV-infected clients in Central Malawi:** Margaret R Caplan, Khumbo Phiri, Julie Parent, Ann Phoya , Alan Schooley, and Risa M. Hoffman, PLOS ONE.

**Factors Associated with Retention in Option B+ in Malawi: A Case Control Study:** Risa M. Hoffman, khumbo phiri, Julie parent, J Grotts D Elashoff, Paul Kawale, Sara Yeatman, J S Currier, A Schooley, JIAS.

**Training Course in Focused Assessment with Sonography for HIV/TB in HIV Prevalent Medical Centers in Malawi:** Timothy Canan, R Hoffman, Alan Schooley, Zachary Boas, Kristin Schwab, Daniel Kahn, Roger Shih, Khumbo Phiri, Julie Parent, Ben Allan Banda, Ronald Chagoma, Chifundo Chipungu. Kara-Lee Pool, Journal of Global Radiology

**REFEREES**

**Risa Hoffman (MD),** Assistant Clinical Professor**,** David Gaffen school of medicine, UCLA**,**

**Rachel Sibande (PhD),** Program Director, United Nations Foundation, +27670236497

**Godfrey Chapola (PhD),** Managing Director, RUMARK, P.O Box 31290, Lilongwe.

Cel: 0999792 070,

**Julie Hubbard**

94 Culford Road London N1 4HN

+44 7845 445338

**EDUCATION**

London School of Hygiene and Tropical Medicine

MSc Control of Infectious Diseases

Seattle Pacific University

Bachelor of Arts: Sociology & Women’s Studies

Cum Laude GPA: 3.74

Graduated July 2012

**PROFESSIONAL EXPERIENCE**

University of California Los Angeles (UCLA), March 2017- Current

Research Coordinator – ‘INTERVAL’ Study

Lilongwe, Malawi and Lusaka, Zambia

•	Supervise data collection by study personnel across 15 health facilities in southern and central Malawi. Coordinate field supervision to ensure data quality. Work with Principle Investigator (PI) to develop operating procedures for study implementation. Provide leadership and technical support to Zambia study team.

Harvest India USA, January 2016 – March 2017

Director of Operations

Costa Mesa, California and Andhrah Pradesh, India

•	Managed all aspects of operations to support, fundraise, and raise awareness for education and poverty alleviation initiatives amongst the Dalit, or ‘untouchable’, caste. Drafted and executed marketing campaigns to meet fundraising goals.

31 Bits International, December 2012 – February 2015

Director of Operations

Gulu, Northern Uganda

•	Directed 160 beneficiaries and 6 Ugandan counselors in income generating projects. Developed and implemented in-depth monthly reports to evaluate income. Used data to identify hindrances to livelihood, such as domestic violence and HIV health complications. Organized necessary support through internal management or accessing external resources.

One Days Wages, March 2011- December 2012

Chief Grant Analyst

Seattle, Washington

•	Generated extensive research on project proposals pertaining to the UN Millennium Development Goals and presented analyses for grant decisions.

Seattle Pacific University, September 2011- July 2012

Research Assistant

Seattle, Washington

•	Edited, reviewed, and prepared research documents for Assistant Director of Women’s Studies Program.

**PUBLICATIONS AND PRESENTATIONS**

*Publications*

Julie Hubbard, Gift Kakwesa, Mike Nyirenda, James Mwambene, Ashley Bardon, Kelvin Balakasi, Kathryn Dovel, Thokozani Kalua, Risa M Hoffman; Towards the third 90: improving viral load testing with a simple quality improvement program in health facilities in Malawi, International Health, , ihy083, <https://doi.org/10.1093/inthealth/ihy083>

Hubbard J, Moucheraud C, Lungu E, Bardon A, Balakasi K, Kakwesa G, Hoffman R ““I forget that I am a patient”: A qualitative assessment of 6 month dispensing of ART” (Under review)

Dovel K, Beagley M, Hubbard J, Orombi G, Thompson K “Including men without sidelining women: the feasibility of male involvement within a women’s empowerment program in northern Uganda” (Under review)

Dovel K, Hubbard J, Phiri K. “Gender and HIV services: The role of gender norms on ART initiation among men and women in Malawi.” (In preparation)

Peer reviewed poster presentations

“Gender and HIV services: The role of gender norms on ART initiation among men and women in Malawi.” Women in Global Health Scientific Conference. New York, New York. April 2018

“Towards the third 90: improving viral load testing with a simple quality improvement program in health facilities in Malawi” International Aids Society (IAS) Conference, Amsterdam, Netherlands. July 2018

*Presentations*

“Innovations in differentiated service delivery: Six-month scripting lessons from Ethiopia, Malawi and Zambia” Colombia University Mailman School of Public Health. Webinar, April 2019

**CERTIFICATIONS**

Confronting Gender Based Violence: Global Lessons with Case Studies from India Certification Course Coursera (Johns Hopkins University) - Online

October 2015

•	Epidemiology of gender-based violence, clinical care issues and how to provide psychosocial support for victims.

**FELLOWSHIP**

Mennonite Central Committee

Community Development Associate, July-August 2011

•	Rural and urban poverty field study in Recife, Brazil association under the direction of the Chair of the Sociology Department at Seattle Pacific University.

**HONORS**

Seattle Pacific University Deans Scholar, 2008-2012

**BROOKE E. NICHOLS, PHD, MS**

| **OFFICE ADDRESS:** |  | 801 Massachusetts Avenue | |
| --- | --- | --- | --- |
|  |  | Crosstown Center, 3rd Floor, Room 304 | |
|  |  | Boston, MA, USA 02118 | |
|  |  | +1 617 358 2403 | |
|  |  | | |
| **ACADEMIC TRAINING:** | |  |  |
| 2015 | Ph.D. |  | Department of Viroscience, Erasmus Medical Center (Rotterdam, the Netherlands) |
|  |  |  | *Mathematical Modeling and Cost-Effectiveness of Antiretroviral-Based HIV-1 Prevention Strategies.* |
| 2011 | M.S. |  | School of Public Health & Health Science, University of Massachusetts, Amherst |
|  |  |  | (Amherst, MA, USA), Epidemiology |
| 2009 | B.A. |  | Mount Holyoke College (South Hadley, MA, USA), International Relations, *cum laude* |

| **RESEARCH APPOINTMENTS:** | | |  |
| --- | --- | --- | --- |
| 2019 | – Present | Assistant Professor | Department of Global Health, School of Public Health, |
|  |  |  | Boston University, Boston, MA |
| 2018 | – 2019 | Instructor | Department of Global Health, School of Public Health, |
|  |  |  | Boston University, Boston, MA |
| 2017 | – 2018 | Research Scientist | Department of Global Health, School of Public Health, |
|  |  |  | Boston University, Boston, MA |
| 2017 | – Present | Principal Researcher | Health Economics & Epidemiology Research Office, Wits |
|  |  |  | Health Consortium, Faculty of Health Sciences, University of |
|  |  |  | Witwatersrand, Johannesburg, South Africa |
| 2017 | – Present | Researcher | Joint Faculty Appointment, School of Clinical Medicine, |
|  |  |  | Faculty of Health Sciences, University of the Witwatersrand, |
|  |  |  | Johannesburg, South Africa |
| 2015 | – 2016 | Postdoctoral Fellow | Department of Viroscience, Erasmus Medical Center, |
|  |  |  | Rotterdam, the Netherlands |
| 2009 | – 2010 | Research Assistant | University of Massachusetts Amherst, School of Public Health |
|  |  |  | & Health Sciences, Amherst, MA |
| **OTHER RESEARCH EXPERIENCE:** | | |  |
| 2012 | – 2014 | Epidemiologist | Médicins Sans Frontières |
|  |  |  | Amsterdam, the Netherlands. |
|  |  |  | Project: *Spinal cord injury outcomes in Sri Lanka* |
| 2008 | – 2009 | Researcher | Ministry of Health and Social Services, Lüderitz, Namibia |
|  |  |  | Project: *Ecologic study on alcohol establishments and HIV prevalence* |
| **PROFESSIONAL APPOINTMENTS:** | | |  |
| 2008 | – 2010 | Research Associate: Epidemiology | Environ Corporation, Amherst, MA, USA. |
| **AWARDS AND HONOURS:** | | **Mary Lyon Award.** Mount Holyoke College Alumni Association. Award honors an alumna who | |
| 2019 |  |  |  |
|  |  | has demonstrated sustained achievement in her life and career consistent with the humane values | |
|  |  | that Mary Lyon exemplified and inspired in others. | |

**CONFERENCE ORAL PRESENTATIONS:**

*Denotes graduate student or mentee

1. Popping S*, Kall M, Stempher E, Versteegh L, **Nichols B**, van Sighem A, van de Vijver D, Boucher C, Verbon A, Delpech V. Country specific factors determine the quality of life among people with HIV in two western European countries.*:* 4th European Workshop on Health Living with HIV, Barcelona, Spain, September 2019.
2. Dovel K, Balakasi K, Shaba F, Offorjebe O, Gupta S, Wong S, Phiri K, Lungu E, Nyirenda M, **Nichols B**, Ngona K, Hoffman R. A randomized trial on index HIV self-testing for partners of ART clients in Malawi. Conference on Retroviruses and Opportunistic Infections (CROI), Seattle, USA, March 2019.
3. **Nichols BE**, Girdwood SJ*, Crompton T, Stewart-Isherwood L, Berrie L, Chimhamhiwa D, Moyo C, Kuehnle J, Rosen S. Monitoring viral load for the last mile: what will it cost? AIDS, Amsterdam, Netherlands, July 2018.
4. Girdwood SJ*, **Nichols BE**, Moyo C, Crompton T, Chimhamhiwa D, Rosen S. Optimizing access for the last mile: Geospatial cost model for point of care viral load instrument placement in Zambia. AIDS, Amsterdam, Netherlands, July 2018.
5. Dovel K, Nyirenda M, Shaba F, Offorjebe OA, Balakasi K, **Nichols BE**, Phiri K, Schooley A, Hoffman RM. Facility-based HIV self-testing for outpatients dramatically increases HIV testing in Malawi: a cluster randomized trial. AIDS, Amsterdam, Netherlands, July 2018.
6. **Nichols BE**, Hendrickson C, Sigwebela N, Moyo C, Fox MP, Rosen S. Prioritizing healthcare facilities for on-site mentorship to increase HIV treatment uptake: results from EQUIP. International AIDS Economics Network (IAEN) Conference, Amsterdam, Netherlands, July 2018.
7. van de Vijver DA, Richter A-K, Boucher CA, Gunsenheimer-Bartmeyer B, Kollan C, **Nichols BE**, Spinner C, Wasem J, Schewe K, Neumann A. Cost-effectiveness of pre-exposure prophylaxis in Germany (Kosteneffektivität der HIV-Präexpositionsprophylaxe in Deutschland). DGGÖ (German Society for health economics) Annual Meeting, Hamburg, Germay, March 2018.
8. van de Vijver DA, Richter A-K, Boucher CA, Gunsenheimer-Bartmeyer B, Kollan C, **Nichols BE**, Spinner C, Wasem J, Schewe K, Neumann A. Cost-effectiveness of pre-exposure prophylaxis for HIV-1 prevention in Germany. European AIDS Conference (EACS), Milan, Italy, October 2017.
9. Smit M, van Zoest RA, **Nichols BE**, Vaartjes I, Smit C, van der Valk M, van Sighem A, Wit FW, Hallett TB, Reiss P. Cardiovascular prevention policy in HIV: recommendations from a modeling study. Conference on Retroviruses and Opportunistic Infections (CROI), Seattle, WA. February 2017.
10. Popping S*, **Nichols BE**, van Kampen JJA, Verbon A, Boucher CAB, van de Vijver DA. Intensive hepatitis C monitoring in previously HCV infected HIV-positive MSM is a cost saving method to reduce the HCV epidemic. Netherlands Conference on HIV Pathogenesis, Epidemiology, Prevention and Treatment (NCHIV), Amsterdam, the Netherlands, November 2016.
11. **Nichols BE**, Boucher CAB, van der Valk M, Rijnders BJA, van de Vijver DA. PrEP is Only Cost-Effective Among MSM in the Netherlands When Used on Demand. Conference on Retroviruses and Opportunistic Infections (CROI), Boston, MA. February 2016.
12. **Nichols BE**, Boucher CAB, van der Valk M, Rijnders BJA, van de Vijver DA. On demand PrEP among MSM in the Netherlands: a cost-effective approach for preventing HIV-1 infections. Netherlands Conference on HIV

**Peer reviewed publications:**

*Authors contributed equally

**Denotes graduate student or mentee

1. Dovel K, Nyirenda M, Shaba F, Offorjebe OA, Balakaksi K, **Nichols BE**, Cele R. Phiri K, Wong V, Gupta S, Hoffman RM. Effect of facility-based HIV self-testing on uptake of testing among adult outpatients in Malawi: a cluster-randomized trial. *The Lancet Global Health. In press.*
2. van Vliet MM**, Hendrickson C**, **Nichols BE**, Boucher CAB, Peters RPH, Polis CB, van de Vijver DAMC. Epidemiological impact and cost-effectiveness of long-acting pre-exposure prophylaxis combined with injectable contraceptives for HIV prevention in South Africa: a modelling study. *JIAS.* 2019, 22:e25427.
3. Long, L., Kuchukhidze, S., Pascoe, S., **Nichols, B.,** Cele R., Govathson, C., Flynn, D., Rosen, S. Differentiated Models of Service Delivery for Antiretroviral Treatment of HIV in sub-Saharan Africa: A Rapid Review Protocol. *Systematic* *Reviews*. 2019, 8:314.
4. Masuku S**, Berhanu R, van Rensburg C, Ndjeka N, Rosen S, Long L, Evans D, **Nichols BE**. The costs of managing multi drug-resistant tuberculosis in South Africa: an economic evaluation of moving to a short-course treatment regimen containing bedaquiline. *International Journal of Tuberculosis and Lung Disease. In press.*
5. Hendrickson C*,**, Long L*, van de Vijver DA, Boucher CA, O’Bra H, Claassen CW, Njelesani M, Moyo C, Mumba DB, Subedar H, Mulenga L, Rosen S, **Nichols BE**. Novel metric for evaluating PrEP program effectiveness in real-world settings. *Lancet HIV*. *In press.*
6. Girdwood SJ**, **Nichols BE,** Moyo C, Crompton T, Chimhamhiwa D, Rosen S. Optimizing access for the last mile: Geospatial cost model for point of care viral load instrument placement. *PLoS ONE.* 14(8):e0221586.
7. **Nichols BE**, Girdwood SJ**, Crompton T, Stewart-Isherwood L, Berrie L, Chimhamhiwa D, Moyo C, Kuehnle J, Stevens W, Rosen S. Monitoring viral load for the last mile: what will it cost? *JIAS.* 2019, 22:e25337.
8. Popping S**, **Nichols BE**, van Kampen JJA, Verbon A, Boucher CAB, van de Vijver DA. Targeted HCV core antigen monitoring among HIV-positive men-who-have-sex-with-men is cost-saving. *Journal of Virus Eradication.* 2019; 5:179-190.
9. **Nichols BE**, Girdwood SJ**, Shibemba A, Sikota S, Gill C, Mwananyanda L, Scott L, Noble L, Carmona S, Rosen S, Stevens W. Cost and impact of dried blood spot versus plasma separation card for viral load testing in resource limited settings. *Clinical Infectious Diseases.* Advance article: 10.1093/cid/ciz338.
10. van de Vijver DA, Richter A-K, Boucher CA, Gusenheimer-Bartmeyer B, Kollan C, **Nichols BE**, Spinner CD, Wasem J, Schewe K, Neumann A. Cost-effectiveness and budget impact of generic pre-exposure prophylaxis for HIV-1 prevention in Germany. *Eurosurveillance*. 2019 Feb; 24(7).
11. Popping S**, Hulligie SJ, Boerekamps A, Rijnders BJA, de Knegt RJ, Rockstroh JK, Verbon A, Boucher CAB, **Nichols** **BE**, van de Vijver DA. Early treatment of acute HCV infection is cost-effective in HIV-infected men-who-have-sex-with-men. *PLoS One*, 2019. 14(1):e0210179.
12. **Nichols BE**, Girdwood SJ**, Crompton T, Stewart-Isherwood L, Berrie L, Chimhamhiwa D, Moyo C, Kuehnle J, Stevens W, Rosen S. Impact of a borderless sample transport network for scaling up viral load monitoring: results of a geospatial optimization model for Zambia. *JIAS.* 2018, 21:e25206.
13. Dovel K, Shaba F, Nyirenda M, Ogechukwu AO, Balakasi K, Phiri K, **Nichols BE**, Tseng C-H, Bardon A, Namachapa KN, Hoffman RM. Evaluating the integration of HIV self-testing into low-resource health systems: a study protocol for a cluster randomized control trial from EQUIP Innovations. *Trials,* 2018. 19:498.
14. Phillips AN, Cambiano V, Nakagawa F, Revill P, Jordan MR, Hallett TB, Doherty M, De Luca A, Lundgren JD, Mhangara M, Apollo T, Mellors J, **Nichols B**, Parikh U, Pillay D, Rinke de Wit T, Sigaloff K, Havlir D, Kuritzkes DR, Pozniak A, van de Vijver D, Vitoria M, Wainberg MA, Raizes E, Bertagnolio S, Working Group on Modelling Potential Responses to High Levels of Pre-ART Drug Resistance in Sub-Saharan Africa. Cost-effectiveness of public-health policy options in the presence of pretreatment NNRTI drug resistance in sub-Saharan: a modelling study. *Lancet HIV*, 2018. 5(3):e146-e154.
15. Smit M, van Zoest RA, **Nichols BE**, Vaartjes I, Smit C, van der Valk M, van Sighem A, Wit FW, Hallett TB, Reiss P; Netherlands ATHENA observational HIV cohort. Cardiovascular disease prevention policy in HIV: recommendations from a modelling study. *Clinical Infectious Diseases*, 2018. 66(5):743-750.
16. Luiken GPM, Joore IK, Taselaar A, Schuit SCE, Geerlings SE, Govers A, Rood PPM, Prins JM, **Nichols BE**, Verbon A, de Vries-Sluijs TEMS. Non-targeted HIV screening in emergency departments in the Netherlands. *The Netherlands* *Journal of Medicine*, 2017. 75(9):386-393.
17. **Nichols BE**, Boucher CAB, van der Valk M, Rijnders BJA, van de Vijver DAMC. Cost-effectiveness analysis of pre-exposure prophylaxis for HIV-1 prevention in the Netherlands: a mathematical modelling study. *Lancet Infectious Diseases*, 2016. 16(12):1423-1429.
18. **Working Group on Modelling of ART Monitoring Strategies in Sub-Saharan Africa**. Sustainable HIV Treatment in Africa through Viral Load-Informed Differentiated Care. *Nature*, 2015. 528(7580):S68-76.
19. **Nichols BE**, Gotz HM, van Gorp ECM, Verbon A, Rokx C, Boucher CAB, van de Vijver DAMC. Partner notification for reduction of HIV-1 transmission and related costs among men who have sex with men: a mathematical modeling study. *PLoS One,* 2015. 10(11):e0142576.
20. **Nichols BE**, Sigaloff KC, Kityo C, Hamers RL, Baltussen R, Bertagnolio S, Jordan MR, Hallett TB, Boucher CA, Rinke de Wit TF, van de Vijver DA. Increasing the use of second-line therapy is a cost-effective approach to prevent the spread of drug-resistant HIV: a mathematical modelling study. *J Int AIDS Soc*, 2014. 17:19164.
21. **Nichols BE**, Baltussen R, van Dijk JH, Thuma PE, Nouwen JL, Boucher CA, van de Vijver DA. Cost-effectiveness of PrEP in HIV/AIDS control in Zambia: a stochastic league approach. *JAIDS*, 2014. 66(2):221-8.
22. Eaton JW, Menzies NA, Stover J, Cambiano V, Chindelevitch L, Cori A, Hontelez JAC, Humair S, Kerr CC, Klein DJ, Mishra S, Mitchell KM, **Nichols BE**, Vickerman P, Bakker R, Barnighausen T… Hallett TB. How should HIV programmes respond to evidence for the benefit of earlier treatment initiation? A combined analysis of twelve mathematical models. *Lancet Global Health*, 2014. 2:e23-34.
23. Armstrong JC*, **Nichols BE***, Wilson JM, Cosico RA, Shanks L. Spinal cord injury in the emergency context: review of program outcomes of a spinal cord injury rehabilitation program in Sri Lanka. *Conflict and Health*, 2014. 8(1):4.
24. **Nichols BE,** Sigaloff KC, Kityo C, Mandaliya K, Hamers RL, Bertagnolio S, Jordan MR, Boucher CA, Rinke de Wit TF, van de Vijver DA. Averted HIV infections due to expanded antiretroviral treatment eligibility offsets risk of transmitted drug resistance: A modeling study. *AIDS*, 2014. 28(1):73-83.
25. van de Vijver DA, **Nichols BE,** Abbas UL, Boucher CA, Cambiano V, Eaton JW, Glaubius R, Lythgoe K, Mellors J, Phillips A, Sigaloff KC, Hallett TB. Pre-Exposure Prophylaxis will have a limited impact on HIV-1 drug resistance in sub-Saharan Africa: A comparison of mathematical models. *AIDS*, 2013. 27(18):2943-2951.

**Alemayehu Amberbir**

POSITION TITLE: Epidemiologist

eRA Commons User Name: A.AMBERBIR_DI

EDUCATION/TRAINING

|  | DEGREE |  |  |
| --- | --- | --- | --- |
| INSTITUTION AND LOCATION | *(if* | YEAR(s) | FIELD OF STUDY |
|  | *applicable)* |  |  |
| Haramaya University, Ethiopia | BSc | 2004 | Health Officer |
| Jimma University, Ethiopia | MPH | 2007 | Public Health |
| University of Nottingham, UK | PhD | 2012 | Epidemiology |

**Positions and Honors**

Sept 2003 – Jun 2004 Intern (Clinical/Public Health), Haramaya University & Hiwot Fana Hospital, Ethiopia Oct 2004 – Sept 2005 HIV/AIDS Prevention and Care Program Officer, Menschen Für Menschen, Ethiopia

Aug 2007 - Mar 2008	Lecturer, Department of Epidemiology and Biostatistics, Jimma University, Ethiopia

Feb 2008 – Mar 2011	Honorary Lecturer, Addis Ababa University, Ethiopia

Feb 2008 – Feb 2012	Research Fellow, University of Nottingham, UK

Mar 2012 – Sep 2013	Research Fellow, London School of Hygiene and Tropical Medicine, UK

Oct 2013 – Jan 2016	Lecturer, London School of Hygiene and Tropical Medicine, UK

Jan 2016 – Jun 2019	Epidemiologist, Dignitas International

Jan 2018 – Jun 2019	Adjunct Lecturer, Dalla Lana School of Public Health, University of Toronto

Jan 2018 – Dec 2019	Postdoctoral Fellow; CIHR Canadian HIV Trials Network (CTN), Canada

Aug 2019 – present	Science Director, University of California Los Angles; David Geffen School of Medicine

**Contribution to Science**

*Investigating non-communicable diseases (hypertension, diabetes and asthma) in Africa (selected)*

*HIV risk behaviours and status disclosure in African settings (selected)*

1) Dessalegn NG, Hailemichael RG, Shewa-Amare A, Sawleshwarkar S, Lodebo B, **Amberbir A**, Hillman RJ. [HIV Disclosure: HIV-positive status disclosure to sexual partners among individuals receiving HIV care in](https://www.ncbi.nlm.nih.gov/pubmed/30768642) [Addis Ababa, Ethiopia.](https://www.ncbi.nlm.nih.gov/pubmed/30768642) PLoS One**. 2019** Feb 15;14(2):e0211967. doi: 10.1371/journal.pone.0211967. eCollection 2019.
2) Deribe K, Woldemichael K, Wondafrash M, Haile A, **Amberbir A**. [Disclosure experience and associated](http://www.ncbi.nlm.nih.gov/pubmed/18312653) [factors among HIV positive men and women clinical service users in Southwest Ethiopia.](http://www.ncbi.nlm.nih.gov/pubmed/18312653) *BMC* *Public* *Health*. 2008 Feb 29;8:81. doi: 10.1186/1471-2458-8-81.
3) Biadgilign S, Deribew A, **Amberbir A**, Escudero HR, Deribe K. [Factors associated with HIV/AIDS](http://www.ncbi.nlm.nih.gov/pubmed/21445289) [diagnostic disclosure to HIV infected children receiving HAART: a multi-center study in Addis Ababa,](http://www.ncbi.nlm.nih.gov/pubmed/21445289) [Ethiopia.](http://www.ncbi.nlm.nih.gov/pubmed/21445289) *PLoS* *ONE*. 2011 Mar 21;6(3):e17572. doi: 10.1371/journal.pone.0017572.

*Health system research in to infectious diseases – HIV – in Africa (selected)*

1. Alhaj M^1,2¶^,**Amberbir A**^1¶^, Singogo E, Banda V, van Lettow M, Matengeni A, Kawalazira G, Theu J, Jagriti MR, Chan AK, van Oosterhout JJ. Retention on antiretroviral therapy during universal test and treat implementation in Zomba district, Malawi: a retrospective cohort study. J Int AIDS Soc. 2019 Feb;22(2):e25239. doi: 10.1002/jia2.25239.
2. Singano V^¶^, **Amberbir A**^¶^, Garone D, Kandionamaso C, Msonko J; van Lettow M, Kalima K, Mataka, Kawalazira G, Mateyu G, Kwekwesa A, Matengeni A; van Oosterhout JJ. The burden of gynecomastia among men on Antiretroviral Therapy in Zomba, Malawi. *PLoS ONE 12(11): e0188379:* *doi.org/10.1371/journal.pone.0188379*
3. Mpawa H, Kwekwesa A, **Amberbir A**, Garone D, Divala OH, Kawalazira G, van Schoor V , Ndindi H, van Oosterhout JJ. Virological outcomes of Antiretroviral Therapy in Zomba Central Prison, Malawi: *J Int AIDS* *Soc* 2017, 20:21623
4. **Amberbir A**, Woldemichael K, Getachew S, Girma B, Deribe K. [Predictors of adherence to antiretroviral](http://www.ncbi.nlm.nih.gov/pubmed/18667066) [therapy among HIV-infected persons: a prospective study in Southwest Ethiopia.](http://www.ncbi.nlm.nih.gov/pubmed/18667066) *BMC* *Public Health***. 2008** Jul 30;8:265. doi: 10.1186/1471-2458-8-265.
5. Biadgilign S, Deribew A, **Amberbir A**, Deribe K. [Barriers and facilitators to antiretroviral medication](http://www.ncbi.nlm.nih.gov/pubmed/20485854) [adherence among HIV-infected paediatric patients in Ethiopia: A qualitative study.](http://www.ncbi.nlm.nih.gov/pubmed/20485854) *SAHARA* *J***.** 2009 Dec;6(4):148-54.
6. Deribe K, Hailekiros F, Biadgilign S, **Amberbir A**, Beyene BK. [Defaulters from antiretroviral treatment in](http://www.ncbi.nlm.nih.gov/pubmed/18298607) [Jimma University Specialized Hospital, Southwest Ethiopia.](http://www.ncbi.nlm.nih.gov/pubmed/18298607) *Trop* *Med Int Health*. 2008 Mar;13(3):328-33. doi: 10.1111/j.1365-3156.2008.02006.x. Epub 2008 Feb 21.

**LAWRENCE C. LONG, PHD, MCOM**

| **OFFICE ADDRESS:** | | 801 Massachusetts Avenue, | | | |  |
| --- | --- | --- | --- | --- | --- | --- |
|  |  | Crosstown Center, 3rd Floor, Room 369 | | | |  |
|  |  | Boston, MA, 02119, USA. | | | |  |
|  |  | +1 (617) 358-3122 | | |  |  |
|  |  | | | | |  |
| **ACADEMIC TRAINING:** | | |  |  |  |  |
| 2016 |  | Ph.D. | | School of Public Health, University of Witwatersrand (Johannesburg, South Africa) | | |
| 2009 |  | M.Com. | | School of Economic and Business Sciences, University of Witwatersrand (Johannesburg, | | |
|  |  |  |  | South Africa), Economics, *cum laude* | | |
| 2001 |  | B.Bus.Sci. | | School of Economics, University of Cape Town (Cape Town, South Africa), Economics and | | |
|  |  |  |  | Finance | |  |
| **ACADEMIC LEADERSHIP:** | | | |  |  |  |
| 2011 | - 2017 | Deputy Division Head | | | | Health Economics & Epidemiology Research Office |
|  |  |  |  |  |  | Wits Health Consortium |
|  |  |  |  |  |  | Faculty of Health Sciences, University of Witwatersrand |
| 2008 | – 2011 | Research Coordinator | | | | Health Economics & Epidemiology Research Office |
|  |  |  |  |  |  | Wits Health Consortium |
|  |  |  |  |  |  | Faculty of Health Sciences, University of Witwatersrand |
| **RESEARCH APPOINTMENTS:** | | | |  |  |  |
| 2017 | – Present | Research Assistant Professor | | | | Department of Global Health, School of Public Health, |
|  |  |  |  |  |  | Boston University |
| 2008 | – Present | Associate Researcher | | | | Joint Faculty Appointment, School of Clinical Medicine, |
|  |  |  |  |  |  | Faculty of Health Sciences, University of the Witwatersrand |
| 2008 | – Present | Principal Researcher (Visiting) | | | | Health Economics & Epidemiology Research Office |
|  |  |  |  |  |  | Wits Health Consortium |
|  |  |  |  |  |  | Faculty of Health Sciences, University of Witwatersrand |
| 2006 | – 2007 | Research Associate | | | | Health Economics & Epidemiology Research Office |
|  |  |  |  |  |  | Wits Health Consortium |
|  |  |  |  |  |  | Faculty of Health Sciences, University of Witwatersrand |
| 2005 |  | Study Coordinator | | | | Health Economics & Epidemiology Research Office |
|  |  |  |  |  |  | Wits Health Consortium |
|  |  |  |  |  |  | Faculty of Health Sciences, University of Witwatersrand |
| **OTHER RESEARCH EXPERIENCE:** | | | |  |  |  |
| 2007 | – 2008 | Consultant | | | | Center for International Health and Development, |
|  |  |  |  |  |  | Boston University School of Public Health |
|  |  |  |  |  |  | Project: *Cost and Outcomes of Models for Delivering Antiretroviral* |
|  |  |  |  |  |  | *Therapy for HIV/AIDS in Zambia* |
| 2007 | – 2008 | Consultant | | | | Center for International Health and Development, |
|  |  |  |  |  |  | Boston University School of Public Health |
|  |  |  |  |  |  | Project: *Cost and Outcomes of Models for Delivering Antiretroviral* |
|  |  |  |  |  |  | *Therapy for HIV/AIDS in Kenya* |

| **TEACHING EXPERIENCE:** | |  |
| --- | --- | --- |
| 2019 |  | **Structural factors affecting HIV – an economists perspective.** Audience: MPH Students. Guest |
|  |  | Lecture in Individual, Community, and Population Health SPH GH 720 Boston University, Boston, |
|  |  | USA. Presented in two classes (Profs Monica Onyango & Jennifer Schlezinger). |
| 2019 |  | **Economic evaluation.** Audience: MPH Students. Guest Lecture in Monitoring and Evaluation of |
|  |  | Global Health Programs SPH GH 745, Boston University, Boston, USA. |
| 2019 |  | **Using economics to influence policy.** Audience: MPH Students. Guest Lecture in Essential of |
|  |  | Economics and Finance for Global Health SPH GH 762, Boston University, Boston, USA. |
| 2017 |  | **From cost to clinic – Economics changing health policy.** Audience: MPH Students. Guest |
|  |  | Lecture in Essentials of Economics and Finance for Global Health SPH GH 762, Boston University, |
|  |  | Boston, USA. |
| 2016 |  | **Data collection and analysis for economic evaluations.** Audience: Technical implementing |
|  |  | partners. EQUIP Partners Meeting, Johannesburg, South Africa. |
| 2016 |  | **From patient to policy – Ensuring that your clinical practice is positioned to inform evidenced** |
|  |  | **based policy.** Audience: HIV Clinicians. Chair of Research Skills Building Session, Southern African |
|  |  | HIV Clinicians Society Conference, Johannesburg, South Africa. |
| 2014 |  | **Introduction to Health Programme Evaluation**. Audience: MSc students – Module of |
|  |  | Epidemiology for Health Researchers II. University of Witwatersrand, Johannesburg. |

| **INVITED PRESENTATION:** |  |
| --- | --- |
| 2019 | **Costs and resources needed to provide HIV services to key populations over the next 10 years.** |
|  | Idea creation meeting – Defining and addressing HIV treatment and prevention needs of underserved |
|  | and high-risk populations. BMGF & Journal of International AIDS Society, New York, USA. |
| 2019 | **Direct action to achieve a result.** Spotlight on “Think, Teach, Do”, School of Public Health, Boston |
|  | University, Boston, USA. |
| 2019 | **Learning Community – Take home.** Academy for Faculty Advancement, School of Medicine, |
|  | Boston University, Boston, USA. |
| 2018 | **Urban public health issues – the transition to Boston.** Health Economics and Epidemiology |
|  | Research Office, Johannesburg, South Africa. |
| 2017 | **Partner’s Area of Expertise – Health Economics.** USAID South Africa, Partners Meeting, Pretoria, |
|  | South Africa. Presented in absentia by Denise Evans. |
| 2017 | **Innovations Research on AIDS (INROADS).** Director Doug Arbuckle, Office HIV AIDS (OHA, |
|  | USA). USAID, Johannesburg, South Africa. Presented in absentia by Denise Evans. |
| 2016 | **Test and Start – Research supporting evidence based policy change.** Zambian Department of |
|  | Health & USAID, EQUIP Project, Johannesburg, South Africa. |
| 2016 | **Initiating ART at a patients first clinic visit: the RapIT randomised trial.** Faculty of Health |
|  | Sciences Research Day, University of Witwatersrand, Johannesburg, South Africa. |

**CONFERENCE ORAL PRESENTATIONS:**

**BIBLIOGRAPHY:**

| ORCID ID: | 0000-0003-4986-4988 |
| --- | --- |
| Google Scholar: | https://scholar.google.co.za/citations?user=fao8zDgAAAAJ&hl=en&oi=ao |
| **Peer reviewed publications:** | |

1. **Long L**, Kuchukhidze S, Pascoe S, Nichols B, Cele R, Govathson C, Huber A, Flynn D, Rosen S. Differentiated models of service delivery for antiretroviral treatment of HIV in sub-Saharan Africa: a rapid review protocol. *Systematic Reviews*. DOI: 10.1186/s13643-019-1210-6. 2019.
2. Jamieson L, Evans D, Berhanu R, Ismail N, Aucock S, Wallengren K, **Long L**. Data quality of drug-resistant tuberculosis and antiretroviral therapy electronic registers in South Africa. *BMC Public Health*. DOI: 10.1186/s12889-019-7965-9. 2019.
3. Mokhele I, Mashamaite S, Majuba P, Xulu T, **Long L**, Onoya D. Effective public-private partnerships for sustainable antiretroviral therapy: outcomes of the Right to Care health services GP down-referral program. *BMC Public Health*. DOI:

10.1186/s12889-019-7660-x. 2019.

4. Lince-Deroche N, Leuner R, Meyer-Rath G, Pillay Y, **Long L**. When Donor Funding Leaves: A retrospective review of the impact of integrating direct HIV care and treatment into public health services in a region of Johannesburg. *Cost Effectiveness* *and Resource Allocation*. DOI: 10.1186/s12962-019-0192-5. 2019.
5. Musakwa N, Feeley A, Magwete M, Patz S, McNamara L, Sanne I, **Long L**, Evans D. Dietary intake among paediatric HIV-positive patients initiating antiretroviral therapy in Johannesburg, South Africa. *Vulnerable Children and Youth Studies (RVCH).* DOI: 10.1080/17450128.2019.166858. 2019.
6. Brennan A, Bonawitz R, Gill CJ, Thea DM, Kleinman M, **Long L**, McCallum C, Fox MP. A Meta-analysis Assessing Diarrhea and Pneumonia in HIV-Exposed Uninfected Compared With HIV-Unexposed Uninfected Infants and Children. *Journal of Acquired Immune Deficiency Syndrome (JAIDS)*. DOI: 10.1097/QAI.0000000000002097. 2019
7. Berry KM, Rodriguez CA, Berhanu R, Ismail N, Rosen S, **Long L**, Evans D. Treatment outcomes among pediatrics, adolescents, and adults on treatment for drug-sensitive TB in two metropolitan municipalities in Gauteng Province, South Africa. *BMC Public Health.* DOI: 10.1186/s12889-019-7257-4. 2019.
